# The impact of vaccination on preventing long COVID in the Omicron era: a systematic review and meta-analysis

**DOI:** 10.1101/2024.11.19.24317487

**Authors:** Rhiannon Green, Zoe Marjenberg, Gregory Y. H. Lip, Amitava Banerjee, Juan Wisnivesky, Brendan C. Delaney, Michael J. Peluso, Elke Wynberg, Sultan Abduljawad

**Author notes:** **Corresponding author:** Sultan Abduljawad.

## Abstract

Millions worldwide are living with long COVID. Since therapeutic research is ongoing, long COVID prevention is a pragmatic public health strategy. While prior analyses have shown the benefit of primary vaccination, the effect of booster vaccination on preventing long COVID caused by an Omicron infection has not been fully investigated. This systematic review identified 31 observational studies, among which 11 were deemed suitable for pairwise meta-analyses. Herein, the pooled risk of long COVID was 22–29% (*P*<0.0001 and *P*<0.0001, respectively) lower for vaccinated (any vaccination) populations versus unvaccinated (10 studies); 19% (*P*<0.0001) lower for primary course vaccination versus no vaccination (3 studies); 26% (*P*<0.0001) lower for booster vaccination versus no vaccination (4 studies) and 23% (*P*=0.0044) lower for booster vaccination versus primary course vaccination (3 studies). These findings indicate that booster vaccination can provide additional protection against long COVID; thereby, highlighting the importance of seasonal vaccination against new SARS-CoV-2 variants.

## Introduction

In 2022, the World Health Organization (WHO) estimated that roughly 10–20% of individuals experience persistent symptoms following the acute phase of SARS-CoV-2 infection^1^. More recent reports, from 2024, place the proportion at 2–7%^2,3^, alluding to potential era-related and vaccination effects^4^. This condition is known as long COVID and, in certain contexts, is referred to as post-acute sequelae of COVID-19 (PASC), post-COVID condition (PCC), or post-acute COVID-19 syndrome (PACS)^5,6^. The US National Academies of Sciences, Engineering, and Medicine recently published a consensus definition for long COVID as “an infection-associated chronic condition that occurs after SARS-CoV-2 infection and is present for at least 3 months as a continuous, relapsing and remitting, or progressive disease state that affects one or more organ systems”^7^.

Long COVID is a multifaceted disease with differing pathologies that may result in overlapping symptoms. Symptoms can manifest in almost any aspect of physical or mental health, making management more difficult^2,8^. Living with symptoms of long COVID can negatively impact work productivity and social relationships, contributing to a general decline in quality of life^9–11^. In particular, fatigue, cognitive dysfunction, insomnia, and inability to exercise have been reported as some of the symptoms with greatest burden by participants in a cross-sectional survey study (n=5163) conducted in an online COVID-19 support group^11^. A previous meta-analysis reported that across studies, an average of 52% of people with long COVID (defined as persistent effects after 3 months) had a long-lasting decrease in health-related quality of life (HRQoL), with a mean follow-up of 4.5 months^9^. This decline in HRQoL was exacerbated in participants who experienced severe COVID-19 compared to those with milder disease^9^. In relation to overall disease burden, long COVID was found to account for 80.4 and 642.8 daily adjusted life years per 1000 non-hospitalized and hospitalized persons, respectively^6^. Further, long COVID has lasting economic implications and places significant burden on healthcare systems, largely due to hospitalization costs and healthcare visits^12^. Al-Aly et al (2024) reported a conservative estimate based on available data that places the annual global economic cost of long COVID at $1 trillion, which equates to 1% of the global gross domestic product for 2024^13^.

Currently, there are no validated, effective treatments for long COVID, and management is largely symptomatic^5,7^. This places additional value on preventative measures. Vaccination prior to SARS-CoV-2 infection has been shown to reduce the risk of developing long COVID^14–21^. This is likely due, at least in part, to the protective effect of vaccination against severe acute COVID-19 and COVID-19–related hospitalization. Booster and seasonal vaccines have been introduced to provide greater protection against emerging SARS-CoV-2 variants containing escape mutations, and reduce the risk of severe COVID-19. It follows that booster vaccination may have added benefits in preventing long COVID. Indeed, studies have shown that the effectiveness of vaccination prior to infection in protecting against long COVID is increased with additional booster doses received^22,23^.

Omicron (B.1.1.529) was identified in November 2021 and designated a variant of concern by the WHO^24^. This variant has demonstrated a distinct genomic signature compared with the original Wuhan SARS-CoV-2 strain. Omicron variants, including currently circulating subvariants, display increased transmissibility and infectivity. Evidence suggests an increased ability to evade humoral immunity, suspected to be driven by its extensive mutational profile, resulted in a surge in Omicron sub-variant infections throughout 2022^25^. More recently, in the summer of 2024, prevalence of the Omicron subvariant JN.1 surged, and at the time of writing, circulating JN.1-derived variants were dominant globally^26^.

An up-to-date and comprehensive understanding of the impact of vaccination on the prevalence and risk of long COVID is needed to reflect evolving SARS-CoV-2 strains and changing global immunity due to evolving sub-variants, reinfection, and seasonal vaccination. The majority of previous SLRs investigated the impact of primary course vaccination versus no vaccination or included non-Omicron variants in the analysis^19,20,27,28^. Therefore, there is a gap in understanding the effect of additional booster doses on the risk of developing long COVID in the context of more recent Omicron-variant data. This systematic review aimed to investigate the impact of COVID-19 vaccination, including booster versus primary course vaccination, on preventing long COVID caused by Omicron variants infection.

## Methods

This study was conducted in accordance with the Meta-analysis of Observational Studies in Epidemiology and the Preferred Reporting Items for Systematic Reviews and Meta-analyses (PRISMA) guidelines for conducting and reporting systematic reviews^29^. The study protocol was registered with PROSPERO (CRD42024501445).

### Data sources and searches

The following databases were used to identify relevant studies from 1 January 2022 to 1 March 2024: Embase, MEDLINE, PubMed, Europe PMC (including medRxiv and bioRxiv reprints), Latin American and Caribbean Health Sciences Literature (LILACS), Cochrane COVID-19 Study Register, and the WHO COVID-19 Database. Search strategies were developed using search terms related to long COVID and vaccination status. The complete search strategies are detailed in **Supplementary Methods (1.1 Search Strategy)**. Hand searching was also performed, and bibliographies of identified studies were checked.

### Eligibility criteria, screening, and data extraction

Studies considered eligible for inclusion in the review met all the following criteria:

#### Population

participants (all ages) with Omicron variant SARS-CoV-2 infection. Infections were considered Omicron variants if studies either reported infections to be caused only by Omicron variants, included infections only occurring after November 2021, or reported infections that were only caused by B.1.1.529 and all subsequent sub-variants arising from the Omicron lineage.

#### Intervention

participants who received BNT162b2, mRNA-1273, Ad26.CoV2.S, ChAdOx1, or other European Union (EU)-authorized COVID-19 vaccines before SARS-CoV-2 infection.

#### Comparison

either participants who were unvaccinated (for comparison with vaccinated) or received at least one fewer vaccine dose than the intervention population.

#### Outcomes

prevalence or incidence of long COVID (percentage of participants with symptoms, incidence rates), severity outcomes or symptom burden of long COVID disease (any measure of long COVID severity, average number of long COVID symptoms), and the risk of long COVID (e.g., odds ratio [OR], hazard ratio, risk ratio, rate ratio, incidence rate ratio, or vaccine effectiveness estimates). In this study, we defined long COVID as any condition or symptom described by the study as long COVID, PASC, PCC, or PACS, provided it was reported at least four weeks after the acute SARS-CoV-2 infection (based on the definition of long COVID used by the Centers for Disease Control and Prevention at the time of analysis^30^).

#### Study design

Observational (cohort, case-control, cross-sectional) studies.

#### Publication type and language

published and pre-print studies with full-texts available in English.

Titles and abstracts of identified references were assessed by two independent reviewers to determine whether they met the inclusion criteria. Any discrepancies were resolved by discussion until a consensus was reached. Full-text screening was performed on the studies that met the criteria for inclusion and data were extracted into a pre-defined table. One reviewer extracted data on study and participant characteristics, and outcomes of interest, and a second reviewer verified the accuracy of extraction.

### Quality assessment

The risk of bias of included studies was assessed using the Newcastle-Ottawa scale (NOS) for cohort and case-control studies^31^ (**Supplementary Table 1),** and by the Joanna Briggs Institute tool for cross-sectional studies^32^. Studies assessed by the NOS were categorized as low, medium, or high risk of bias according to their combined domain (selection, comparability, and outcome) scores, where the maximum score for each study can be 9. Studies were classed as low risk of bias if they scored 7–9 (7–8 for retrospective studies), medium risk if they scored 4–6, and high risk if they scored 0–3. Additionally, if a study did not score at least 1 in any of the three domains it was categorized as high risk.

### Data analysis

Qualitative data synthesis was performed to identify key trends among the studies included in the SLR. Study populations were divided into different vaccination status groups (unvaccinated, primary course vaccination, booster vaccination, and additional booster vaccination) according to the status and number of vaccine doses reported by the studies.

### Statistical analysis

A feasibility assessment was conducted among studies selected through the SLR to determine whether pairwise meta-analyses could be performed for the risk of long COVID outcomes (**Supplementary Methods: 1.2.1 Feasibility assessment**).

Pairwise meta-analyses were conducted to assess the risk of developing long COVID in those who had contracted the Omicron variant of SARS-CoV-2 after receiving a vaccine and/or booster doses compared with those who were unvaccinated or had received primary course vaccination. If multiple timepoints were reported, the timepoint closest to that of other studies in the meta-analysis was used for comparison. Risk outcomes (ORs, hazard ratios, and rate ratios), were considered equal estimates (all referred to as relative risk [RR])^33^. Vaccine effectiveness was converted to OR using OR=1−(VE/100)^34^. Weights were calculated using the inverse variance method (weight=1/variance). DerSimonian and Laird models (random effects)^35^ were fitted to calculate pooled RRs and 95% CI for all outcomes.

Sensitivity analyses were performed to assess the robustness of results, **(Supplementary Methods: 1.2.2 Sensitivity analyses**). Heterogeneity was measured using Cochran’s Q test with statistical significance set as *P*<0.05. The thresholds for interpretation of the I² statistic were defined by the Cochrane Handbook for Systematic Reviews of Interventions^36^. Publication bias was assessed using funnel plots and the Egger’s test^37^ for analyses that included at least 10 studies^36^. All statistical analyses were conducted using R version 4.1.1 with the meta package.

## Results

### Overview of included studies

In total, 9107 records were identified through searches. Following elimination of duplicates, 5360 studies underwent title and abstract screening, of which 368 full-text articles were assessed for eligibility. Following this, 31 studies were selected for the review, including two additional studies identified by hand searching (**Figure 1**).

**Figure 1.**
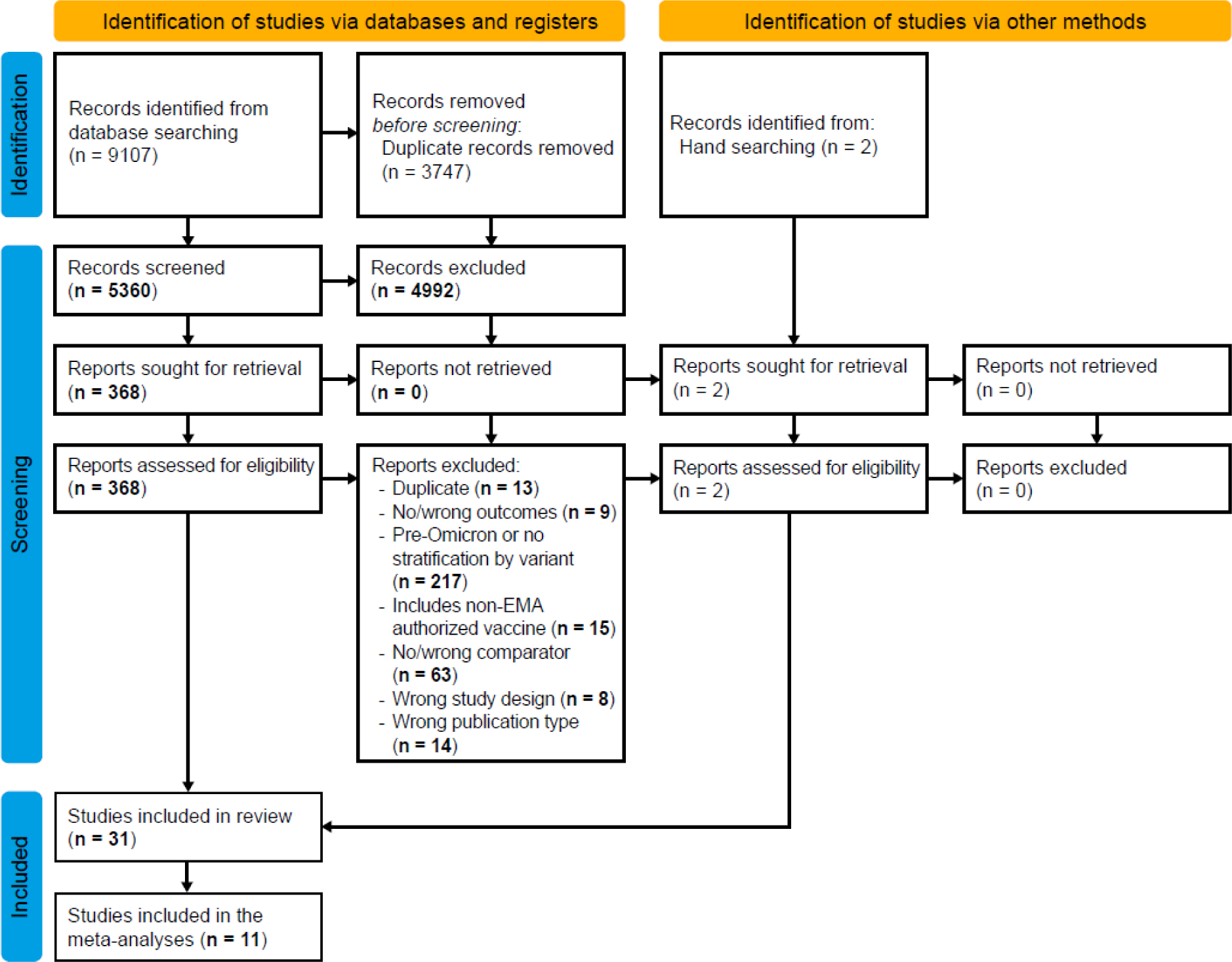
PRISMA flow diagram. EMA, European Medicines Agency

Study characteristics are summarized in **Table 1**. The studies were performed in 16 different countries across Europe, North America, Asia, and Australia. Eleven studies were conducted retrospectively^22,38–46^, 17 were prospective^10,47–62^ and three were cross-sectional^63–65^ in design. Data sources varied across the studies and included medical centers, COVID-19 cohort studies and registries, electronic health records, and online surveys. The definitions of long COVID adopted by the studies varied and are reported in **Table 2**.

**Table 1.**
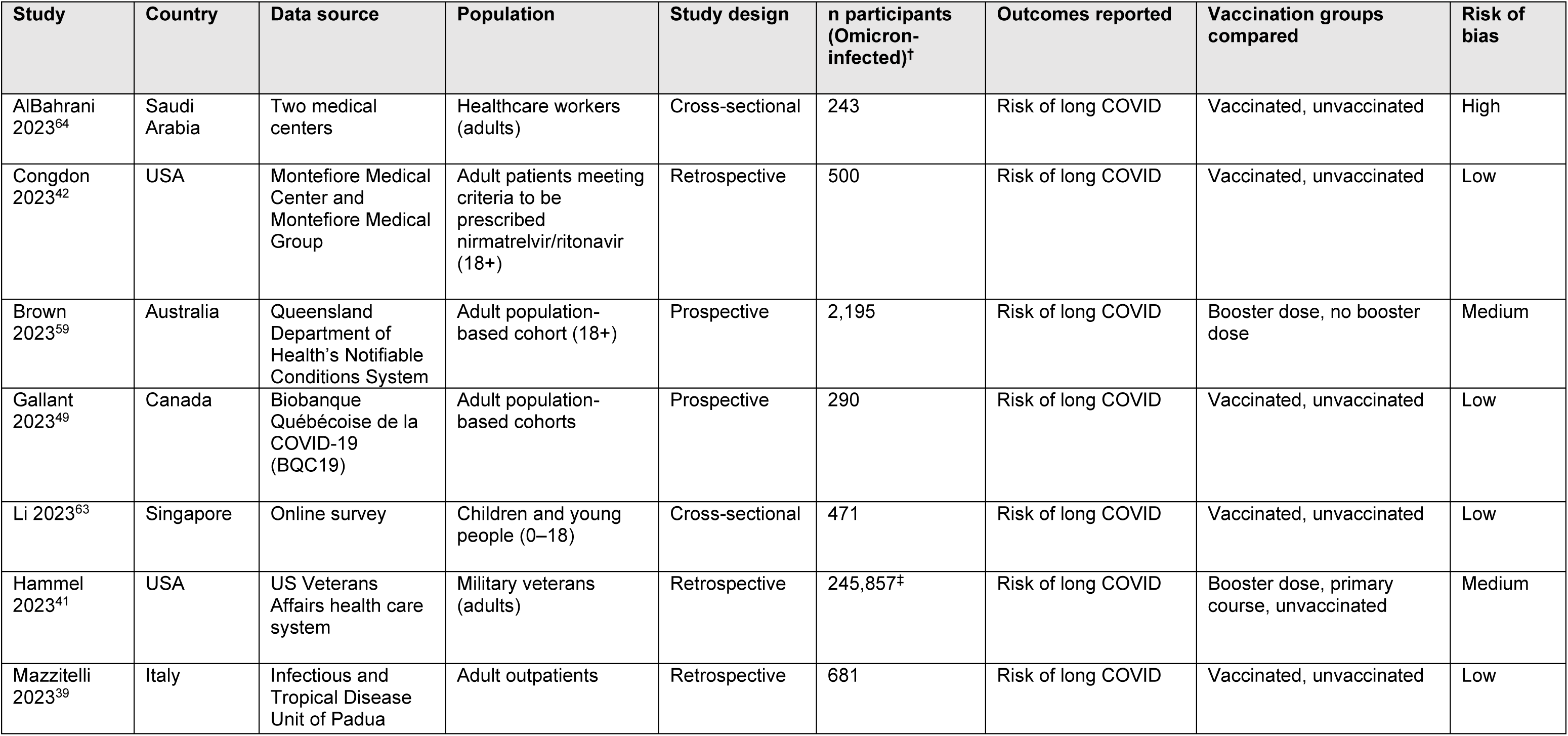
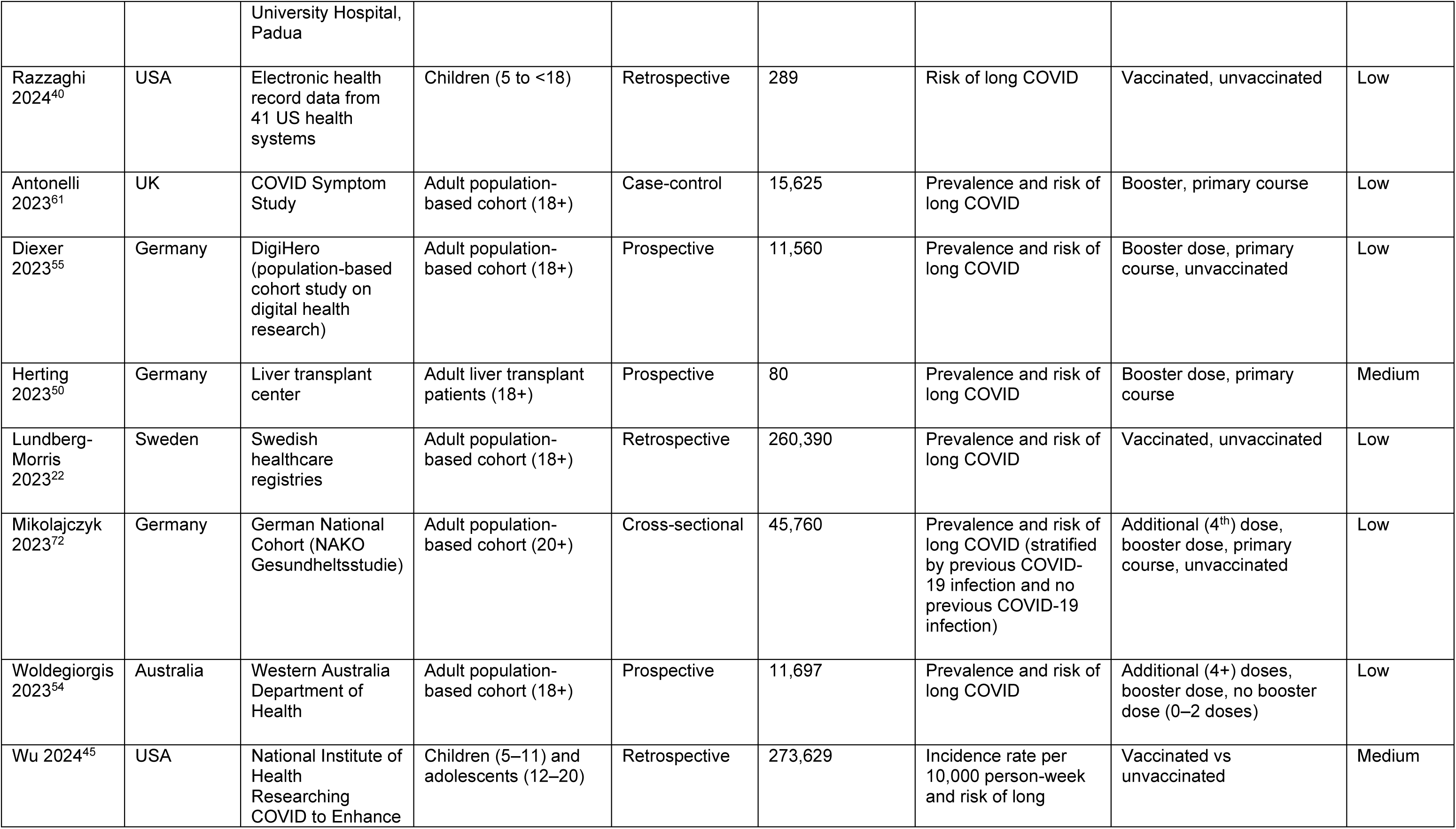
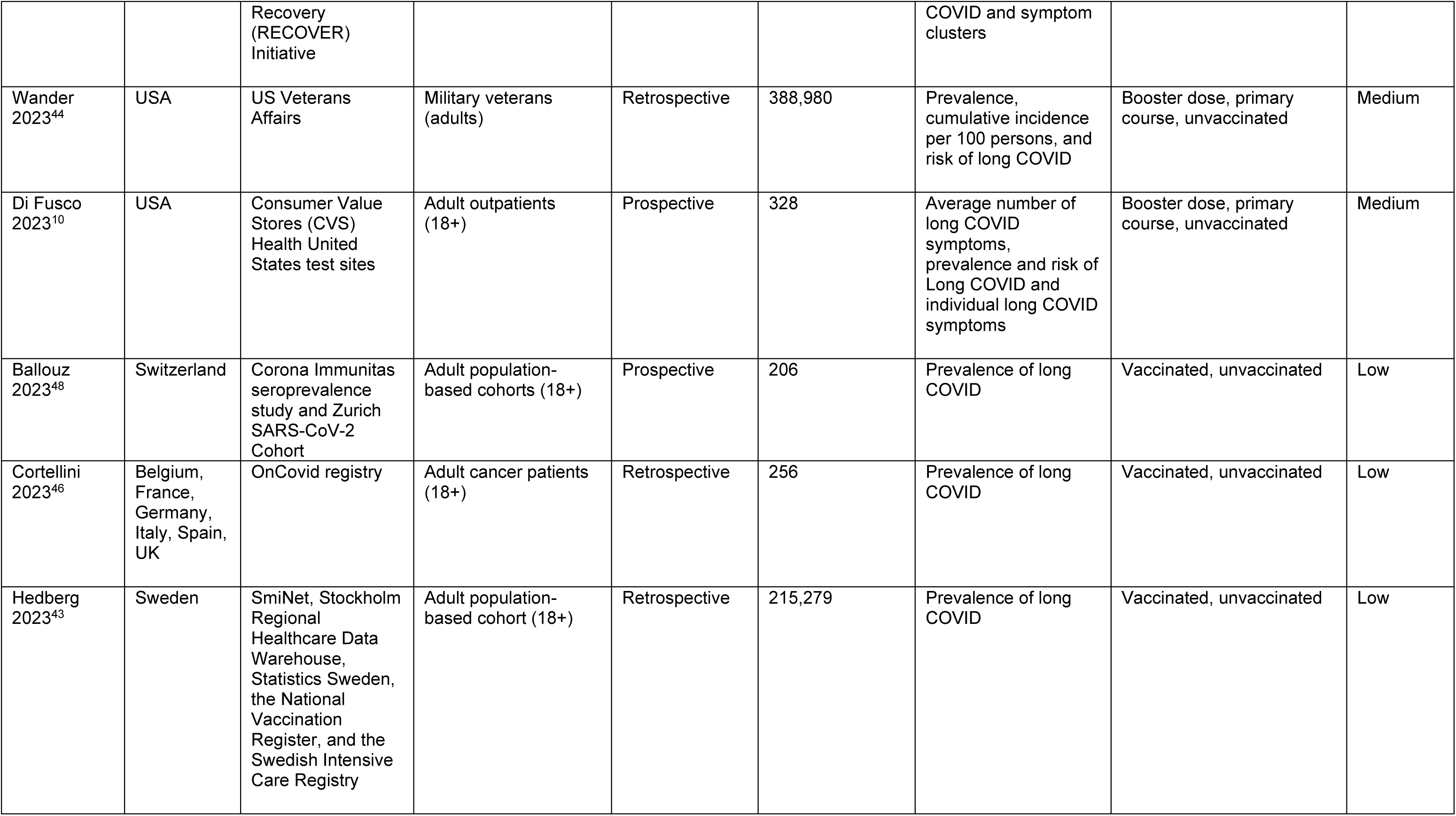
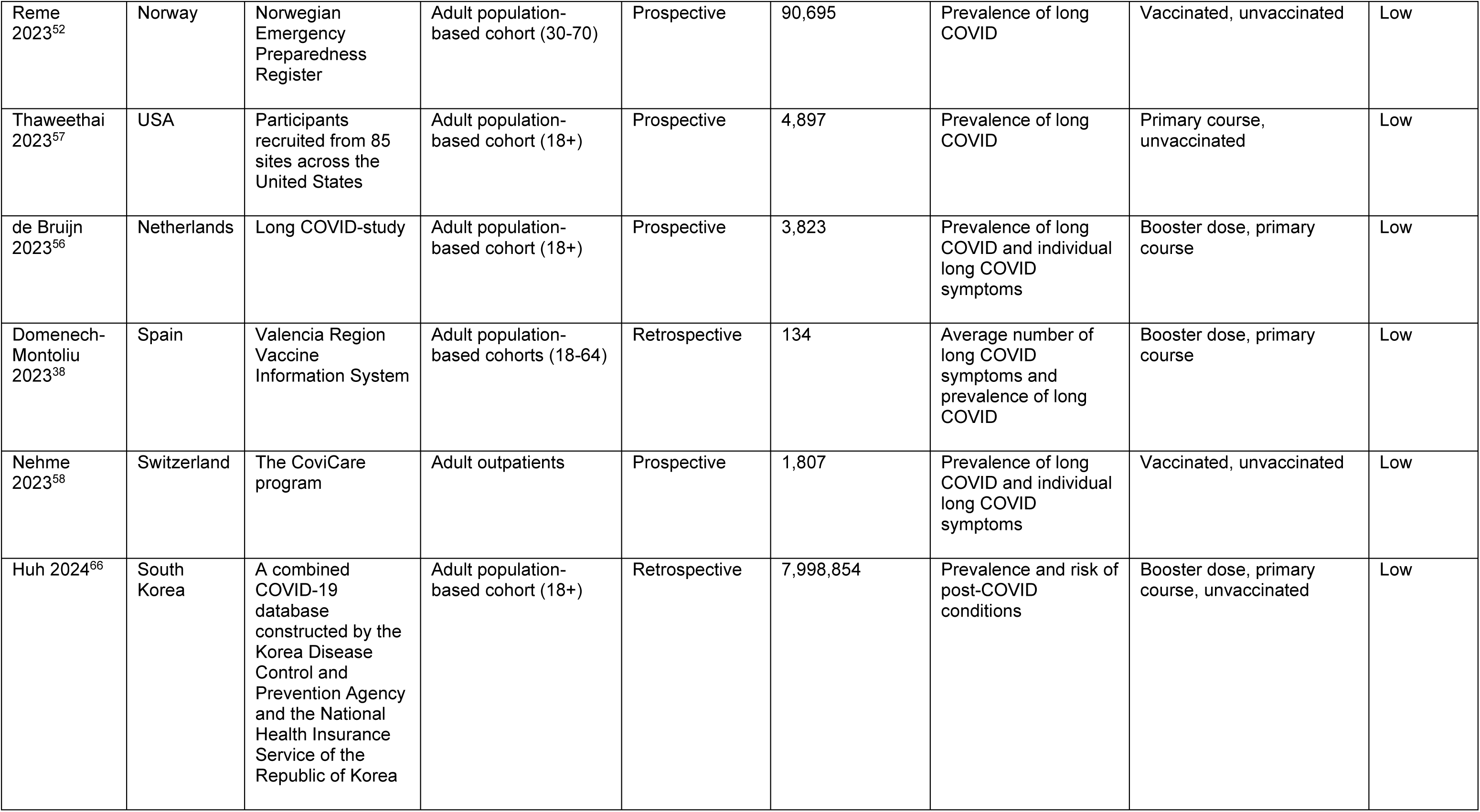
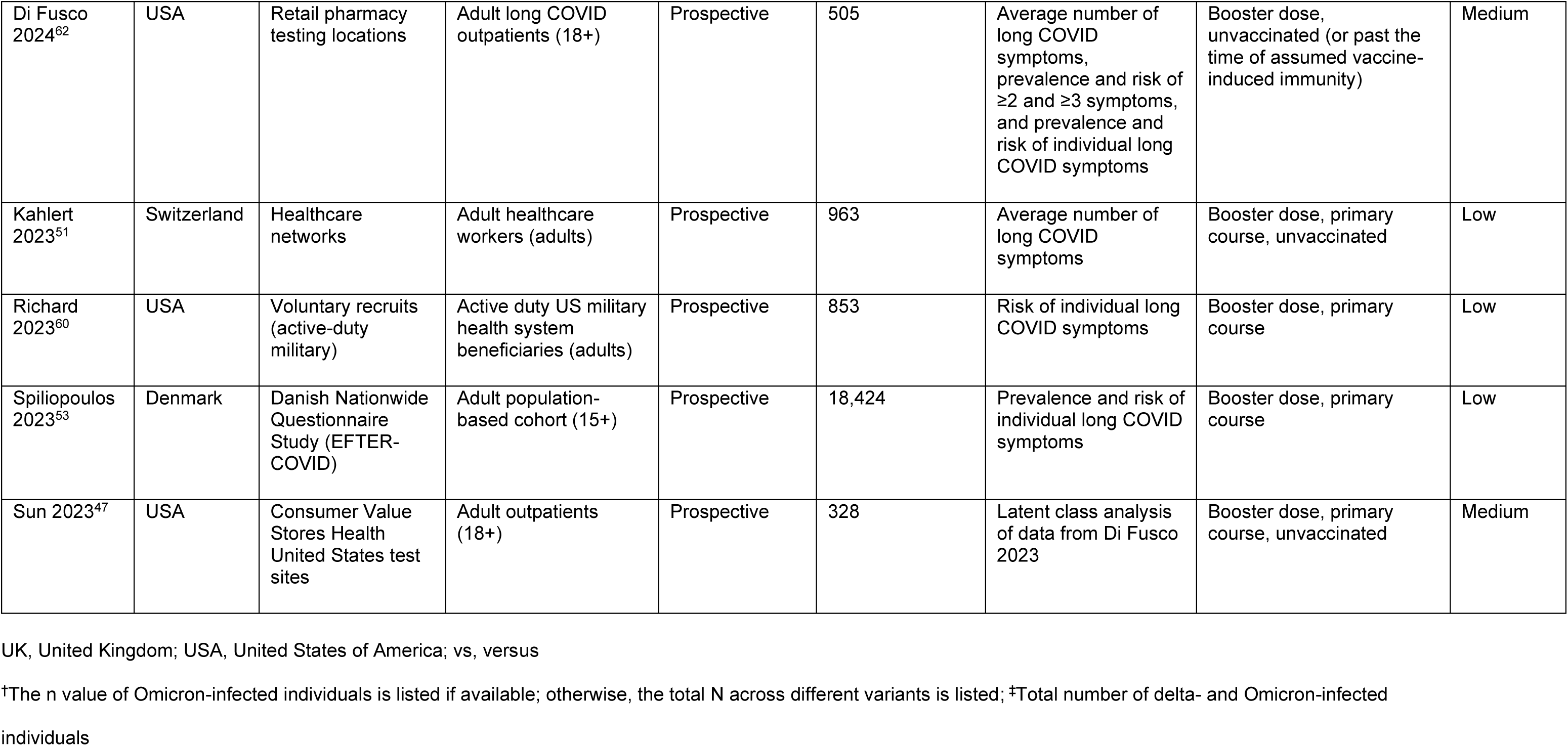
Characteristics of studies included in the SLR.

**Table 2.**
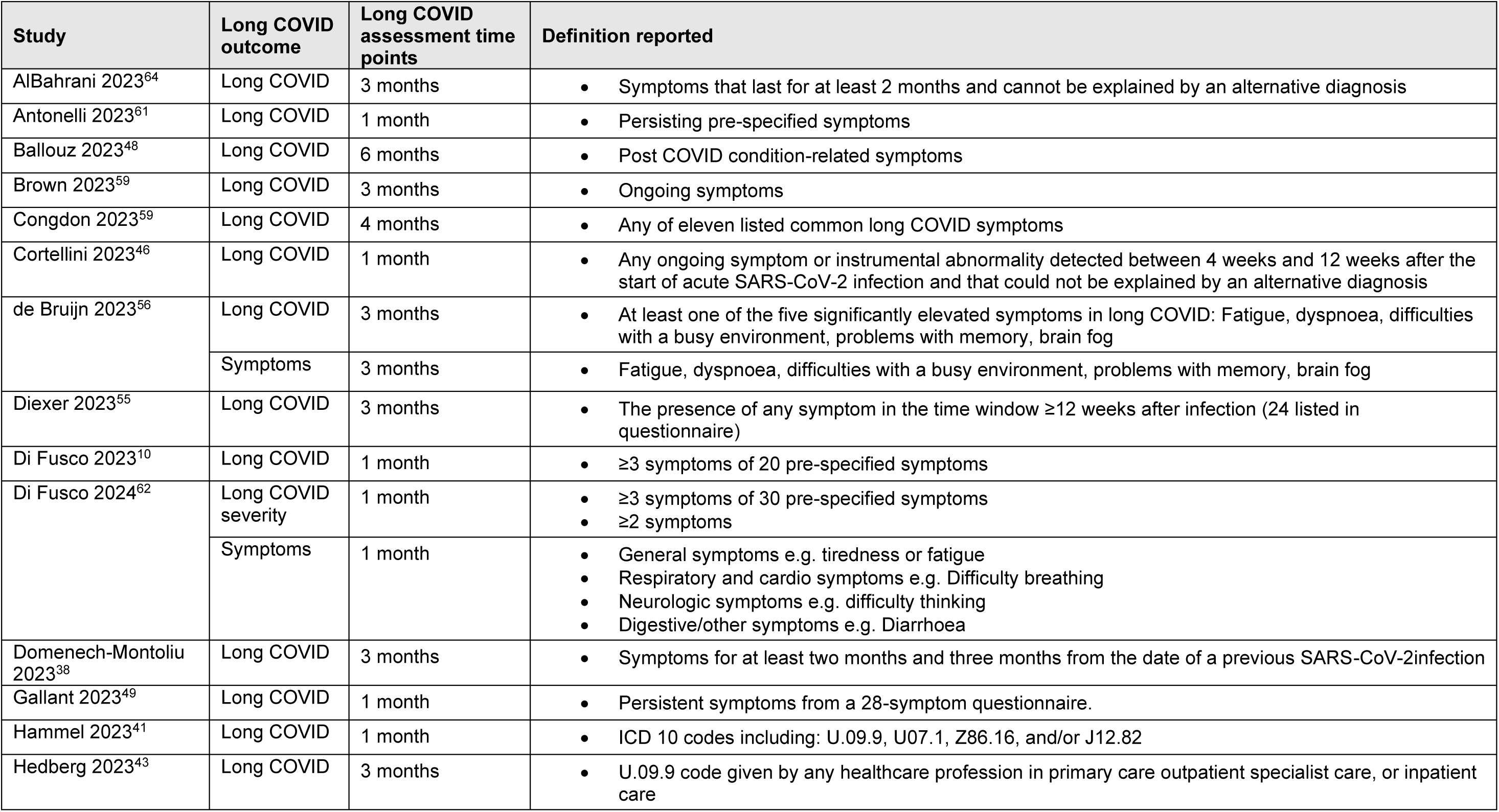
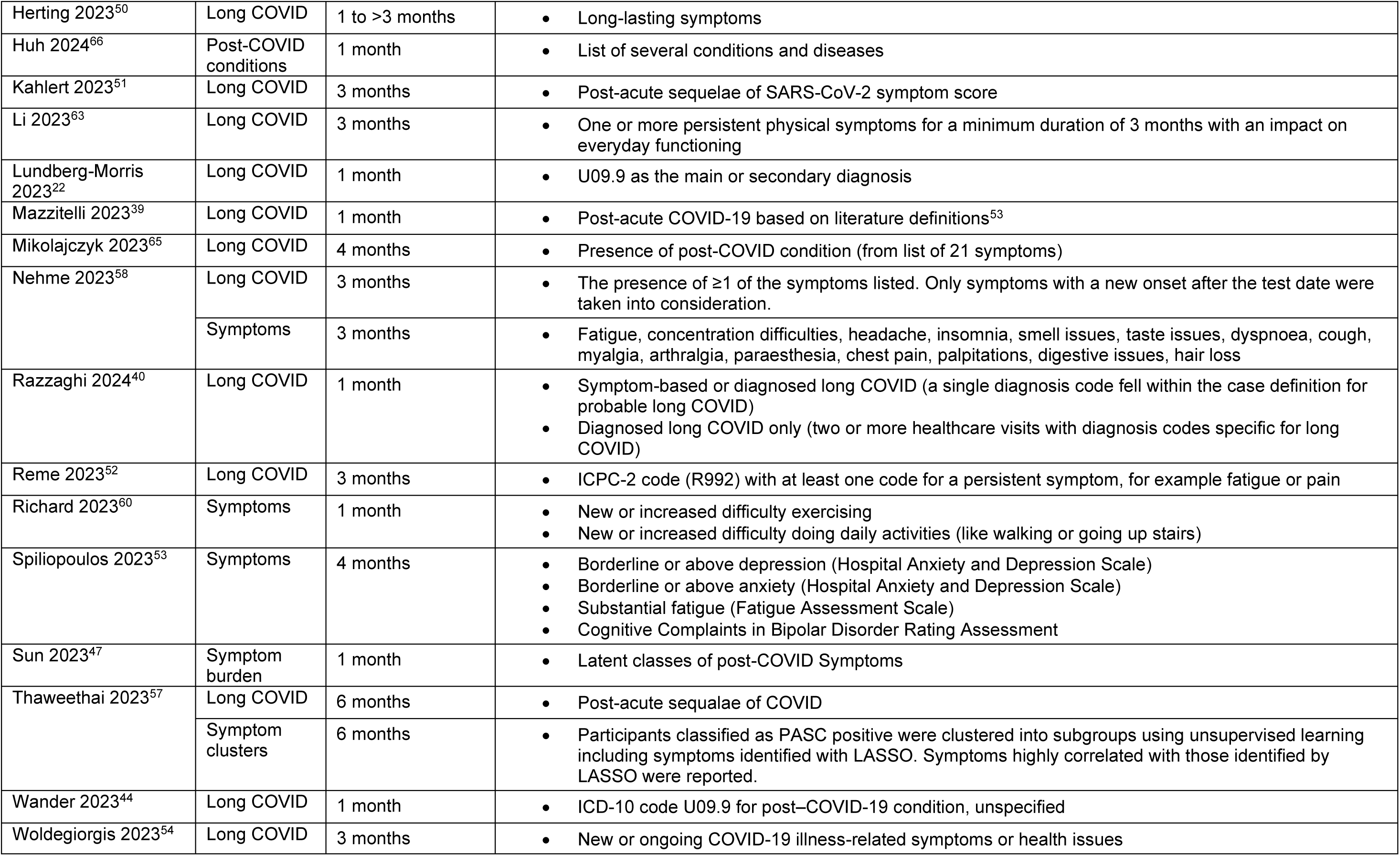
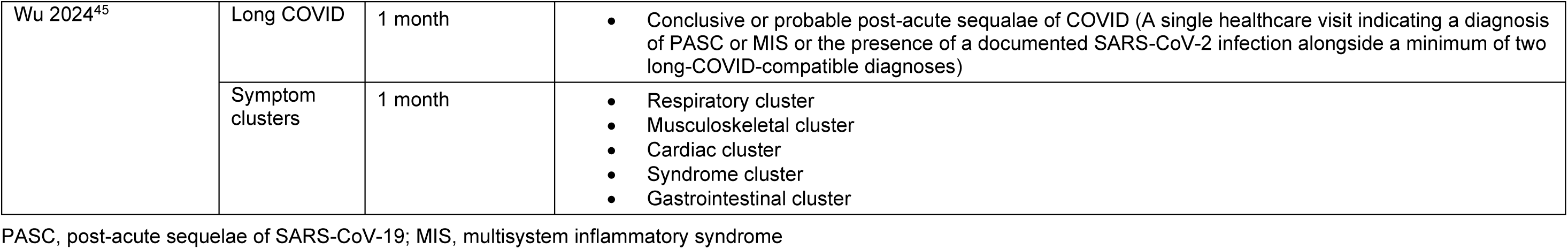
Study outcomes and outcome definitions.

| Study | Long COVID outcome | Long COVID assessment time points | Definition reported |
| --- | --- | --- | --- |
| AlBahrani 2023<sup>64</sup> | Long COVID | 3 months | Symptoms that last for at least 2 months and cannot be explained by an alternative diagnosis |
| Antonelli 2023<sup>61</sup> | Long COVID | 1 month | Persisting pre-specified symptoms |
| Ballouz 2023<sup>48</sup> | Long COVID | 6 months | Post COVID condition-related symptoms |
| Brown 2023<sup>59</sup> | Long COVID | 3 months | Ongoing symptoms |
| Congdon 2023<sup>59</sup> | Long COVID | 4 months | Any of eleven listed common long COVID symptoms |
| Cortellini 2023<sup>46</sup> | Long COVID | 1 month | Any ongoing symptom or instrumental abnormality detected between 4 weeks and 12 weeks after the start of acute SARS-CoV-2 infection and that could not be explained by an alternative diagnosis |
| de Bruijn 2023<sup>56</sup> | Long COVID | 3 months | At least one of the five significantly elevated symptoms in long COVID: Fatigue, dyspnoea, difficulties with a busy environment, problems with memory, brain fog |
|  | Symptoms | 3 months | Fatigue, dyspnoea, difficulties with a busy environment, problems with memory, brain fog |
| Diexer 2023<sup>55</sup> | Long COVID | 3 months | The presence of any symptom in the time window $\geq 12$ weeks after infection (24 listed in questionnaire) |
| Di Fusco 2023<sup>10</sup> | Long COVID | 1 month | $\geq 3$ symptoms of 20 pre-specified symptoms |
| Di Fusco 2024<sup>62</sup> | Long COVID severity | 1 month | $\geq 3$ symptoms of 30 pre-specified symptoms<br>$\geq 2$ symptoms |
|  | Symptoms | 1 month | General symptoms e.g. tiredness or fatigue<br>Respiratory and cardio symptoms e.g. Difficulty breathing<br>Neurologic symptoms e.g. difficulty thinking<br>Digestive/other symptoms e.g. Diarrhoea |
| Domenech-Montoliu 2023<sup>38</sup> | Long COVID | 3 months | Symptoms for at least two months and three months from the date of a previous SARS-CoV-2 infection |
| Gallant 2023<sup>49</sup> | Long COVID | 1 month | Persistent symptoms from a 28-symptom questionnaire. |
| Hammel 2023<sup>41</sup> | Long COVID | 1 month | ICD 10 codes including: U09.9, U07.1, Z86.16, and/or J12.82 |
| Hedberg 2023<sup>43</sup> | Long COVID | 3 months | U.09.9 code given by any healthcare profession in primary care outpatient specialist care, or inpatient care |
| Herting 2023<sup>50</sup> | Long COVID | 1 to >3 months | Long-lasting symptoms |
| Huh 2024<sup>66</sup> | Post-COVID conditions | 1 month | List of several conditions and diseases |
| Kahlert 2023<sup>51</sup> | Long COVID | 3 months | Post-acute sequelae of SARS-CoV-2 symptom score |
| Li 2023<sup>63</sup> | Long COVID | 3 months | One or more persistent physical symptoms for a minimum duration of 3 months with an impact on everyday functioning |
| Lundberg-Morris 2023<sup>22</sup> | Long COVID | 1 month | U09.9 as the main or secondary diagnosis |
| Mazzitelli 2023<sup>39</sup> | Long COVID | 1 month | Post-acute COVID-19 based on literature definitions<sup>53</sup> |
| Mikolajczyk 2023<sup>65</sup> | Long COVID | 4 months | Presence of post-COVID condition (from list of 21 symptoms) |
| Nehme 2023<sup>58</sup> | Long COVID | 3 months | The presence of $\geq 1$ of the symptoms listed. Only symptoms with a new onset after the test date were taken into consideration. |
|  | Symptoms | 3 months | Fatigue, concentration difficulties, headache, insomnia, smell issues, taste issues, dyspnoea, cough, myalgia, arthralgia, paraesthesia, chest pain, palpitations, digestive issues, hair loss |
| Razzaghi 2024<sup>40</sup> | Long COVID | 1 month | Symptom-based or diagnosed long COVID (a single diagnosis code fell within the case definition for probable long COVID)<br>Diagnosed long COVID only (two or more healthcare visits with diagnosis codes specific for long COVID) |
| Reme 2023<sup>52</sup> | Long COVID | 3 months | ICPC-2 code (R992) with at least one code for a persistent symptom, for example fatigue or pain |
| Richard 2023<sup>60</sup> | Symptoms | 1 month | New or increased difficulty exercising<br>New or increased difficulty doing daily activities (like walking or going up stairs) |
| Spiliopoulos 2023<sup>53</sup> | Symptoms | 4 months | Borderline or above depression (Hospital Anxiety and Depression Scale)<br>Borderline or above anxiety (Hospital Anxiety and Depression Scale)<br>Substantial fatigue (Fatigue Assessment Scale)<br>Cognitive Complaints in Bipolar Disorder Rating Assessment |
| Sun 2023<sup>47</sup> | Symptom burden | 1 month | Latent classes of post-COVID Symptoms |
| Thaweethai 2023<sup>57</sup> | Long COVID | 6 months | Post-acute sequelae of COVID |
|  | Symptom clusters | 6 months | Participants classified as PASC positive were clustered into subgroups using unsupervised learning including symptoms identified with LASSO. Symptoms highly correlated with those identified by LASSO were reported. |
| Wander 2023<sup>44</sup> | Long COVID | 1 month | ICD-10 code U09.9 for post-COVID-19 condition, unspecified |
| Woldegiorgis 2023<sup>54</sup> | Long COVID | 3 months | New or ongoing COVID-19 illness-related symptoms or health issues |
| Wu 2024<sup>45</sup> | Long COVID | 1 month | • Conclusive or probable post-acute sequelae of COVID (A single healthcare visit indicating a diagnosis of PASC or MIS or the presence of a documented SARS-CoV-2 infection alongside a minimum of two long-COVID-compatible diagnoses) |
|  | Symptom clusters | 1 month | • Respiratory cluster<br>• Musculoskeletal cluster<br>• Cardiac cluster<br>• Syndrome cluster<br>• Gastrointestinal cluster |

PASC, post-acute sequelae of SARS-CoV-19; MIS, multisystem inflammatory syndrome

Most studies identified in the SLR included adults only (27 studies^10,22,38,39,41–44,46–52,54–62,64–66^). One study included adolescents (≥15 years old) and adults^53^, and three studies included children and adolescents only^40,45,63^. Of the adult-only studies, the mean age ranged between 36.1 (standard deviation [SD] 7.6) and 61.4 (SD 16.1) years old, and the median ranged from 37 (interquartile range [IQR] 23.0, 48.0) to 66 (IQR 54, 77) years old. The percentage of female participants ranged from 12.7% to 82.0%.

Study population sizes of Omicron-infected participants ranged from 80 to 7,998,854. The Omicron infection period ranged from 13 days to 15 months. Only seven studies reported the specific Omicron subvariant present^38,50,51,58,62,63,66^ and these included BA.1, BA.2, BA.4, BA.5, and XBB. The severity of SARS-CoV-2 infection was reported by 13 studies, where hospitalization rate ranged from 0.1% to 10.3% and the rate of intensive care requirement ranged from 0.02% to 13.8%, across study cohorts. No studies stratified outcomes by acute infection severity.

Of the studies that recorded vaccine types, the following vaccines were reported: BNT162b2, mRNA-1273, Ad26.COV2.S, and ChAdOx1. Four studies included only participants who had received the BNT162b2 vaccine^10,45,47,62^, three included a mixture of mRNA vaccines only^40,49,58^ and 10 reported inclusion of a mixture of EU-authorized vaccines. Two studies included almost entirely mRNA vaccines apart from two patients who received Sinovac^63^ and one patient who received Ad26.COV2.S^48^, respectively. Twelve studies did not report vaccine type but were assumed to include a mixture of EU-authorized vaccines as they were performed in countries that only used EU-authorized vaccines at the time of the study.

### Risk of bias

Risk of bias was found to be either low or medium (20 and 8 studies, respectively) in all studies assessed using NOS (**Supplementary Table 2**). Three cross-sectional studies were analyzed for potential risk of bias using the JBI Critical Appraisal Checklist for Analytical Cross-Sectional Studies, with two low risk^63^ and one potentially high risk due to a lack of reporting and accounting for confounding factors^64,65^.

### Vaccination status

This review classified vaccination status reported by the included studies (full details in **Supplementary Table 3**) into unvaccinated, primary course vaccination only, and booster vaccinated groups. Primary course vaccination was frequently defined by studies as participants receiving two doses, while booster vaccination status was generally defined as participants receiving ≥3 doses (additional booster doses reported by some studies included a total of four or more vaccine doses). There was an exception with the Ad26.COV2.S vaccine, where a single dose was considered a primary course and subsequent vaccination was considered a booster dose. Study participants were generally considered unvaccinated if they had not received any vaccine doses; however, one study also considered participants to be unvaccinated if they received their last monovalent dose more than 12 months before study enrolment^62^.

### Qualitative data synthesis: Outcomes in long COVID

Reported long COVID outcomes assessed by this review were symptom burden (mean number of symptoms) of long COVID (five studies), the prevalence or incidence of long COVID (18 studies), and the risk of long COVID (20 studies). Overall, participants who received primary course vaccination, booster vaccination, and additional booster vaccine doses generally had a numerically or statistically significantly lower number of long COVID symptoms, lower prevalence, and lower risk of long COVID compared with either those who received no vaccination, or vaccination with a lower number of doses. The long COVID outcomes reported by these studies are presented in **Supplementary Tables 4 to 8**.

### Meta-analyses: Impact of vaccination on prevention of long COVID caused by Omicron variant infection

The feasibility assessment identified 11/31 studies^38,43,46–48,52,56–58,62^ that could be included in meta-analyses of the impact of vaccination on risk of developing long COVID after Omicron infections (**Supplementary Results: 2.1 Meta-analysis feasibility assessment and Supplementary Table 9)**

### Impact of vaccination prior to Omicron variant infection on long COVID compared with no vaccination

Ten studies were included in a pooled analysis of the risk of long COVID after vaccination (any number of doses) compared with no vaccination.

Three studies^10,41,44^ reported separate results for both primary vaccination and booster vaccination, which prevented us from conducting one single, pooled analysis of “any vaccination” versus no vaccination across all 10 studies. Therefore, two separate analyses were performed according to the separate results reported in the aforementioned three studies: analysis A included the primary vaccination outcome and analysis B included the booster dose outcome. In each analysis, the pooled estimate from the seven non-stratified studies was taken into account.

Vaccination was associated with a significantly lowered pooled risk of long COVID compared with unvaccinated cohorts by both analysis A (RR 0.78 [95% CI, 0.71–0.85; *P*<0.0001]; **Figure 2**) and B (RR 0.71 [95% CI, 0.64–0.80; *P*<0.0001]; **Figure 3**). Between-study heterogeneity for both analyses was high (I^2^ of 73% and 76%, respectively).

**Figure 2.**
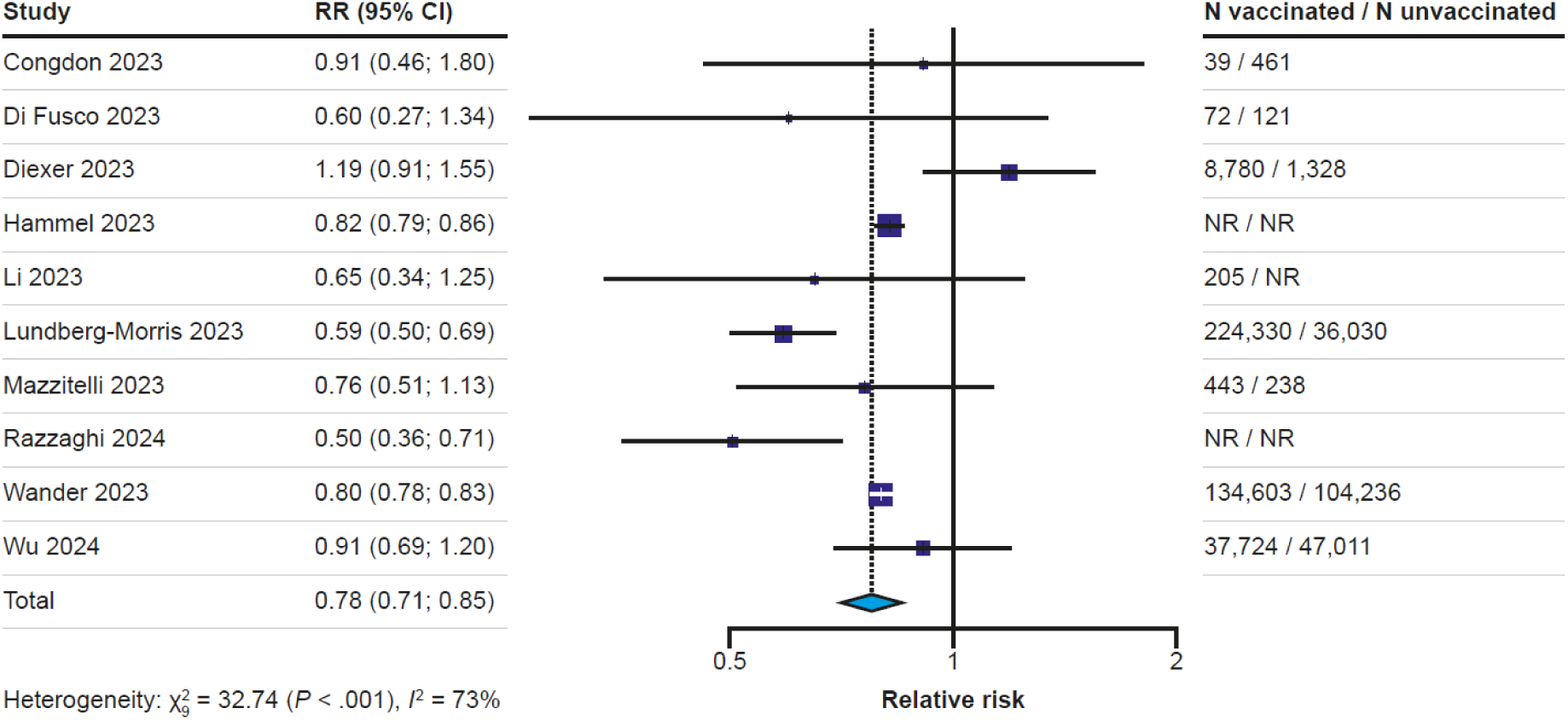
Forest plot for the effect of “any vaccination” (analysis A) on the risk of long COVID compared with no vaccination (random effects) CI, confidence interval; NR, not reported; RR, relative risk. Two separate analyses were performed for “any vaccination” versus no vaccination to account for stratification of results in three studies by primary course vaccination and booster vaccination; analysis A (presented here) included the primary course vaccination data for those stratified studies. In the forest plot, the squares and horizontal bars represent the relative risk and 95% CI, respectively, for each study. The dotted line and diamond represent the pooled relative risk and 95% CI, respectively, for all the studies included in the analysis.

**Figure 3.**
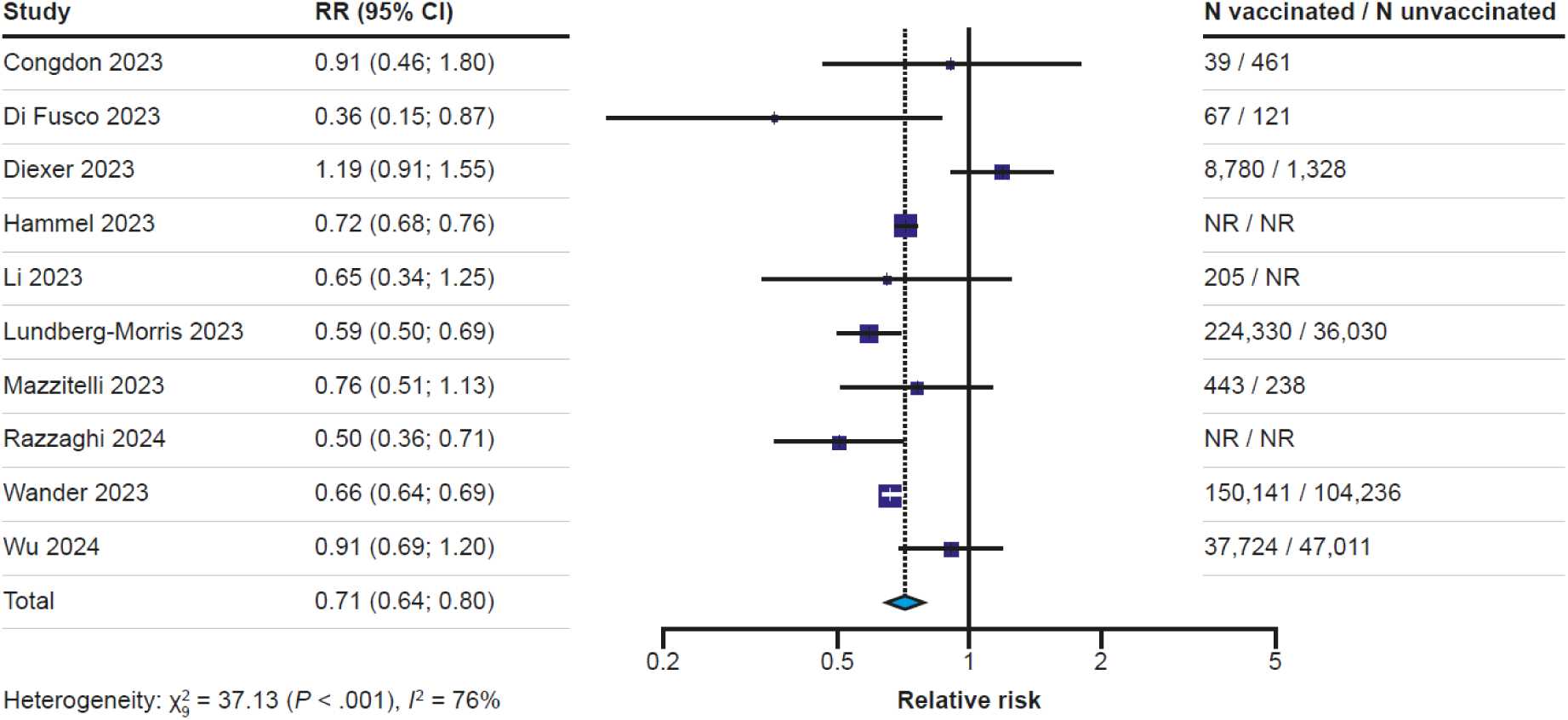
Forest plot for the effect of any vaccination (analysis B) on the risk of long COVID compared with no vaccination (random effects) CI, confidence interval; NR, not reported; RR, relative risk Two separate analyses were performed for “any vaccination” versus no vaccination to account for stratification of results in three studies by primary course vaccination and booster vaccination; analysis B (presented here) included the booster vaccination data for those stratified studies. In the forest plot, the squares and horizontal bars represent the relative risk and 95% CI, respectively, for each study. The dotted line and diamond represent the pooled relative risk and 95% CI, respectively, for all the studies included in the analysis.

The results of the Egger’s test of publication bias^36^ were non-significant for both analysis A and B, and funnel plots were generally symmetrical (**Supplementary Figures 1 and 2**), indicating no evidence of publication bias.

Sensitivity analyses indicated that the results of both analysis A and B were robust. Leave-one-out plots for both analyses did not result in loss of statistical significance of the pooled RR value (**Supplementary Figure 3 and 4**). Removal of a potential overlapping population between studies, children/adolescent-only studies, studies with a high hospitalization for acute illness rate, and pre-prints for both analyses did not result in loss of statistical significance of the pooled RR value (**Supplementary Tables 10 and 11**).

#### Impact of primary course vaccination prior to Omicron variant infection on long COVID compared with no vaccination

Three studies^10,41,44^ that reported risk of long COVID in individuals who had received the primary course of vaccines versus unvaccinated individuals were included in this analysis (Figure 4). Primary course vaccination was associated with a significantly lower risk of long COVID (*P*<0.0001) compared with unvaccinated individuals, with a pooled RR of 0.81 (95% CI, 0.79–0.83; *P*<0.0001). Statistical heterogeneity was low (I^2^=0%). However, sensitivity analyses could not be performed due to the small number of studies.

**Figure 4.**
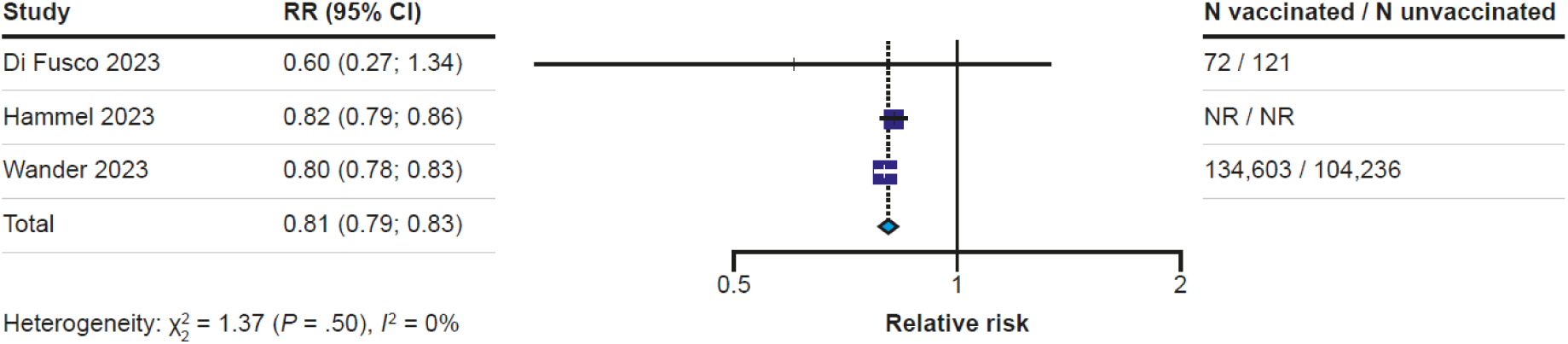
Forest plot for the effect of primary course vaccination on the risk of long COVID compared with no vaccination (random effects) CI, confidence interval; NR, not reported; RR, relative risk In the forest plot, the squares and horizontal bars represent the relative risk and 95% CI, respectively, for each study. The dotted line and diamond represent the pooled relative risk and 95% CI, respectively, for all the studies included in the analysis.

#### Impact of booster vaccination prior to Omicron variant infection on long COVID compared with no vaccination

Four studies^10,41,44,55^ that reported risk of long COVID in individuals who had received booster dose vaccination versus unvaccinated individuals were included in this analysis (Figure 5).

**Figure 5.**
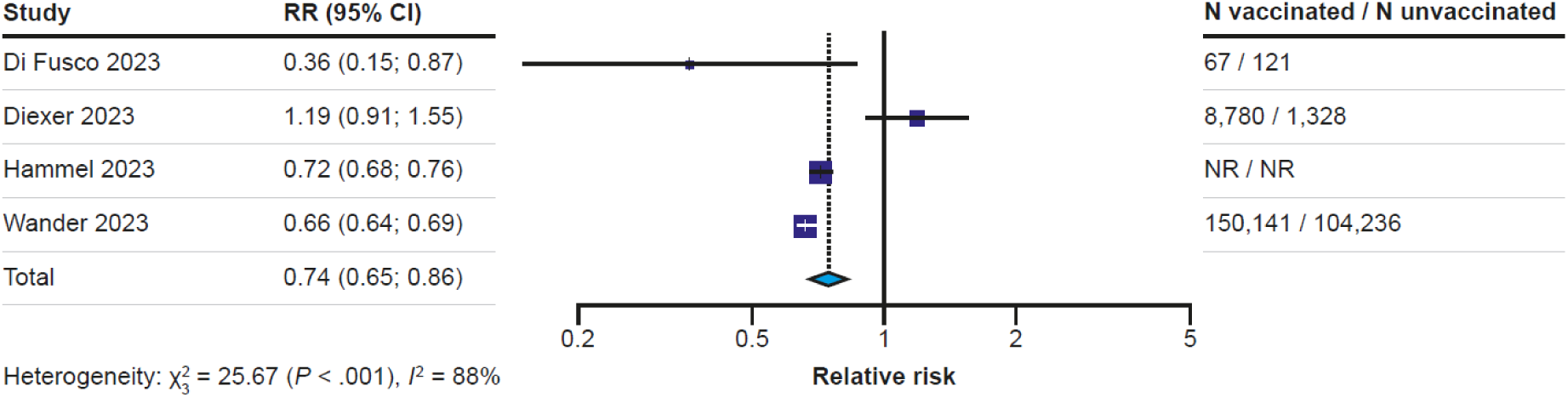
Forest plot for the effect of booster vaccination on the risk of long COVID compared with no vaccination. CI, confidence interval; NR, not reported; RR, relative risk In the forest plot, the squares and horizontal bars represent the relative risk and 95% CI, respectively, for each study. The dotted line and diamond represent the pooled relative risk and 95% CI, respectively, for all the studies included in the analysis.

Booster vaccination was associated with a significantly lowered risk of long COVID (*P*<0.0001) compared with individuals who were unvaccinated, with a pooled RR of 0.74 (95% CI, 0.65–0.86; *P*<0.0001). Between study heterogeneity was high (I^2=^88%). Sensitivity analyses found that removal of either Wander 2023^44^ or Hammel 2023^41^ resulted in loss of statistical significance of the pooled RR value (**Supplementary Figure 5**. Removal of a potential overlapping population between studies also resulted in loss of significance of the RR value (**Supplementary Table 12**).

#### Impact of booster vaccination on long COVID compared with primary course vaccination prior to Omicron variant infection

Three studies^10,55,61^ that reported risk of long COVID in individuals who had received booster dose vaccination versus individuals who had received the primary course only were included in this analysis (Figure 6). Booster vaccination was associated with a significantly lowered risk of long COVID compared with primary course vaccination only, with a pooled RR of 0.77 (95% CI, 0.65–0.92; *P*=0.004). Between study heterogeneity was low (I^2^=0%). Sensitivity analyses could not be performed due to the small number of studies.

**Figure 6.**
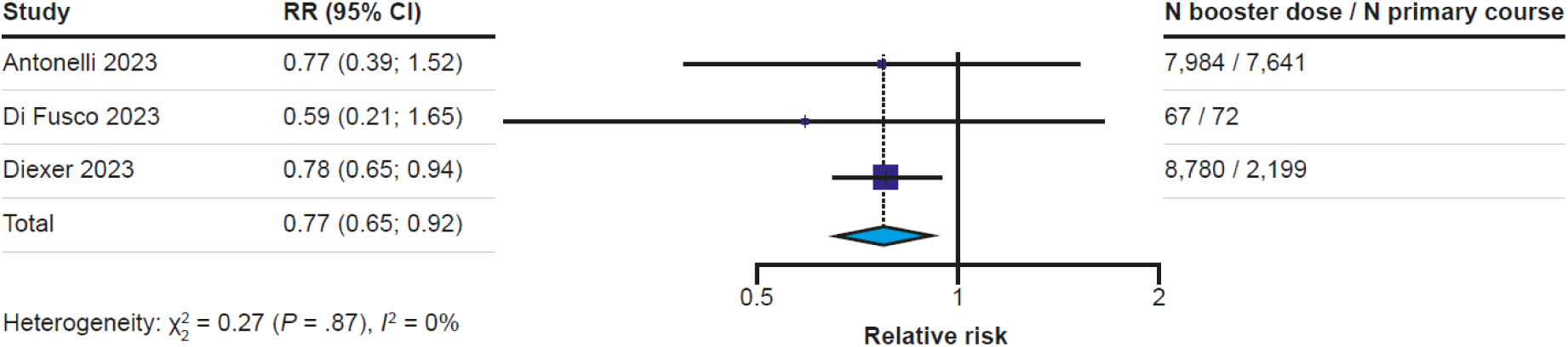
Forest plot for the effect of booster vaccination on the risk of long COVID compared with primary course vaccination. CI, confidence interval; RR, relative risk In the forest plot, the squares and horizontal bars represent the relative risk and 95% CI, respectively, for each study. The dotted line and diamond represent the pooled relative risk and 95% CI, respectively, for all the studies included in the analysis.

## Discussion

This SLR and meta-analysis investigated the impact of COVID-19 vaccination, including booster doses, on the risk of developing long COVID caused by Omicron variants infections. The risk of long COVID was 22% and 29% lower for participants receiving “any vaccination” compared with unvaccinated participants for analysis A and B, respectively (separate analyses to account for stratification of results in several studies), while 19% and 26% lower among those receiving primary course and booster vaccination, respectively, compared with unvaccinated participants. Among the vaccinated population, additional doses were associated with added protection against long COVID, demonstrated by a 23% risk reduction in the boosted versus primary course participants. This novel comparison between booster and primary course vaccination is critical, as it underscores the additional clinical advantages of seasonal COVID-19 vaccinations beyond the prevention of acute infections and severe disease.

Our analysis comparing primary vaccination versus no vaccination yielded results similar to those published in several pre-Omicron analyses^19,20,27^, which reported significantly lower pooled risks of long COVID development of 0.82 (95% CI, 0.74–0.91)^20^, 0.64 (95% CI, 0.45–0.92)^27^, and 0.54 (95% CI, 0.30–0.99)^19^. For studies that included Omicron variants, in combination with non-Omicron variants, a significantly lower pooled risk of long COVID was reported for primary course vaccination (0.65 [95% CI, 0.62–0.68])^23,67^; for primary course and booster vaccination (0.77 [95% CI, 0.75–0.79])^68^ and for booster doses only (0.31 [95% CI, 0.28–0.35])^23^, all versus no vaccination. A previous meta-analysis performed a sub-analysis of Omicron infections only and reported a pooled risk of 0.68 (95% CI, 0.54–0.86) for vaccinated versus unvaccinated participants, similar to the results presented here (RR 0.78 [95% CI, 0.71–0.85] and 0.71 [95% CI, 0.64–0.80] for analyses A and B, respectively).

In summary, our findings support recent evidence suggesting that primary course vaccination and, to a greater extent, booster vaccination prior to SARS-CoV-2 infection may reduce the risk of long COVID symptoms. Further, an SLR currently under peer-review has reported that existing data on COVID-19 vaccination in adults demonstrates that the effect of vaccination prior to infection on long COVID occurs regardless of the predominant variant in circulation^69^.

### Factors affecting long COVID risk

A recent cohort study of 441,583 veterans from the Department of Veterans Affairs Health Care System databases by Xie et al reported 5.23 fewer long COVID events per 100 persons at 1 year post infection during the Omicron era versus during the pre-Delta and Delta eras combined^4^. Further, it was estimated that 28% of the reduction in long COVID events was attributable to viral era-related changes, with the remaining attributable to the effect of vaccination^4^. These findings demonstrate that the incidence of long COVID in the Omicron era remains substantial.

For the data presented here, it cannot be determined whether the protective effect of vaccination against long COVID arises solely from a reduced risk of infection and severe disease (population-level) or whether this can occur irrespective of acute disease severity. Severe COVID-19, as determined by hospitalization or intensive care unit admittance, is among the strongest predictors for the risk of developing long COVID^70,71^. It follows that, by effectively reducing the incidence of severe COVID-19, vaccination may, in turn, prevent progression to long COVID, thus achieving a protective effect. The impact of repeated SARS-CoV-2 infection on risk of developing long COVID has not been fully elucidated. While some studies indicate that previous infection with SARS-CoV-2 may decrease the risk of long COVID after a second infection^55,72^, there is also evidence to suggest that reinfection leads to a cumulative increased risk of long COVID^73,74^.

There are a number of potential mechanisms by which vaccination may decrease the risk of long COVID. There is evidence to suggest long COVID could, in at least a subset of individuals, be attributed to persistence of SARS-CoV-2 RNA or protein, driving inflammatory, microbiome, and coagulation abnormalities^75,76^. Hence, a pre-existing vaccine-induced immune response could protect against the establishment of this so-called “viral reservoir” and, in turn, long COVID^75^. Findings have also suggested autoimmunity as a potential mechanism for long COVID pathogenesis^77^, with a recent study indicating that transfer of IgG antibodies from participants with long COVID can induce replicate symptoms in mice^78,79^ as well as hyperinflammation caused by atherosclerotic plaque formation in cases of cardiovascular complications^13,80^; these processes could be mitigated by vaccination. Vaccination may also reduce the risk of long COVID by protecting against acute and post-acute inflammation that may lead to organ dysfunction or damage^81^. Studies have found that sustained viral burden and viral RNA load in the acute phase of COVID-19 is associated with an increased risk of long COVID, though the underlying mechanisms have not been determined^82–85^. It follows that vaccination could increase the speed of viral clearance and in turn mitigate downstream pathways for long COVID pathology.

### Implications for practice

Much of the initial messaging around COVID-19 vaccination was focused on the prevention of severe illness, hospitalization, and death. As the pandemic evolves and these outcomes become less common, understanding the effectiveness of vaccination against long COVID could help provide evidence for both the individual patient-level benefit and public health value of ongoing seasonal vaccination. This may be especially true for young, healthy individuals, who might otherwise consider themselves to be at low risk for acute complications of COVID-19. Findings on vaccine effectiveness, and more so the added benefit of boosters, could therefore inform guidelines on preventative medicine or primary care and assist healthcare professionals in providing evidence-based advice to their patients regarding vaccination. For public health, increasing seasonal vaccine uptake is an important element of the recently outlined global research and policy response strategy required to address the multifaceted challenges posed by long COVID^13^. Our analysis on booster versus primary vaccination provides evidence supporting COVID-19 seasonal vaccination policies.

### Quality of evidence

To our knowledge, this is the first SLR and meta-analysis to compare the impact of booster vaccination with primary course vaccination for Omicron-only infections, thus providing an improved understanding of the importance of seasonal vaccination against newer variants. In addition, there was a low or medium risk of bias in all but one^64^ of the studies identified in the SLR, and sensitivity analyses did not negate the statistical significance of the RR values, except in booster dose versus unvaccinated analyses.

It is important to note several limitations of this review. Only English-language studies were included, which may have limited the selection of data. Further, the statistical heterogeneity estimate (I^2^) provided for the analyses should be interpreted with caution due to the limited number of studies^86^. In addition, the analyses do not explore the effect of infection prevention since all participants had been infected with SARS-CoV-2 and presented with symptoms; therefore, the risk reductions described here are conservative. There were limited subgroup analyses in the identified studies, meaning that sub-analyses could not be performed for sex, vaccine type, comorbidities, or acute infection severity. In addition, there was significant variation in the definition of long COVID across studies^49,56,87^; the lack of consensus on the definition of long COVID is an ongoing issue^88^. The follow-up periods of the included studies varied from 4 weeks to 6 months depending on the definition of long COVID used and the aim of the study. Further, there were variations in vaccination status definitions across studies. For example, in one study, unvaccinated participants included those who received a monovalent dose ≥12 months before enrolment^62^. There is evidence that the effects of vaccination have not fully waned at 12 months after vaccination^89^, so participants grouped using this definition may not be comparable to those who have never been vaccinated.

Another major limitation is the lack of randomization in the studies included in the meta-analysis and although studies adjusted for confounders, methods were not consistent across studies. Thus, the reduced risk of long COVID among patients who were vaccinated (or received a booster dose) may be due to systematic differences in the baseline characteristics of both groups.

### Implications for further research

As discussed, our meta-analysis is limited by varying definitions and heterogeneity in subgroup analyses. As such, an individual patient data meta-analysis could help address some of these drawbacks. In addition, there are still many unresolved research questions regarding long COVID in the current-Omicron era. For instance, the potential impact of seasonal vaccination on the risk of developing long COVID is unclear in people with hybrid immunity (repeat infection and seasonal vaccinations) compared with primary course vaccinated individuals. Given the success of global vaccination efforts and estimated seroprevalence, it will be difficult to recruit unvaccinated individuals or people who have not been exposed to SARS-CoV-2 infection, as a baseline comparator for future studies. Another important factor to investigate is the impact of combined flu and COVID-19 vaccination on the rates of long COVID, particularly as COVID-19 vaccinations have moved to a seasonal roll out (e.g. UK 2024 COVID-19 autumn vaccination^90^). Further, it is important to recognize that long COVID does not exist in isolation, and thereby, gain understanding of its intersection with broader healthcare challenges. A retrospective analysis of UK health records investigated these so called “compound pressures” and observed that flu vaccination and the preventative treatment for cardiovascular disease were associated with a reduced risk of hospitalisation due to long COVID^91^.

Previous studies have reported on the considerable healthcare costs associated with long COVID in the Omicron era^92–94^; however, it is important to investigate the cost-effectiveness of COVID-19 vaccines and update cost analyses, in relation to direct and indirect cost to healthcare systems and society, and as SARS-CoV-2 continues to evolve, to help inform government/public health policy.

## Conclusion

Both primary course and, to a greater extent, booster COVID-19 vaccination, were significantly associated with a reduced risk of developing long COVID following Omicron variant infection. This cumulative protective effect against long COVID from additional booster doses is likely due to improved protection against severe acute COVID-19, known to be a key risk factor for the development of long COVID. Our study highlights the importance of continued surveillance of SARS-CoV-2 variants in parallel to ongoing assessment of the effectiveness of current seasonal vaccines in preventing long COVID. These findings can inform evidence-based public health messaging around the individual and public health benefits of seasonal vaccination programs.

## Acknowledgements

The authors would like to thank Triantafyllos Pliakas (BioNTech SE and Impact Epilysis) for their contribution to the manuscript.

Statistical support, including the design and running of the meta-analyses, was provided by Medha Shrivastava, MSc (Maverex Ltd). Kate Misso, MSc/MCLIP (Maverex Ltd), designed and performed the electronic searches in this systematic review.

Medical writing support, including assisting authors with the development of the outline and initial draft, and incorporation of comments, was provided by Jasmine Besilum, MSc, and editorial support was provided by Sarah Christopher, of Paragon (a division of Prime, Knutsford, UK), supported by BioNTech SE according to Good Publication Practice guidelines (<u>Link</u>). The sponsor was involved in the study design and collection, analysis, and interpretation of data, as well as data checking of information provided in the manuscript. However, ultimate responsibility for opinions, conclusions, and data interpretation lies with the authors.

## Data availability

The datasets generated during and/or analyzed during the current study are available from the corresponding author on reasonable request.

## Author contributions

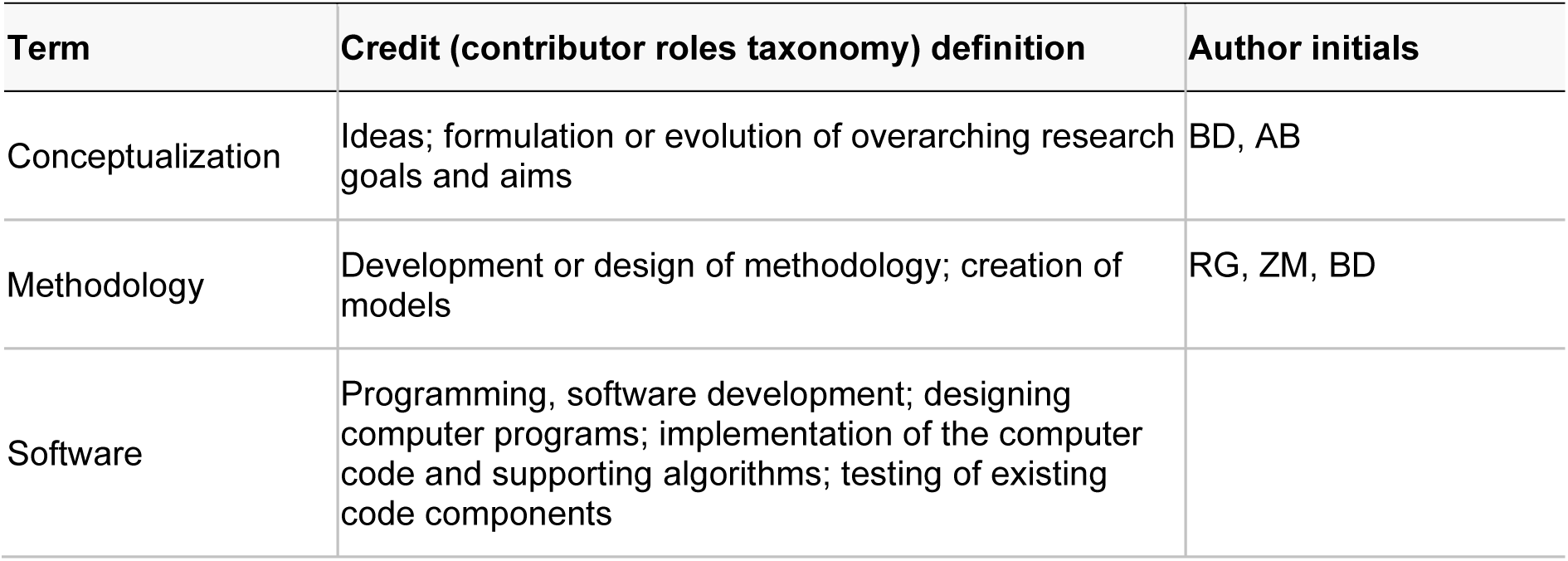
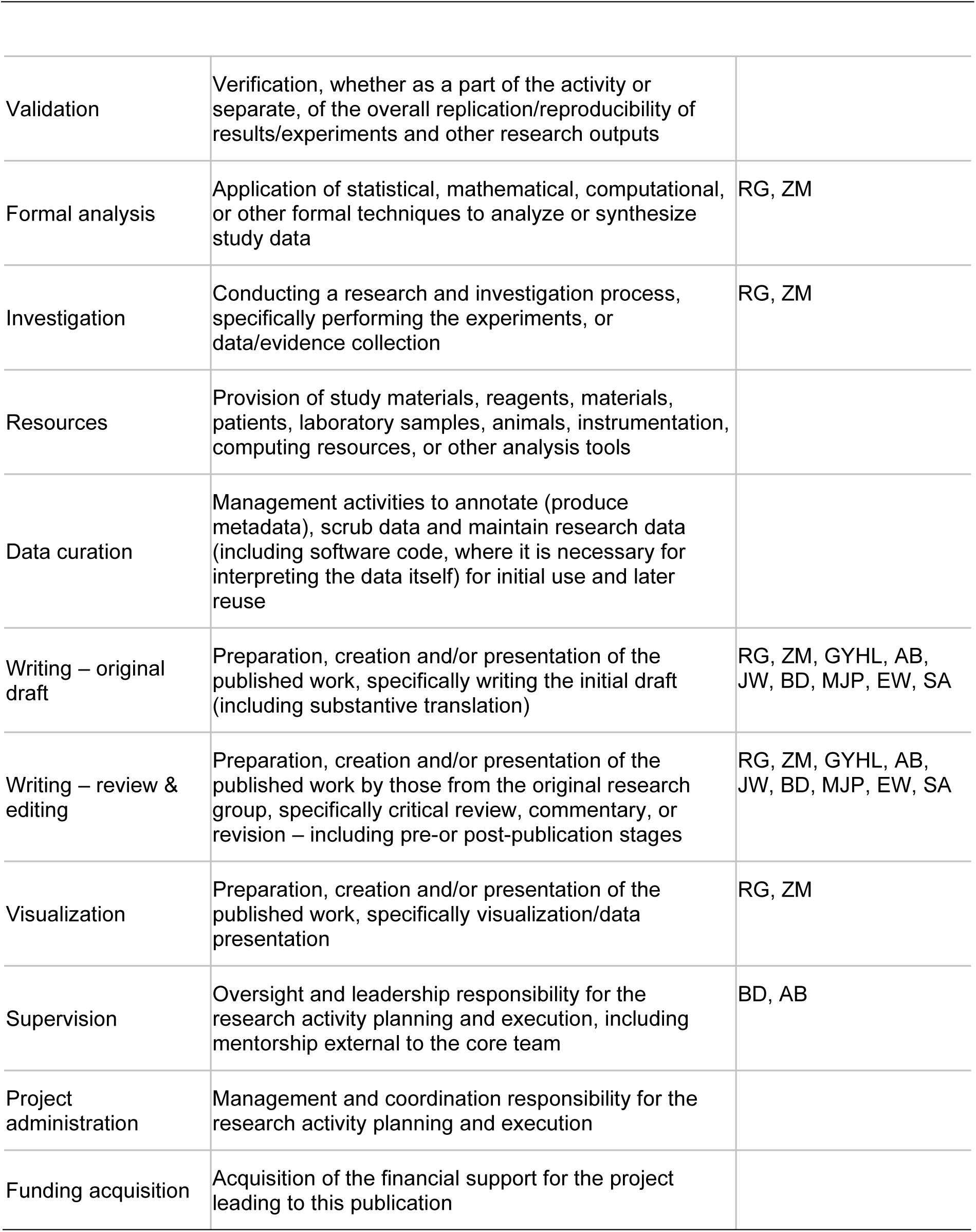

## References

1 World Health Organization. Post COVID-19 condition (Long COVID), <https://www.who.int/europe/news-room/fact-sheets/item/post-covid-19-condition> (2022).

2 Greenhalgh, T., Sivan, M., Perlowski, A. & Nikolich, J. Z. Long COVID: a clinical update. Lancet 404, 707–724 (2024). 10.1016/S0140-6736(24)01136-X

3 Fang, Z., Ahrnsbrak, R. & Rekito, A. Evidence mounts that about 7% of US adults have had long COVID. JAMA 332, 5–6 (2024). 10.1001/jama.2024.11370

4 Xie, Y., Choi, T. & Al-Aly, Z. Postacute sequelae of SARS-CoV-2 infection in the pre-Delta, Delta, and Omicron eras. N Engl J Med 391, 515–525 (2024). 10.1056/NEJMoa2403211

5 Davis, H. E., McCorkell, L., Vogel, J. M. & Topol, E. J. Author Correction: Long COVID: major findings, mechanisms and recommendations. Nat Rev Microbiol 21, 408 (2023). 10.1038/s41579-023-00896-0

6 Bowe, B., Xie, Y. & Al-Aly, Z. Postacute sequelae of COVID-19 at 2 years. Nat Med 29, 2347–2357 (2023). 10.1038/s41591-023-02521-2

7 National Academy of Sciences. A long COVID definition. (2024).

8 National institute of Health and Care Excellence (NICE). COVID-19 rapid guideline: managing the long-term effects of COVID-19, <https://www.nice.org.uk/guidance/ng188> (2024).

9 Sanchez-Ramirez, D. C., Normand, K., Zhaoyun, Y. & Torres-Castro, R. Long-term impact of COVID-19: a systematic review of the literature and meta-analysis. Biomedicines 9, 900 (2021).

10 Di Fusco, M. et al. Impact of COVID-19 and effects of booster vaccination with BNT162b2 on six-month long COVID symptoms, quality of life, work productivity and activity impairment during Omicron. J Patient Rep Outcomes 7, 77 (2023). 10.1186/s41687-023-00616-5

11 Lambert, N. et al. The other COVID-19 survivors: timing, duration, and health impact of post-acute sequelae of SARS-CoV-2 infection. J Clin Nurs 33, 76–88 (2022). 10.1111/jocn.16541

12 Wolff Sagy, Y., Feldhamer, I., Brammli-Greenberg, S. & Lavie, G. Estimating the economic burden of long-Covid: the additive cost of healthcare utilisation among COVID-19 recoverees in Israel. BMJ Glob Health 8 (2023). 10.1136/bmjgh-2023-012588

13 Al-Aly, Z. et al. Long COVID science, research and policy. Nat Med 30, 2148–2164 (2024). 10.1038/s41591-024-03173-6

14 Byambasuren, O., Stehlik, P., Clark, J., Alcorn, K. & Glasziou, P. Effect of covid-19 vaccination on long covid: systematic review. BMJ Med 2, e000385 (2023). 10.1136/bmjmed-2022-000385

15 Català, M. et al. The effectiveness of COVID-19 vaccines to prevent long COVID symptoms: staggered cohort study of data from the UK, Spain, and Estonia. Lancet Respir Med 12, 225–236 (2024). 10.1016/s2213-2600(23)00414-9

16 Malden, D. E. et al. Post-COVID conditions following COVID-19 vaccination: a retrospective matched cohort study of patients with SARS-CoV-2 infection. Nat Commun 15, 4101 (2024). 10.1038/s41467-024-48022-9

17 De Domenico, M. Prevalence of long COVID decreases for increasing COVID-19 vaccine uptake. PLOS Glob Public Health 3, e0001917 (2023). 10.1371/journal.pgph.0001917

18 Brannock, M. D. et al. Long COVID risk and pre-COVID vaccination in an EHR-based cohort study from the RECOVER program. Nat Commun 14, 2914 (2023). 10.1038/s41467-023-38388-7

19 Ceban, F. et al. COVID-19 vaccination for the prevention and treatment of long COVID: a systematic review and meta-analysis. Brain Behav Immun 111, 211–229 (2023). 10.1016/j.bbi.2023.03.022

20 Gao, P., Liu, J. & Liu, M. Effect of COVID-19 vaccines on reducing the risk of long COVID in the real world: a systematic review and meta-analysis. Int J Environ Res Public Health 19, 12422 (2022). 10.3390/ijerph191912422

21 Trinh, N. T. et al. Effectiveness of COVID-19 vaccines to prevent long COVID: data from Norway. Lancet Respir Med 12, e33–e34 (2024). 10.1016/s2213-2600(24)00082-1

22 Lundberg-Morris, L. et al. Covid-19 vaccine effectiveness against post-covid-19 condition among 589 722 individuals in Sweden: population based cohort study. BMJ 383, e076990 (2023). 10.1136/bmj-2023-076990

23 Marra, A. R. et al. The effectiveness of COVID-19 vaccine in the prevention of post-COVID conditions: a systematic literature review and meta-analysis of the latest research. Antimicrob Steward Healthc Epidemiol 3, e168 (2023). 10.1017/ash.2023.447

24 World Health Organization. Classification of Omicron (B.1.1.529): SARS-CoV-2 variant of concern, <https://www.who.int/news/item/26-11-2021-classification-of-omicron-(b.1.1.529)-sars-cov-2-variant-of-concern> (2021).

25 Hattab, D., Amer, M. F. A., Al-Alami, Z. M. & Bakhtiar, A. SARS-CoV-2 journey: from alpha variant to omicron and its sub-variants. Infection 52, 767–786 (2024). 10.1007/s15010-024-02223-y

26 World Health Organization. COVID-19 epidemiological update – 13 August 2024. (2024). <https://www.who.int/publications/m/item/covid-19-epidemiological-update-edition-170>.

27 Watanabe, A., Iwagami, M., Yasuhara, J., Takagi, H. & Kuno, T. Protective effect of COVID-19 vaccination against long COVID syndrome: a systematic review and meta-analysis. Vaccine 41, 1783–1790 (2023). 10.1016/j.vaccine.2023.02.008

28 Gutfreund, M. C. et al. The effectiveness of the COVID-19 vaccines in the prevention of post-COVID conditions in children and adolescents: a systematic literature review and meta-analysis. Antimicrobial Steward Healthc Epidemiol 4, e54 (2024). 10.1017/ash.2024.42

29 Page, M. J. et al. The PRISMA 2020 statement: an updated guideline for reporting systematic reviews. Int J Surg 88, 105906 (2021). 10.1016/j.ijsu.2021.105906

30 Centers for Disease Control and Prevention. Post-COVID conditions: information for healthcare providers, <https://archive.cdc.gov/#/details?q=long%20covid&start=0&rows=10&url=https://www.cdc.gov/coronavirus/2019-ncov/hcp/clinical-care/post-covid-conditions.html> (2022).

31 Wells G, S. B., O’Connell D, Peterson J, Welch V, Losos M. The Newcastle-Ottawa Scale (NOS) for assessing the quality of nonrandomised studies in meta-analyses, <https://www.ohri.ca/programs/clinical_epidemiology/oxford.asp> (2013).

32 JBI. Critical appraisal tools, <https://jbi.global/critical-appraisal-tools> (2024).

33 Greenland, S. Quantitative methods in the review of epidemiologic literature. Epidemiol Rev 9, 1–30 (1987). 10.1093/oxfordjournals.epirev.a036298

34 Hightower, A. W., Orenstein, W. A. & Martin, S. M. Recommendations for the use of Taylor series confidence intervals for estimates of vaccine efficacy. Bull World Health Organ 66, 99–105 (1988).

35 DerSimonian, R. & Laird, N. Meta-analysis in clinical trials. Control Clin Trials 7, 177–188 (1986). 10.1016/0197-2456(86)90046-2

36 Higgins JPT, T. J., Chandler J, Li T, Page MJ, Welch V, eds. Cochrane Handbook for Systematic Reviews of Interventions [Internet]. Version 6.4 Cochrane, updated August 2023. [cited 2024 May 28]. Available from: https://training.cochrane.org/handbook/current.

37 Egger, M., Davey Smith, G., Schneider, M. & Minder, C. Bias in meta-analysis detected by a simple, graphical test. BMJ 315, 629–634 (1997). 10.1136/bmj.315.7109.629

38 Domenech-Montoliu, S. et al. Long COVID prevalence and the impact of the third SARS-CoV-2 vaccine dose: A cross-sectional analysis from the third follow-ip of the Borriana cohort, Valencia, Spain (2020-2022). Vaccines (Basel) 11 (2023). 10.3390/vaccines11101590

39 Mazzitelli, M. et al. Risk of hospitalization and sequelae in patients with COVID-19 treated with 3-day early remdesivir vs. controls in the vaccine and Omicron era: A real-life cohort study. J Med Virol 95, e28660 (2023). 10.1002/jmv.28660

40 Razzaghi, H. et al. Vaccine effectiveness against long COVID in children. Pediatrics 153 (2024). 10.1542/peds.2023-064446

41 Hammel, I. S., Tosi, D. M., Tang, F., Pott, H. & Ruiz, J. G. Frailty as a risk factor for post-acute sequelae of COVID-19 among US veterans during the Delta and Omicron waves. J Am Geriatr Soc 71, 3826–3835 (2023). 10.1111/jgs.18584

42 Congdon, S. et al. Nirmatrelvir/ritonavir and risk of long COVID symptoms: a retrospective cohort study. Sci Rep 13, 19688 (2023). 10.1038/s41598-023-46912-4

43 Hedberg, P. & Naucler, P. Post-COVID-19 condition after SARS-CoV-2 infections during the omicron surge vs the delta, alpha, and wild type periods in Stockholm, Sweden. J Infect Dis 229, 133–136 (2024). 10.1093/infdis/jiad382

44 Wander, P. L. et al. Rates of ICD-10 Code U09.9 documentation and clinical characteristics of VA patients with post-COVID-19 condition. JAMA Netw Open 6, e2346783 (2023). 10.1001/jamanetworkopen.2023.46783

45. Wu, Q., et al. Real-world effectiveness and causal mediation study of BNT162b2 on long COVID risks in children and adolescents. medRxiv (2024).

46 Cortellini, A. et al. SARS-CoV-2 omicron (B.1.1.529)-related COVID-19 sequelae in vaccinated and unvaccinated patients with cancer: results from the OnCovid registry. Lancet Oncol 24, 335–346 (2023). 10.1016/S1470-2045(23)00056-6

47 Sun, X., et al. Postacute sequelae SARS-CoV-2 infection by vaccination status: A six-month latent class analysis. medRxiv (2023).

48 Ballouz, T. et al. Post COVID-19 condition after Wildtype, Delta, and Omicron SARS-CoV-2 infection and prior vaccination: Pooled analysis of two population-based cohorts. PLoS One 18, e0281429 (2023). 10.1371/journal.pone.0281429

49 Gallant, M., Mercier, K., Rioux-Perreault, C., Lemaire-Paquette, S. & Piché, A. Prevalence of persistent symptoms at least 1 month after SARS-CoV-2 Omicron infection in adults. J Assoc Med Microbiol Infect Dis Can 8, 57–63 (2023). 10.3138/jammi-2022-0026

50 Herting, A. et al. Clinical outcomes of SARS-CoV-2 breakthrough infections in liver transplant recipients during the omicron wave. Viruses 15 (2023). 10.3390/v15020297

51 Kahlert, C. R. et al. Post-acute sequelae after severe acute respiratory syndrome coronavirus 2 infection by viral variant and vaccination status: A multicenter cross-sectional study. Clin Infect Dis 77, 194–202 (2023). 10.1093/cid/ciad143

52 Reme, B. A., Gjesvik, J. & Magnusson, K. Predictors of the post-COVID condition following mild SARS-CoV-2 infection. Nat Commun 14, 5839 (2023). 10.1038/s41467-023-41541-x

53 Spiliopoulos, L. et al. Post-acute symptoms 4 months after SARS-CoV-2 infection during the Omicron period: a nationwide Danish questionnaire study. Am J Epidemiol (2023). 10.1093/aje/kwad225

54 Woldegiorgis, M., Cadby, G., Ngeh, S. & Korda, R. Long COVID in a highly vaccinated population infected during a SARS-CoV-2 Omicron wave – Australia, 2022. medRxiv (2023).

55 Diexer, S. et al. Association between virus variants, vaccination, previous infections, and post-COVID-19 risk. Int J Infect Dis 136, 14–21 (2023). 10.1016/j.ijid.2023.08.019

56 de Bruijn, S. et al. Lower prevalence of post-Covid-19 condition following Omicron SARS-CoV-2 infection. Heliyon 10, e28941 (2024). 10.1016/j.heliyon.2024.e28941

57 Thaweethai, T. et al. Development of a definition of postacute sequelae of SARS-CoV-2 infection. JAMA 329, 1934–1946 (2023). 10.1001/jama.2023.8823

58 Nehme, M. et al. Prevalence of post-coronavirus disease condition 12 weeks after omicron infection compared with negative controls and association with vaccination status. Clin Infect Dis 76, 1567–1575 (2023). 10.1093/cid/ciac947

59 Brown, M. et al. Ongoing symptoms and functional impairment 12 weeks after testing positive for SARS-CoV-2 or influenza in Australia: an observational cohort study. BMJ Public Health 1, e000060 (2023).

60 Richard, S. A. et al. Decreased self-reported physical fitness following SARS-CoV-2 infection and the impact of vaccine boosters in a cohort study. Open Forum Infect Dis 10, ofad579 (2023). 10.1093/ofid/ofad579

61 Antonelli, M. et al. SARS-CoV-2 infection following booster vaccination: Illness and symptom profile in a prospective, observational community-based case-control study. J Infect 87, 506–515 (2023). 10.1016/j.jinf.2023.08.009

62 Di Fusco, M. et al. Effectiveness of BNT162b2 BA.4/5 bivalent COVID-19 vaccine against long COVID symptoms: A US nationwide study. Vaccines (Basel) 12, 183 (2024). 10.3390/vaccines12020183

63 Li, J., Nadua, K., Chong, C. Y. & Yung, C. F. Long COVID prevalence, risk factors and impact of vaccination in the paediatric population: a survey study in Singapore. Ann Acad Med Singap 52, 522–532 (2023). 10.47102/annals-acadmedsg.2023238

64 AlBahrani, S. et al. Self-reported long COVID-19 symptoms are rare among vaccinated healthcare workers. J Infect Public Health 16, 1276–1280 (2023). 10.1016/j.jiph.2023.05.037

65 Mikolajczyk R, D. S., Klee B, Pfrommer L, Purschke O, Fricke J, et al. Risk of Post-COVID Condition Under Hybrid Immunity - Results from the German National Cohort (NAKO). SSRN (2023).

66 Huh, K. et al. Vaccination and the risk of post-acute sequelae after COVID-19 in the Omicron-predominant period. Clin Microbiol Infect 30, 666–673 (2024). 10.1016/j.cmi.2024.01.028

67 Marra, A. R. et al. The effectiveness of coronavirus disease 2019 (COVID-19) vaccine in the prevention of post-COVID-19 conditions: A systematic literature review and meta-analysis. Antimicrob Steward Healthc Epidemiol 2, e192 (2022). 10.1017/ash.2022.336

68 Man, M. A. et al. Impact of pre-infection COVID-19 vaccination on the incidence and severity of post-COVID syndrome: a systematic review and meta-analysis. Vaccines (Basel) 12, 189 (2024). 10.3390/vaccines12020189

69 Rudolph, A. E. et al. Factors affecting the impact of COVID-19 vaccination on post COVID-19 conditions among adults: A Systematic Literature Review. medRxiv, 2024.2010.2002.24314603 (2024). 10.1101/2024.10.02.24314603

70 Al-Aly, Z., Bowe, B. & Xie, Y. Long COVID after breakthrough SARS-CoV-2 infection. Nat Med 28, 1461–1467 (2022). 10.1038/s41591-022-01840-0

71 Tsampasian, V. et al. Risk factors associated with post-COVID-19 condition: a systematic review and meta-analysis. JAMA Intern Med 183, 566–580 (2023). 10.1001/jamainternmed.2023.0750

72 Mikolajczyk, R. et al. Likelihood of post-COVID condition in people with hybrid immunity; data from the German National Cohort (NAKO). J Infect 89, 106206 (2024). 10.1016/j.jinf.2024.106206

73 Bowe, B., Xie, Y. & Al-Aly, Z. Acute and postacute sequelae associated with SARS-CoV-2 reinfection. Nat Med 28, 2398–2405 (2022). 10.1038/s41591-022-02051-3

74 Barboza, A. Long COVID in a Multicentric Brazilian Cohort: Acute Phase Symptom Burden and Second Booster Dose Vaccination Effects. SSRN (2023). 10.2139/ssrn.4571726

75 Proal, A. D. et al. SARS-CoV-2 reservoir in post-acute sequelae of COVID-19 (PASC). Nat Immunol 24, 1616–1627 (2023). 10.1038/s41590-023-01601-2

76 Altmann, D. M., Whettlock, E. M., Liu, S., Arachchillage, D. J. & Boyton, R. J. The immunology of long COVID. Nat Rev Immunol 23, 618–634 (2023). 10.1038/s41577-023-00904-7

77 Peluso, M. J. & Deeks, S. G. Mechanisms of long COVID and the path toward therapeutics. Cell 187, 5500–5529 (2024). 10.1016/j.cell.2024.07.054

78 Chen, H. Transfer of IgG from Long COVID patients induces symptomology in mice. bioRxiv (2024). 10.1101/2024.05.30.596590

79 Wong, C. What causes long COVID? Case builds for rogue antibodies. Nature 630, 798–799 (2024). 10.1038/d41586-024-02010-7

80 Eberhardt, N. et al. SARS-CoV-2 infection triggers pro-atherogenic inflammatory responses in human coronary vessels. Nat Cardiovasc Res 2, 899–916 (2023). 10.1038/s44161-023-00336-5

81 Grady, C. B. et al. Impact of COVID-19 vaccination on symptoms and immune phenotypes in vaccine-naïve individuals with long COVID. medRxiv (2024). 10.1101/2024.01.11.24300929

82 Herbert, C. et al. Relationship between acute SARS-CoV-2 viral clearance with long COVID symptoms: a cohort study. medRxiv (2024). 10.1101/2024.07.04.24309953

83 Antar, A. A. R. et al. Long COVID brain fog and muscle pain are associated with longer time to clearance of SARS-CoV-2 RNA from the upper respiratory tract during acute infection. Front Immunol 14, 1147549 (2023). 10.3389/fimmu.2023.1147549

84 Ozonoff, A. et al. Features of acute COVID-19 associated with post-acute sequelae of SARS-CoV-2 phenotypes: results from the IMPACC study. Nat Commun 15, 216 (2024). 10.1038/s41467-023-44090-5

85 Lu, S. et al. Early biological markers of post-acute sequelae of SARS-CoV-2 infection. Nat Commun 15, 7466 (2024). 10.1038/s41467-024-51893-7

86 von Hippel, P. T. The heterogeneity statistic I(2) can be biased in small meta-analyses. BMC Med Res Methodol 15, 35 (2015). 10.1186/s12874-015-0024-z

87 Di Fusco, M. et al. Impact of COVID-19 and effects of BNT162b2 on patient-reported outcomes: quality of life, symptoms, and work productivity among US adult outpatients. J Patient Rep Outcomes 6, 123 (2022). 10.1186/s41687-022-00528-w

88 Chaichana, U. et al. Definition of post-COVID-19 condition among published research studies. JAMA Netw Open 6, e235856 (2023). 10.1001/jamanetworkopen.2023.5856

89 Wisnivesky, J. P. et al. Long-term persistence of neutralizing antibodies to SARS-CoV-2 following infection. J Gen Intern Med 36, 3289–3291 (2021). 10.1007/s11606-021-07057-0

90 UK Health Security Agency. A guide to the COVID-19 autumn vaccination, <https://www.gov.uk/government/publications/covid-19-vaccination-autumn-booster-resources/a-guide-to-the-covid-19-autumn-vaccination#:~:text=The%20Coronavirus%20(COVID%2D19),protect%20them%20ahead%20of%20winter.> (2024).

91 Dashtban, A. et al. Vaccinations, cardiovascular drugs, hospitalization, and mortality in COVID-19 and Long COVID. Int J Infect Dis 146, 107155 (2024). 10.1016/j.ijid.2024.107155

92 Walter, N., et al. A Comprehensive Report of German Nationwide Inpatient Data on the Post-COVID-19 Syndrome Including Annual Direct Healthcare Costs. Viruses 14 (2022). 10.3390/v14122600

93 Pike, J., Kompaniyets, L., Lindley, M. C., Saydah, S. & Miller, G. Direct Medical Costs Associated With Post-COVID-19 Conditions Among Privately Insured Children and Adults. Prev Chronic Dis 20, E06 (2023). 10.5888/pcd20.220292

94 Gandjour, A. Long COVID: Costs for the German economy and health care and pension system. BMC Health Serv Res 23, 641 (2023). 10.1186/s12913-023-09601-6

95 Zhou, S. & Shen, C. Avoiding definitive conclusions in meta-analysis of heterogeneous studies with small sample sizes. JAMA Otolaryngol Head Neck Surg 148, 1003–1004 (2022). 10.1001/jamaoto.2022.2847

