## Supplementary material for "The impact of vaccination on preventing long COVID in the Omicron era: a systematic review and meta-analysis"

### Supplementary materials

#### 1. Supplementary Methods

##### 1.1 Search strategy

##### Databases searched:

- Embase (Ovid): 2022-2024/03/01
- Medline ALL (Ovid): 2022-2024/03/01
- PubMed (NLM): 2022-2024/03/01
- LILACS (www): 2022-2024/03/01
- Europe PMC, includes BioRxiv & medRxiv (www): 2022-2024/03/01
- WHO Covid-19 database† (www): 2022-2024/01/25
- Cochrane COVID Register‡ (www): 2022-2024/02/02

†WHO COVID-19 database ceased on June 2023, therefore update searches were not necessary; ‡Cochrane COVID-9 Study Register ceased at the end of January 2024, therefore update searches were not necessary

**Embase (Ovid): 2022-2024/01/08**
**Searched 9.1.24**

1 immunization/ or active immunization/ or mass immunization/ or vaccination/ or vaccination coverage/ or vaccine failure/ 324831
2 revaccination/ 3068
3 (immunis$ or immuniz$ or immunity or vaccin$ or jab or jabs or shot or shots or booster or boosters or revaccin$ or unvaccin$).ti,ab,ot,kf,kw. 885844
4 or/1-3 940180
5 (PASC or "chronic covid syndrome$").ti,ab,ot,kf,kw. 1139
6 ("post acute" adj2 sequela$ adj2 (covid or coronavirus or coronovirus or "corona virus" or COV)).ti,ab,ot,kw. 476
7 (("long$ term$" or longterm$ or "long$ haul$" or longhaul$ or "long$ tail$" or longtail$ or longduration$ or "long duration$" or longlast$ or "long last$" or longstanding$ or "long standing$" or "medium$ term$" or mediumterm$ or "late effect$" or recurren$ or prolong$ or post-viral$ or chronic$ or postacute or "post acute" or persistent$) adj3 (covid$ or coronavirus$ or corona$ virus$ or coronovirus$ or corono$ virus$ or coronavirinae$ or corona$ virinae$ or Cov or "2019-nCoV$" or 2019nCoV$ or "19-nCoV$" or 19nCoV$ or nCoV2019$ or "nCoV-2019$" or nCoV19$ or "nCoV-19$" or "HCoV-19$" or HCoV19$ or "HCoV-2019$" or HCoV2019$ or "2019 novel$" or Ncov$ or "n-cov" or "SARS-CoV-2$" or "SARSCoV-2$" or "SARSCoV2$" or "SARS-CoV2$" or SARSCov19$ or "SARS-Cov19$" or "SARSCov-19$" or "SARS-Cov-19$" or SARSCov2019$ or "SARS-Cov2019$" or "SARSCov-2019$" or "SARS-Cov-2019$" or SARS2$ or "SARS-2$" or SARScoronavirus2$ or "SARS-coronavirus-2$" or "SARScoronavirus 2$" or "SARS coronavirus2$" or

SARScoronovirus2$ or "SARS-coronovirus-2$" or "SARScoronovirus 2$" or "SARS coronovirus2$" or "severe acute respiratory syndrome$")).ti,ab,ot,kw. 10596

8 (longcovid$ or long covid$ or longcoronavirus$ or longcorona$ virus$ or long coronavirus$ or long corona$ virus$ or longcoronovirus$ or longcorono$ virus$ or long coronovirus$ or long corono$ virus$ or longcoronavirinae$ or longcorona$ virinae$ or long coronavirinae$ or long corona$ virinae$ or longCov or long Cov or longsars$ or long sars$ or "long severe acute respiratory syndrome$" or longncov$ or long ncov$ or longhcov$ or long hcov$ or "post-acute covid" or "post-acute corona$" or "post-acute corono$" or "post-acute cov" or "post-acute ncov" or "post-acute sars" or "post-acute severe respiratory syndrome$").ti,ab,ot,kw. 6458

9 ((long$ or endur$ or legacy$ or slow$ or gradual$ or protract$ or lengthy$ or chronic$ or persist$ or relaps$ or remit$ or remission$ or residual$ or delay$ or prolong$ or extend$ or linger$ or permanent$ or fluctuat$ or sequela$ or multisystem$ or "multi system$" or nonrecover$ or "non recover$" or subacute$ or "sub acute$" or lasting$ or continuous$ or continual$ or continuing$ or postacute$ or "post acute$" or postdischarg$ or "post discharg$" or postinfect$ or "post infect$" or postviral$ or "post viral$" or postvirus$ or "post virus$" or "late effect$") adj2 (covid$ or coronavirus$ or corona$ virus$ or coronovirus$ or corono$ virus$ or coronavirinae$ or corona$ virinae$ or Cov or "2019-nCoV$" or 2019nCoV$ or "19-nCoV$" or 19nCoV$ or nCoV2019$ or "nCoV-2019$" or nCoV19$ or "nCoV-19$" or "HCoV-19$" or HCoV19$ or "HCoV-2019$" or HCoV2019$ or "2019 novel$" or Ncov$ or "n-cov" or "SARS-CoV-2$" or "SARSCoV-2$" or "SARSCoV2$" or "SARS-CoV2$" or SARSCov19$ or "SARS-Cov19$" or "SARSCov-19$" or "SARS-Cov-19$" or SARSCov2019$ or "SARS-Cov2019$" or "SARSCov-2019$" or "SARS-Cov-2019$" or SARS2$ or "SARS-2$" or SARScoronavirus2$ or "SARS-coronavirus-2$" or "SARScoronavirus 2$" or "SARS coronavirus2$" or SARScoronovirus2$ or "SARS-coronovirus-2$" or "SARScoronovirus 2$" or "SARS coronovirus2$" or "severe acute respiratory syndrome$")).ti,ab,ot,kw. 17052

10 (("long$ term$" or longterm$ or "long$ haul$" or longhaul$ or "long$ tail$" or longtail$ or longduration$ or "long duration$" or longlast$ or "long last$" or longstanding$ or "long standing$" or "medium$ term$" or mediumterm$ or "post-virus$" or "post-viral") adj3 (covid$ or coronavirus$ or corona$ virus$ or coronovirus$ or corono$ virus$ or coronavirinae$ or corona$ virinae$ or Cov or "2019-nCoV$" or 2019nCoV$ or "19-nCoV$" or 19nCoV$ or nCoV2019$ or "nCoV-2019$" or nCoV19$ or "nCoV-19$" or "HCoV-19$" or HCoV19$ or "HCoV-2019$" or HCoV2019$ or "2019 novel$" or Ncov$ or "n-cov" or "SARS-CoV-2$" or "SARSCoV-2$" or "SARSCoV2$" or "SARS-CoV2$" or SARSCov19$ or "SARS-Cov19$" or "SARSCov-19$" or "SARS-Cov-19$" or SARSCov2019$ or "SARS-Cov2019$" or "SARSCov-2019$" or "SARS-Cov-2019$" or SARS2$ or "SARS-2$" or SARScoronavirus2$ or "SARS-coronavirus-2$" or "SARScoronavirus 2$" or "SARS coronavirus2$" or SARScoronovirus2$ or "SARS-coronovirus-2$" or "SARScoronovirus 2$" or "SARS coronovirus2$" or "severe acute respiratory syndrome$")).ti,ab,ot,kw. 3714

11 ((postcovid$ or post covid$ or postcoronavirus$ or postcorona$ virus$ or post coronavirus$ or post corona$ virus$ or postcoronovirus$ or postcorono$ virus$ or post coronovirus$ or post corono$ virus$ or postcoronavirinae$ or postcorona$ virinae$ or post coronavirinae$ or post corona$ virinae$ or postCov or post Cov or postsars$ or post sars$ or "post severe acute respiratory syndrome$" or postncov$ or post ncov$ or posthcov$ or post hcov$) adj3 (syndrome$ or disorder$ or illness$ or sickness$ or disease$ or condition$ or symptom$ or sign$ or prognos$ or followup$ or "follow up$" or feature$ or comorbid$ or "co morbid$" or multimorbid$ or "multi morbid$" or survivor$ or survival$ or risk$ or care$ or convalescen$ or recuperat$ or aftercare$ or ambulatory$ or outpatient$ or "out patient$")).ti,ab,ot,kw. 4125

12 ((ongoing$ or long$ or endur$ or legacy$ or slow$ or gradual$ or protract$ or lengthy$ or chronic$ or persist$ or relaps$ or remit$ or remission$ or residual$ or delay$ or prolong$ or extend$ or linger$ or permanent$ or fluctuat$ or multisystem$ or "multi system$"

or nonrecover$ or "non recover$" or subacute$ or "sub acute$" or lasting$ or continuous$ or continual$ or continuing$ or postacute$ or "post acute$" or postdischarg$ or "post discharg$" or postinfect$ or "post infect$" or postviral$ or "post viral$" or postvirus$ or "post virus$" or "medium$ term$" or mediumterm$) adj4 (sequela$ or illness$ or symptom$ or sign$ or prognos$ or rehab$ or convalescen$ or recuperat$ or followup$ or "follow up$" or feature$) adj10 (covid$ or coronavirus$ or corona$ virus$ or coronovirus$ or corono$ virus$ or coronavirinae$ or corona$ virinae$ or Cov or "2019-nCoV$" or 2019nCoV$ or "19-nCoV$" or 19nCoV$ or nCoV2019$ or "nCoV-2019$" or nCoV19$ or "nCoV-19$" or "HCoV-19$" or HCoV19$ or "HCoV-2019$" or HCoV2019$ or "2019 novel$" or Ncov$ or "n-cov" or "SARS-CoV-2$" or "SARSCoV-2$" or "SARSCoV2$" or "SARS-CoV2$" or SARSCov19$ or "SARS-Cov19$" or "SARSCov-19$" or "SARS-Cov-19$" or SARSCov2019$ or "SARS-Cov2019$" or "SARSCov-2019$" or "SARS-Cov-2019$" or SARS2$ or "SARS-2$" or SARScoronavirus2$ or "SARS-coronavirus-2$" or "SARScoronavirus 2$" or "SARS coronavirus2$" or SARScoronovirus2$ or "SARS-coronovirus-2$" or "SARScoronovirus 2$" or "SARS coronovirus2$" or "severe acute respiratory syndrome$")).ti,ab,ot,kw. 8927
13 or/5-12 26634
14 ((chronic$ or Long or post-acute or longterm or late or persistent$) adj5 (sequela$ or effect$ or symptom$)).ti,ab,ot,kf,kw. 410115
15 sars-related coronavirus/ or exp Severe acute respiratory syndrome coronavirus 2/ 107363
16 exp coronavirus disease 2019/ 376660
17 (coronavirinae/ or betacoronavirus/ or coronavirus infection/) and (epidemic/ or pandemic/) 10708
18 (Coronavirus$ or "covid 19" or 2019-ncov).ti,ab,kw,kf,ot. 432354
19 (2019-ncov or 2019ncov or corona-virus$ or cov19 or cov-19 or 19nCoV or COVID19 or COVID2019 or "Covid 2019").ti,ab,kw,kf,ot. 15890
20 (ncov$ or "sars cov$" or sarscov$ or "sars coronavirus$" or coronovirus$ or corono$ virus$ or "19-nCoV$" or 19nCoV$).ti,ab,kw,kf,ot. 161417
21 (SARS2$ or "SARS-2$" or SARScoronavirus$ or SARS-coronavirus$ or SARScoronovirus$ or SARS-coronovirus$).ti,ab,kw,kf,ot. 2922
22 ("HCoV-19$" or HCoV19$ or "HCoV-2019$" or HCoV2019$).ti,ab,kw,kf,ot. 70
23 ("2019 novel$" or Ncov$).ti,ab,kw,kf,ot. 5863
24 ("Severe Acute Respiratory Syndrome Coronavirus 2" or "Severe Acute Respiratory Syndrome Corona Virus 2").ti,ab,kw,kf,ot. 36929
25 exp "SARS-CoV-2 (lineage B.1.1)"/ 8289
26 exp "SARS-CoV-2 Omicron"/ 7345
27 (omikron or Omicron or "B.1.1.529" or "B11529" or xbb$).af. 13714
28 or/15-27 500310
29 14 and 28 10029
30 13 or 29 30508
31 4 and 30 5751
32 animal/ or animal experiment/ 4726380
33 (rat or rats or mouse or mice or murine or rodent or rodents or hamster or hamsters or pig or pigs or porcine or rabbit or rabbits or animal or animals or dogs or dog or cats or cow or bovine or sheep or ovine or monkey or monkeys).ti,ab,ot,hw. 7733732
34 32 or 33 7733732
35 human experiment/ or exp humans/ 26021849
36 34 not (34 and 35) 5786701
37 31 not 36 5666
38 limit 37 to yr="2022 -Current" 4444
**39 38 not (letter or editorial or conference or "conference abstract" or "conference paper" or "conference review").pt. 3497**

***The Embase strategy was updated on 1.2.24 (204 records) and 1.3.24 (226 records).***

*COVID facet based on terms from:*

World Health Organization (26 May 2021) WHO COVID-19 Database Search Strategy. Systematic search of the COVID-19 literature performed Monday through Friday for the WHO Database. Search strategy as of 26 May 2021. Searches performed by Tomas Allen, Kavita Kothari, and Martha Knuth. Available from: https://www.who.int/docs/default-source/coronaviruse/who-covid-19-database/who-covid-19_sources_searchstrategy_20210526.pdf?sfvrsn=65209cc2_5

Canadian Agency for Drugs and Technologies in Health (2.9.21) CADTH COVID-19 Search Strings: COVID-19 — EMBASE (Internet). Available from: https://covid.cadth.ca/literature-searching-tools/cadth-covid-19-search-strings/

NICE (18 December 2020) [accessed 17.8.21] COVID-19 rapid guideline: managing the long-term effects of COVID-19 [NG188]. Search history record [PDF]. NICE: London. Available from: https://www.nice.org.uk/guidance/ng188/evidence/search-strategies-pdf-8957634445

**Medline ALL (Ovid): 2022-2024/01/23**
**Searched 24.1.24**

1 exp Immunization/ 213463
2 (immunis$ or immuniz$ or immunity or vaccin$ or jab or jabs or shot or shots or booster or boosters or revaccin$ or unvaccin$).ti,ab,ot,kf,kw. 747942
3 or/1-2 805424
4 Post-Acute COVID-19 Syndrome/ 2922
5 (PASC or "chronic covid syndrome$").ti,ab,ot,kf,kw. 867
6 ("post acute" adj2 sequela$ adj2 (covid or coronavirus or coronovirus or "corona virus" or COV)).ti,ab,ot,kw. 342
7 (("long$ term$" or longterm$ or "long$ haul$" or longhaul$ or "long$ tail$" or longtail$ or longduration$ or "long duration$" or longlast$ or "long last$" or longstanding$ or "long standing$" or "medium$ term$" or mediumterm$ or "late effect$" or recurren$ or prolong$ or post-viral$ or chronic$ or postacute or "post acute" or persistent$) adj3 (covid$ or coronavirus$ or corona$ virus$ or coronovirus$ or corono$ virus$ or coronavirinae$ or corona$ virinae$ or Cov or "2019-nCoV$" or 2019nCoV$ or "19-nCoV$" or 19nCoV$ or nCoV2019$ or "nCoV-2019$" or nCoV19$ or "nCoV-19$" or "HCoV-19$" or HCoV19$ or "HCoV-2019$" or HCoV2019$ or "2019 novel$" or Ncov$ or "n-cov" or "SARS-CoV-2$" or "SARSCoV-2$" or "SARSCoV2$" or "SARS-CoV2$" or SARSCov19$ or "SARS-Cov19$" or "SARSCov-19$" or "SARS-Cov-19$" or SARSCov2019$ or "SARS-Cov2019$" or "SARSCov-2019$" or "SARS-Cov-2019$" or SARS2$ or "SARS-2$" or SARScoronavirus2$ or "SARS-coronavirus-2$" or "SARScoronavirus 2$" or "SARS coronavirus2$" or SARScoronovirus2$ or "SARS-coronovirus-2$" or "SARScoronovirus 2$" or "SARS coronovirus2$" or "severe acute respiratory syndrome$")).ti,ab,ot,kw. 9052
8 (longcovid$ or long covid$ or longcoronavirus$ or longcorona$ virus$ or long coronavirus$ or long corona$ virus$ or longcoronovirus$ or longcorono$ virus$ or long coronovirus$ or long corono$ virus$ or longcoronavirinae$ or longcorona$ virinae$ or long coronavirinae$ or long corona$ virinae$ or longCov or long Cov or longsars$ or long sars$ or "long severe acute respiratory syndrome$" or longncov$ or long ncov$ or longhcov$ or long hcov$ or "post-acute covid" or "post-acute corona$" or "post-acute corono$" or "post-acute cov" or "post-acute ncov" or "post-acute sars" or "post-acute severe respiratory syndrome$").ti,ab,ot,kw. 5205

9 ((long$ or endur$ or legacy$ or slow$ or gradual$ or protract$ or lengthy$ or chronic$ or persist$ or relaps$ or remit$ or remission$ or residual$ or delay$ or prolong$ or extend$ or linger$ or permanent$ or fluctuat$ or sequela$ or multisystem$ or "multi system$" or nonrecover$ or "non recover$" or subacute$ or "sub acute$" or lasting$ or continuous$ or continual$ or continuing$ or postacute$ or "post acute$" or postdischarg$ or "post discharg$" or postinfect$ or "post infect$" or postviral$ or "post viral$" or postvirus$ or "post virus$" or "late effect$") adj2 (covid$ or coronavirus$ or corona$ virus$ or coronovirus$ or corono$ virus$ or coronavirinae$ or corona$ virinae$ or Cov or "2019-nCoV$" or 2019nCoV$ or "19-nCoV$" or 19nCoV$ or nCoV2019$ or "nCoV-2019$" or nCoV19$ or "nCoV-19$" or "HCoV-19$" or HCoV19$ or "HCoV-2019$" or HCoV2019$ or "2019 novel$" or Ncov$ or "n-cov" or "SARS-CoV-2$" or "SARSCoV-2$" or "SARSCoV2$" or "SARS-CoV2$" or SARSCov19$ or "SARS-Cov19$" or "SARSCov-19$" or "SARS-Cov-19$" or SARSCov2019$ or "SARS-Cov2019$" or "SARSCov-2019$" or "SARS-Cov-2019$" or SARS2$ or "SARS-2$" or SARScoronavirus2$ or "SARS-coronavirus-2$" or "SARScoronavirus 2$" or "SARS coronavirus2$" or SARScoronovirus2$ or "SARS-coronovirus-2$" or "SARScoronovirus 2$" or "SARS coronovirus2$" or "severe acute respiratory syndrome$")).ti,ab,ot,kw. 14885

10 (("long$ term$" or longterm$ or "long$ haul$" or longhaul$ or "long$ tail$" or longtail$ or longduration$ or "long duration$" or longlast$ or "long last$" or longstanding$ or "long standing$" or "medium$ term$" or mediumterm$ or "post-virus$" or "post-viral") adj3 (covid$ or coronavirus$ or corona$ virus$ or coronovirus$ or corono$ virus$ or coronavirinae$ or corona$ virinae$ or Cov or "2019-nCoV$" or 2019nCoV$ or "19-nCoV$" or 19nCoV$ or nCoV2019$ or "nCoV-2019$" or nCoV19$ or "nCoV-19$" or "HCoV-19$" or HCoV19$ or "HCoV-2019$" or HCoV2019$ or "2019 novel$" or Ncov$ or "n-cov" or "SARS-CoV-2$" or "SARSCoV-2$" or "SARSCoV2$" or "SARS-CoV2$" or SARSCov19$ or "SARS-Cov19$" or "SARSCov-19$" or "SARS-Cov-19$" or SARSCov2019$ or "SARS-Cov2019$" or "SARSCov-2019$" or "SARS-Cov-2019$" or SARS2$ or "SARS-2$" or SARScoronavirus2$ or "SARS-coronavirus-2$" or "SARScoronavirus 2$" or "SARS coronavirus2$" or SARScoronovirus2$ or "SARS-coronovirus-2$" or "SARScoronovirus 2$" or "SARS coronovirus2$" or "severe acute respiratory syndrome$")).ti,ab,ot,kw. 3219

11 ((postcovid$ or post covid$ or postcoronavirus$ or postcorona$ virus$ or post coronavirus$ or post corona$ virus$ or postcoronovirus$ or postcorono$ virus$ or post coronovirus$ or post corono$ virus$ or postcoronavirinae$ or postcorona$ virinae$ or post coronavirinae$ or post corona$ virinae$ or postCov or post Cov or postsars$ or post sars$ or "post severe acute respiratory syndrome$" or postncov$ or post ncov$ or posthcov$ or post hcov$) adj3 (syndrome$ or disorder$ or illness$ or sickness$ or disease$ or condition$ or symptom$ or sign$ or prognos$ or followup$ or "follow up$" or feature$ or comorbid$ or "co morbid$" or multimorbid$ or "multi morbid$" or survivor$ or survival$ or risk$ or care$ or convalescen$ or recuperat$ or aftercare$ or ambulatory$ or outpatient$ or "out patient$")).ti,ab,ot,kw. 3028

12 ((ongoing$ or long$ or endur$ or legacy$ or slow$ or gradual$ or protract$ or lengthy$ or chronic$ or persist$ or relaps$ or remit$ or remission$ or residual$ or delay$ or prolong$ or extend$ or linger$ or permanent$ or fluctuat$ or multisystem$ or "multi system$" or nonrecover$ or "non recover$" or subacute$ or "sub acute$" or lasting$ or continuous$ or continual$ or continuing$ or postacute$ or "post acute$" or postdischarg$ or "post discharg$" or postinfect$ or "post infect$" or postviral$ or "post viral$" or postvirus$ or "post virus$" or "medium$ term$" or mediumterm$) adj4 (sequela$ or illness$ or symptom$ or sign$ or prognos$ or rehab$ or convalescen$ or recuperat$ or followup$ or "follow up$" or feature$) adj10 (covid$ or coronavirus$ or corona$ virus$ or coronovirus$ or corono$ virus$ or coronavirinae$ or corona$ virinae$ or Cov or "2019-nCoV$" or 2019nCoV$ or "19-nCoV$" or 19nCoV$ or nCoV2019$ or "nCoV-2019$" or nCoV19$ or "nCoV-19$" or "HCoV-19$" or HCoV19$ or "HCoV-2019$" or HCoV2019$ or "2019 novel$" or Ncov$ or "n-cov" or "SARS-CoV-2$" or "SARSCoV-2$" or "SARSCoV2$" or "SARS-CoV2$" or SARSCov19$ or

"SARS-Cov19$" or "SARSCov-19$" or "SARS-Cov-19$" or SARSCov2019$ or "SARS-Cov2019$" or "SARSCov-2019$" or "SARS-Cov-2019$" or SARS2$ or "SARS-2$" or SARScoronavirus2$ or "SARS-coronavirus-2$" or "SARScoronavirus 2$" or "SARS coronavirus2$" or SARScoronovirus2$ or "SARS-coronovirus-2$" or "SARScoronovirus 2$" or "SARS coronovirus2$" or "severe acute respiratory syndrome$")).ti,ab,ot,kw. 6706
13 or/4-12 21997
14 ((chronic$ or Long or post-acute or longterm or late or persistent$) adj5 (sequela$ or effect$ or symptom$)).ti,ab,ot,kf,kw. 300919
15 exp Severe acute respiratory syndrome-related coronavirus/ 167482
16 COVID-19/ 253025
17 Coronavirus Infections/ 46122
18 (coronaviridae/ or exp coronavirus/ or betacoronavirus/ or exp betacoronavirus 1/) and (epidemics/ or pandemics/ or Disease Outbreaks/) 76944
19 (Coronavirus$ or "covid 19" or 2019-ncov).ti,ab,kw,kf,ot. 388465
20 (2019-ncov or 2019ncov or corona-virus$ or cov19 or cov-19 or 19nCoV or COVID19 or COVID2019 or "Covid 2019").ti,ab,kw,kf,ot. 10230
21 (ncov$ or "sars cov$" or sarscov$ or "sars coronavirus$" or coronovirus$ or corono$ virus$ or "19-nCoV$" or 19nCoV$).ti,ab,kw,kf,ot. 141131
22 (SARS2$ or "SARS-2$" or SARScoronavirus$ or SARS-coronavirus$ or SARScoronovirus$ or SARS-coronovirus$).ti,ab,kw,kf,ot. 2636
23 ("HCoV-19$" or HCoV19$ or "HCoV-2019$" or HCoV2019$).ti,ab,kw,kf,ot. 66
24 ("2019 novel$" or Ncov$).ti,ab,kw,kf,ot. 5245
25 ("Severe Acute Respiratory Syndrome Coronavirus 2" or "Severe Acute Respiratory Syndrome Corona Virus 2").ti,ab,kw,kf,ot. 37116
26 (omikron or Omicron or "B.1.1.529" or "B11529" or xbb$).af. 10018
27 or/15-26 422651
28 14 and 27 7756
29 13 or 28 24967
30 3 and 29 4424
31 animals/ not (animals/ and humans/) 5155785
32 30 not 31 4394
33 32 not (case reports or clinical conference or comment or editorial or letter).pt. 3846
**34 limit 33 to yr="2022 -Current" 2944**

***The Medline ALL strategy was updated on 1.2.24 (102 records) and 1.3.24 (219 records).***

*COVID facet based on terms from:*

World Health Organization (26 May 2021) WHO COVID-19 Database Search Strategy. Systematic search of the COVID-19 literature performed Monday through Friday for the WHO Database. Search strategy as of 26 May 2021. Searches performed by Tomas Allen, Kavita Kothari, and Martha Knuth. Available from: https://www.who.int/docs/default-source/coronaviruse/who-covid-19-database/who-covid-19_sources_searchstrategy_20210526.pdf?sfvrsn=65209cc2_5

Canadian Agency for Drugs and Technologies in Health (2.9.21) CADTH COVID-19 Search Strings: COVID-19 — EMBASE (Internet). Available from: https://covid.cadth.ca/literature-searching-tools/cadth-covid-19-search-strings/

NICE (18 December 2020) [accessed 17.8.21] COVID-19 rapid guideline: managing the long-term effects of COVID-19 [NG188]. Search history record [PDF]. NICE: London.

Available from: https://www.nice.org.uk/guidance/ng188/evidence/search-strategies-pdf-8957634445

**PubMed (NLM): 2022-2024/01/24**
**Searched 24.1.24**
**https://pubmed.ncbi.nlm.nih.gov/**

**24 #22 AND #23 774**
23 ("2022/01/01"[Date - Publication] : "3000"[Date - Publication]) 3,346,180
22 #20 NOT #21 1,101
21 LETTER[Publication Type] OR EDITORIAL[Publication Type] OR COMMENT[Publication Type] 2,218,993
20 #18 AND #19 1,111
19 pubstatusaheadofprint OR publisher[sb] OR pubmednotmedline[sb] 5,657,369
18 #17 NOT #16 3,711
17 #10 AND #13 3,877
16 #14 NOT (#14 AND #15) 3,770,128
15 Human[tiab] OR humans[tiab] 3,265,545
14 rat[tiab] OR rats[tiab] OR mouse[tiab] OR mice[tiab] OR murine[tiab] OR rodent[tiab] OR rodents[tiab] OR hamster[tiab] OR hamsters[tiab] OR pig[tiab] OR pigs[tiab] OR porcine[tiab] OR rabbit[tiab] OR rabbits[tiab] OR animal[tiab] OR animals[tiab] OR dogs[tiab] OR dog[tiab] OR cats[tiab] OR cow[tiab] OR bovine[tiab] OR sheep[tiab] OR ovine[tiab] OR monkey[tiab] OR monkeys[tiab] 4,725,135
13 #11 OR #12 910,777
12 (immunised[Text Word] OR immunise[Text Word] OR immunisation[Text Word] OR immunisations[Text Word] OR immunized[Text Word] OR immunize[Text Word] OR immunization[Text Word] OR immunizations[Text Word] OR immunity[Text Word] OR vaccine[Text Word] OR vaccines[Text Word] OR vaccination[Text Word] OR vaccinations[Text Word] OR vaccinated[Text Word] OR jab[Text Word] OR jabs[Text Word] OR shot[Text Word] OR shots[Text Word] OR booster[Text Word] OR boosters[Text Word] OR revaccination[Text Word] OR revaccinations[Text Word] OR revaccinated[Text Word] OR unvaccinated[Text Word]) 893,875
11 "Immunization"[Mesh] 213,271
10 #1 OR #2 OR #3 OR #4 OR #5 OR #6 OR #7 OR #8 OR #9 16,571
9 ((ongoing[Text Word] OR long[Text Word] OR endur[Text Word] OR legacy[Text Word] OR slow[Text Word] OR gradual[Text Word] OR protracted[Text Word] OR lengthy[Text Word] OR chronic[Text Word] OR persistent[Text Word] OR relapse[Text Word] OR remitting[Text Word] OR remission[Text Word] OR residual[Text Word] OR delayed[Text Word] OR prolonged[Text Word] OR extended[Text Word] OR lingering[Text Word] OR permanent[Text Word] OR fluctuation[Text Word] OR multisystem[Text Word] OR "multi system"[Text Word] OR nonrecovery[Text Word] OR "non recovery"[Text Word] OR subacute[Text Word] OR "sub acute"[Text Word] OR lasting[Text Word] OR continuous[Text Word] OR continually[Text Word] OR continuing[Text Word] OR postacute[Text Word] OR "post acute"[Text Word] OR postdischarge[Text Word] OR "post discharge"[Text Word] OR postinfection[Text Word] OR "post infection"[Text Word] OR postviral[Text Word] OR "post viral"[Text Word] OR postvirus[Text Word] OR "post virus"[Text Word] OR "medium term"[Text Word] OR mediumterm[Text Word]) AND (sequela[Text Word] OR sequelae[Text Word] OR illness[Text Word] OR symptom[Text Word] OR sign[Text Word] OR symptoms[Text Word] OR signs[Text Word] OR prognosis[Text Word] OR rehab[Text Word] OR convalescence[Text Word] OR recuperation[Text Word] OR followup[Text Word] OR "follow up"[Text Word] OR features[Text Word]) AND (covid[Text Word] OR coronavirus[Text Word] OR "corona virus"[Text Word] OR coronovirus[Text Word] OR "corono virus"[Text Word] OR coronavirinae[Text Word] OR "corona virinae"[Text Word] OR Cov[Text Word] OR

"2019-nCoV"[Text Word] OR "2019nCoV"[Text Word] OR "19- nCoV"[Text Word] OR "19nCoV"[Text Word] OR "nCoV2019"[Text Word] OR omikron[Text Word] OR Omicron[Text Word] OR "B.1.1.529"[Text Word] OR "B11529"[Text Word] OR xbb[Text Word] OR "nCoV-2019"[Text Word] OR "nCoV19"[Text Word] OR "nCoV-19"[Text Word] OR "HCoV-19"[Text Word] OR "HCoV19"[Text Word] OR "HCoV-2019"[Text Word] OR "HCoV2019"[Text Word] OR "2019 novel"[Text Word] OR Ncov[Text Word] OR "n-cov"[Text Word] OR "SARS-CoV-2"[Text Word] OR "SARSCoV-2"[Text Word] OR "SARSCoV2"[Text Word] OR "SARS-CoV2"[Text Word] OR "SARSCov19"[Text Word] OR "SARS-Cov19"[Text Word] OR "SARSCov-19"[Text Word] OR "SARS-Cov-19"[Text Word] OR "SARSCov2019"[Text Word] OR "SARS-Cov2019"[Text Word] OR "SARSCov-2019"[Text Word] OR "SARS-Cov-2019"[Text Word] OR "SARS2"[Text Word] OR "SARS-2"[Text Word] OR "SARScoronavirus2"[Text Word] OR "SARS-coronavirus-2"[Text Word] OR "SARScoronavirus 2"[Text Word] OR "SARS coronavirus2"[Text Word] OR "SARScoronovirus2"[Text Word] OR "SARS-coronovirus-2"[Text Word] OR "SARScoronovirus 2"[Text Word] OR "SARS coronovirus2"[Text Word] OR "severe acute respiratory syndrome"[Text Word] "severe acute respiratory syndrome")) 5,217

8 ((postcovid[Text Word] OR "post covid"[Text Word] OR postcoronavirus[Text Word] OR "postcorona virus"[Text Word] OR "post coronavirus"[Text Word] OR "post corona virus"[Text Word] OR postcoronovirus[Text Word] OR "postcorono virus"[Text Word] OR "post coronovirus"[Text Word] OR "post corono virus"[Text Word] OR postcoronavirinae[Text Word] OR "postcorona virinae"[Text Word] OR "post coronavirinae"[Text Word] OR "post corona virinae"[Text Word] OR postCov[Text Word] OR "post Cov"[Text Word] OR postsars[Text Word] OR "post sars"[Text Word] OR "post severe acute respiratory syndrome"[Text Word] OR postncov[Text Word] OR "post ncov"[Text Word] OR posthcov[Text Word] OR "post hcov"[Text Word]) AND (syndrome[Text Word] OR disorder[Text Word] OR illness[Text Word] OR sickness[Text Word] OR disease[Text Word] OR condition[Text Word] OR symptom[Text Word] OR sign[Text Word] OR symptoms[Text Word] OR signs[Text Word] OR prognosis[Text Word] OR followup[Text Word] OR "follow up"[Text Word] OR features[Text Word] OR comorbidity[Text Word] OR comorbidities[Text Word] OR "co morbidity"[Text Word] OR "co morbidities"[Text Word] OR multimorbidity[Text Word] OR "multi morbidity"[Text Word] OR multimorbidities[Text Word] OR "multi morbidities"[Text Word] OR survivors[Text Word] OR survival[Text Word] OR risk[Text Word] OR risks[Text Word] OR care[Text Word] OR convalescence[Text Word] OR recuperation[Text Word] OR aftercare[Text Word] OR ambulatory[Text Word] OR outpatient[Text Word] OR "out patient"[Text Word] "out patient")) 31

7 (("long term"[Text Word] OR longterm[Text Word] OR "long haul"[Text Word] OR longhaul[Text Word] OR "long tail"[Text Word] OR longtail[Text Word] OR longduration[Text Word] OR "long duration"[Text Word] OR longlasting[Text Word] OR "long lasting"[Text Word] OR longstanding[Text Word] OR "long standing"[Text Word] OR "medium term"[Text Word] OR mediumterm[Text Word] OR "post-virus"[Text Word] OR "post-viral"[Text Word]) AND (covid[Text Word] OR coronavirus[Text Word] OR "corona virus"[Text Word] OR coronovirus[Text Word] OR "corono virus"[Text Word] OR coronavirinae[Text Word] OR "corona virinae"[Text Word] OR Cov[Text Word] OR "2019-nCoV"[Text Word] OR "2019nCoV"[Text Word] OR "19-nCoV"[Text Word] OR "19nCoV"[Text Word] OR "nCoV2019"[Text Word] OR "nCoV-2019"[Text Word] OR "nCoV19"[Text Word] OR "nCoV-19"[Text Word] OR "HCoV-19"[Text Word] OR "HCoV19"[Text Word] OR "HCoV-2019"[Text Word] OR "HCoV2019"[Text Word] OR "2019 novel"[Text Word] OR Ncov[Text Word] OR "n-cov"[Text Word] OR "SARS-CoV-2"[Text Word] OR "SARSCoV-2"[Text Word] OR "SARSCoV2"[Text Word] OR "SARS-CoV2"[Text Word] OR "SARSCov19"[Text Word] OR "SARS-Cov19"[Text Word] OR "SARSCov-19"[Text Word] OR "SARS-Cov-19"[Text Word] OR "SARSCov2019"[Text Word] OR "SARS-Cov2019"[Text Word] OR "SARSCov-2019"[Text Word] OR "SARS-Cov-2019"[Text Word] OR "SARS2"[Text Word] OR "SARS-2"[Text Word] OR omikron[Text Word] OR Omicron[Text Word] OR "B.1.1.529"[Text Word]

OR "B11529"[Text Word] OR xbb[Text Word] OR "SARScoronavirus2"[Text Word] OR "SARS-coronavirus-2"[Text Word] OR "SARScoronavirus 2"[Text Word] OR "SARS coronavirus2"[Text Word] OR "SARScoronovirus2"[Text Word] OR "SARS-coronovirus-2"[Text Word] OR "SARScoronovirus 2"[Text Word] OR "SARS coronovirus2"[Text Word] OR "severe acute respiratory syndrome"[Text Word] "severe acute respiratory syndrome"))
2,257

6 ((long[Text Word] OR enduring[Text Word] OR legacy[Text Word] OR slow[Text Word] OR gradual[Text Word] OR protracted[Text Word] OR lengthy[Text Word] OR chronic[Text Word] OR persistent[Text Word] OR relapsing[Text Word] OR remitting[Text Word] OR remission[Text Word] OR residual[Text Word] OR delayed[Text Word] OR prolonged[Text Word] OR extended[Text Word] OR lingering[Text Word] OR permanent[Text Word] OR fluctuating[Text Word] OR sequelae[Text Word] OR multisystem[Text Word] OR "multi system"[Text Word] OR nonrecovery[Text Word] OR "non recovery"[Text Word] OR subacute[Text Word] OR "sub acute"[Text Word] OR lasting[Text Word] OR continuous[Text Word] OR continual[Text Word] OR continuing[Text Word] OR postacute[Text Word] OR "post acute"[Text Word] OR postdischarge[Text Word] OR "post discharge"[Text Word] OR postinfection[Text Word] OR "post infection"[Text Word] OR postviral[Text Word] OR "post viral"[Text Word] OR postvirus[Text Word] OR "post virus"[Text Word] OR "late effect"[Text Word] OR "late effects"[Text Word]) AND (covid[Text Word] OR coronavirus[Text Word] OR "corona virus"[Text Word] OR coronovirus[Text Word] OR "corono virus"[Text Word] OR coronavirinae[Text Word] OR "corona virinae"[Text Word] OR Cov[Text Word] OR "2019-nCoV"[Text Word] OR "2019nCoV"[Text Word] OR "19-nCoV"[Text Word] OR "19nCoV"[Text Word] OR "nCoV2019"[Text Word] OR "nCoV-2019"[Text Word] OR "nCoV19"[Text Word] OR "nCoV-19"[Text Word] OR "HCoV-19"[Text Word] OR "HCoV19"[Text Word] OR "HCoV-2019"[Text Word] OR "HCoV2019"[Text Word] OR "2019 novel"[Text Word] OR Ncov[Text Word] OR "n-cov"[Text Word] OR "SARS-CoV-2"[Text Word] OR "SARSCoV-2"[Text Word] OR "SARSCoV2"[Text Word] OR "SARS-CoV2"[Text Word] OR "SARSCov19"[Text Word] OR "SARS-Cov19"[Text Word] OR "SARSCov-19"[Text Word] OR "SARS-Cov-19"[Text Word] OR "SARSCov2019"[Text Word] OR "SARS-Cov2019"[Text Word] OR "SARSCov-2019"[Text Word] OR "SARS-Cov-2019"[Text Word] OR "SARS2"[Text Word] OR "SARS-2"[Text Word] OR "SARScoronavirus2"[Text Word] OR "SARS-coronavirus-2"[Text Word] OR "SARScoronavirus 2"[Text Word] OR "SARS coronavirus2"[Text Word] OR omikron[Text Word] OR Omicron[Text Word] OR "B.1.1.529"[Text Word] OR "B11529"[Text Word] OR xbb[Text Word] OR "SARScoronovirus2"[Text Word] OR "SARS-coronovirus-2"[Text Word] OR "SARScoronovirus 2"[Text Word] OR "SARS coronovirus2"[Text Word] OR "severe acute respiratory syndrome"[Text Word] "severe acute respiratory syndrome")) 10,487

5 (longcovid[Text Word] OR "long covid"[Text Word] OR longcoronavirus[Text Word] OR "longcorona virus"[Text Word] OR "long coronavirus"[Text Word] OR "long corona virus"[Text Word] OR longcoronovirus[Text Word] OR "longcorono virus"[Text Word] OR "long coronovirus"[Text Word] OR "long corono virus"[Text Word] OR longcoronavirinae[Text Word] OR "longcorona virinae"[Text Word] OR "long coronavirinae"[Text Word] OR "long corona virinae"[Text Word] OR longCov[Text Word] OR "long Cov"[Text Word] OR longsars[Text Word] OR "long sars"[Text Word] OR "long severe acute respiratory syndrome"[Text Word] OR longncov[Text Word] OR "long ncov"[Text Word] OR longhcov[Text Word] OR "long hcov"[Text Word] OR "post-acute covid"[Text Word] OR "post-acute corona"[Text Word] OR "post-acute corono"[Text Word] OR "post-acute cov"[Text Word] OR "post-acute ncov"[Text Word] OR "post-acute sars"[Text Word] OR "post-acute severe respiratory syndrome"[Text Word]) 5,532

4 (("long term"[Text Word] OR longterm[Text Word] OR "long haul"[Text Word] OR longhaul[Text Word] OR "long tail"[Text Word] OR longtail[Text Word] OR longduration[Text Wo1rd] OR "long duration"[Text Word] OR longlasting[Text Word] OR "long lasting"[Text Word] OR longstanding[Text Word] OR "long standing"[Text Word] OR "medium term"[Text

Word] OR mediumterm[Text Word] OR "late effect"[Text Word] OR recurrence[Text Word] OR prolonged[Text Word] OR post-viral[Text Word] OR chronic[Text Word] OR postacute[Text Word] OR "post acute"[Text Word] OR persistent[Text Word]) AND (covid[Text Word] OR coronavirus[Text Word] OR "corona virus"[Text Word] OR coronovirus[Text Word] OR "corono virus"[Text Word] OR coronavirinae[Text Word] OR "corona virinae"[Text Word] OR Cov[Text Word] OR "2019-nCoV"[Text Word] OR 2019nCoV[Text Word] OR "19-nCoV"[Text Word] OR "19nCoV"[Text Word] OR "nCoV2019"[Text Word] OR "nCoV-2019"[Text Word] OR "nCoV19"[Text Word] OR "nCoV-19"[Text Word] OR "HCoV-19"[Text Word] OR "HCoV19"[Text Word] OR "HCoV-2019"[Text Word] OR "HCoV2019"[Text Word] OR "2019 novel"[Text Word] OR Ncov[Text Word] OR "n-cov"[Text Word] OR "SARS-CoV-2"[Text Word] OR "SARSCoV-2"[Text Word] OR "SARSCoV2"[Text Word] OR "SARS-CoV2"[Text Word] OR "SARSCov19"[Text Word] OR "SARS-Cov19"[Text Word] OR "SARSCov-19"[Text Word] OR "SARS-Cov-19"[Text Word] OR "SARSCov2019"[Text Word] OR "SARS-Cov2019"[Text Word] OR "SARSCov-2019"[Text Word] OR "SARS-Cov-2019"[Text Word] OR "SARS2"[Text Word] OR "SARS-2"[Text Word] OR omikron[Text Word] OR Omicron[Text Word] OR "B.1.1.529"[Text Word] OR "B11529"[Text Word] OR xbb[Text Word] OR "SARScoronavirus2"[Text Word] OR "SARS-coronavirus-2"[Text Word] OR "SARScoronavirus 2"[Text Word] OR "SARS coronavirus2"[Text Word] OR "SARScoronovirus2"[Text Word] OR "SARS-coronovirus-2"[Text Word] OR "SARScoronovirus 2"[Text Word] OR "SARS coronovirus2"[Text Word] OR "severe acute respiratory syndrome"[Text Word] "severe acute respiratory syndrome"))
6,103

3 ("post acute sequela"[Text Word] OR "post acute sequela"[Text Word]) AND (covid[Text Word] OR coronavirus[Text Word] OR coronovirus[Text Word] OR "corona virus"[Text Word] OR COV[Text Word] OR omikron[Text Word] OR Omicron[Text Word] OR "B.1.1.529"[Text Word] OR "B11529"[Text Word] OR xbb[Text Word]) 19

2 (PASC[Text Word] OR "chronic covid syndrome"[Text Word]) 888

1 "Post-Acute COVID-19 Syndrome"[Mesh:NoExp] 2,902

***The PubMed strategy was updated on 1.2.24 (8 records) and 1.3.24 (8 records).***

*COVID facet based on terms from:*

World Health Organization (26 May 2021) WHO COVID-19 Database Search Strategy. Systematic search of the COVID-19 literature performed Monday through Friday for the WHO Database. Search strategy as of 26 May 2021. Searches performed by Tomas Allen, Kavita Kothari, and Martha Knuth. Available from: https://www.who.int/docs/default-source/coronaviruse/who-covid-19-database/who-covid-19_sources_searchstrategy_20210526.pdf?sfvrsn=65209cc2_5

Canadian Agency for Drugs and Technologies in Health (2.9.21) CADTH COVID-19 Search Strings: COVID-19 — EMBASE (Internet). Available from: https://covid.cadth.ca/literature-searching-tools/cadth-covid-19-search-strings/

NICE (18 December 2020) [accessed 17.8.21] COVID-19 rapid guideline: managing the long-term effects of COVID-19 [NG188]. Search history record [PDF]. NICE: London. Available from: https://www.nice.org.uk/guidance/ng188/evidence/search-strategies-pdf-8957634445

*PubMed limit:*
Duffy S, de Kock S, Misso K, Noake C, Ross J, Stirk L. Supplementary searches of PubMed to improve currency of MEDLINE and MEDLINE In-Process searches via Ovid. J Med Libr

Assoc. 2016 Oct;104(4):309-312. doi: 10.3163/1536-5050.104.4.011.
https://www.ncbi.nlm.nih.gov/pmc/articles/PMC5079494/

**Europe PMC, including MedRxiv and bioRxiv preprints (Internet): up to 2024/01/24**
**Searched 24.1.24**
**https://europepmc.org/**

| **Search terms (limited to preprints only)** | **Results** |
|---|---|
| "long-term" OR "long haul" OR "long tail" OR "long duration" OR "long lasting" OR "long standing" OR "medium term" OR "late effects" OR "prolonged" OR "persistent" OR "chronic" OR "post viral" OR "post acute"<br>AND<br>"COVID-19" OR "SARS-CoV-2" OR "coronavirus" OR "COVID" OR "NCOV" OR "omicron" OR "Omicron" OR "B.1.1.529" OR "B11529" OR "xbb"<br>AND<br>"immunised" OR "immunise" OR "immunisation" OR "immunisations" OR "immunized" OR "immunize" OR "immunization" OR "immunizations" OR "immunity" OR "vaccine" OR "vaccines" OR "vaccination" OR "vaccinations" OR "vaccinated" OR "jab" OR "jabs" OR "shot" OR "shots" OR "booster" OR "boosters" OR "revaccination" OR "revaccinations" OR "revaccinated" OR "unvaccinated"<br><br>*All fields* | **17** |
| "COVID-19" OR "SARS-CoV-2" OR "coronavirus" OR "COVID" OR "NCOV" OR omicron OR Omicron OR "B.1.1.529" OR "B11529" OR "xbb"<br>AND<br>"sequelae" OR "sickness"<br>AND<br>"immunised" OR "immunise" OR "immunisation" OR "immunisations" OR "immunized" OR "immunize" OR "immunization" OR "immunizations" OR "immunity" OR "vaccine" OR "vaccines" OR "vaccination" OR "vaccinations" OR "vaccinated" OR "jab" OR "jabs" OR "shot" OR "shots" OR "booster" OR "boosters" OR "revaccination" OR "revaccinations" OR "revaccinated" OR "unvaccinated"<br><br>*All fields* | **2** |
| "PASC" OR "chronic covid syndrome"<br>AND<br>"immunised" OR "immunise" OR "immunisation" OR "immunisations" OR "immunized" OR "immunize" OR "immunization" OR "immunizations" OR "immunity" OR "vaccine" OR "vaccines" OR "vaccination" OR "vaccinations" OR "vaccinated" OR "jab" OR "jabs" OR "shot" OR "shots" OR "booster" OR "boosters" OR "revaccination" OR "revaccinations" OR "revaccinated" OR "unvaccinated" | **0** |
| "post acute sequela" OR "post acute sequela"<br>AND<br>(covid OR coronavirus OR coronovirus OR "corona virus" OR COV OR omicron OR Omicron OR "B.1.1.529" OR "B11529" OR "xbb"<br>AND | **0** |

| | |
|---|---|
| "immunised" OR "immunise" OR "immunisation" OR "immunisations" OR "immunized" OR "immunize" OR "immunization" OR "immunizations" OR "immunity" OR "vaccine" OR "vaccines" OR "vaccination" OR "vaccinations" OR "vaccinated" OR "jab" OR "jabs" OR "shot" OR "shots" OR "booster" OR "boosters" OR "revaccination" OR "revaccinations" OR "revaccinated" OR "unvaccinated" | |
| **Total (including duplicates)** | **19** |

***The Europe PMC strategy was updated on 1.2.24 (0 records) and 1.3.24 (0 records).***

**Latin American and Caribbean Health Sciences Literature (LILACS) (Internet): 2022-2024/01/25**
**Searched 25.1.24**
https://search.bvsalud.org/portal/?lang=en

Searched Title/Abstract/Subject
Limited to 2022-2024/01/25
Limited to LILACS only

| **Search terms** | **Results** |
|---|---|
| ("long-term" OR "long haul" OR "long tail" OR "long duration" OR "long lasting" OR "long standing" OR "medium term" OR "late effects" OR "prolonged" OR "persistent" OR "chronic" OR "post viral" OR "post acute") TITLE, ABSTRACT, SUBJECT<br>AND<br>("COVID-19" OR "SARS-CoV-2" OR "coronavirus" OR "COVID" OR "NCOV" OR "omicron" OR "Omicron" OR "B.1.1.529" OR "B11529" OR "xbb")<br>AND<br>("immunised" OR "immunise" OR "immunisation" OR "immunisations" OR "immunized" OR "immunize" OR "immunization" OR "immunizations" OR "immunity" OR "vaccine" OR "vaccines" OR "vaccination" OR "vaccinations" OR "vaccinated" OR "jab" OR "jabs" OR "shot" OR "shots" OR "booster" OR "boosters" OR "revaccination" OR "revaccinations" OR "revaccinated" OR "unvaccinated") | 63 |
| ("PASC" OR "chronic covid syndrome")<br>AND<br>("immunised" OR "immunise" OR "immunisation" OR "immunisations" OR "immunized" OR "immunize" OR "immunization" OR "immunizations" OR "immunity" OR "vaccine" OR "vaccines" OR "vaccination" OR "vaccinations" OR "vaccinated" OR "jab" OR "jabs" OR "shot" OR "shots" OR "booster" OR "boosters" OR "revaccination" OR "revaccinations" OR "revaccinated" OR "unvaccinated") | 0 |
| ("post acute sequela" OR "post acute sequela") AND ("covid" OR "coronavirus" OR "coronavirus" OR "corona virus" OR COV OR "omicron" OR "Omicron" OR "B.1.1.529" OR "B11529" OR "xbb")<br>AND<br>("immunised" OR "immunise" OR "immunisation" OR "immunisations" OR "immunized" OR "immunize" OR "immunization" OR "immunizations" OR | 0 |

| | |
|---|---|
| "immunity" OR "vaccine" OR "vaccines" OR "vaccination" OR "vaccinations" OR "vaccinated" OR "jab" OR "jabs" OR "shot" OR "shots" OR "booster" OR "boosters" OR "revaccination" OR "revaccinations" OR "revaccinated" OR "unvaccinated") | |
| **Total (including duplicates)** | **63** |

***The LILACS strategy was updated on 1.2.24 (0 records) and 1.3.24 (0 records).***

**Cochrane COVID-19 Study Register (www): 2022-2024/01/25**
**Searched 25.1.24**
https://covid-19.cochrane.org/

Limited to preprints only
Limited 2022-2024/01/25

| Search terms | Results |
|---|---|
| ("long-term" OR "long haul" OR "long tail" OR "long duration" OR "long lasting" OR "long standing" OR "medium term" OR "late effects" OR "prolonged" OR "persistent" OR "chronic" OR "post viral" OR "post acute") AND ("COVID-19" OR "SARS-CoV-2" OR "coronavirus" OR "COVID" OR "NCOV" OR "omicron" OR "Omicron" OR "B.1.1.529" OR "B11529" OR "xbb") ("immunised" OR "immunise" OR "immunisation" OR "immunisations" OR "immunized" OR "immunize" OR "immunization" OR "immunizations" OR "immunity" OR "vaccine" OR "vaccines" OR "vaccination" OR "vaccinations" OR "vaccinated" OR "jab" OR "jabs" OR "shot" OR "shots" OR "booster" OR "boosters" OR "revaccination" OR "revaccinations" OR "revaccinated" OR "unvaccinated") ***Limited to preprints*** | 227 (325 refs) |
| ("PASC" OR "chronic covid syndrome") AND ("immunised" OR "immunise" OR "immunisation" OR "immunisations" OR "immunized" OR "immunize" OR "immunization" OR "immunizations" OR "immunity" OR "vaccine" OR "vaccines" OR "vaccination" OR "vaccinations" OR "vaccinated" OR "jab" OR "jabs" OR "shot" OR "shots" OR "booster" OR "boosters" OR "revaccination" OR "revaccinations" OR "revaccinated" OR "unvaccinated") ***Limited to preprints*** | 6 (9 refs) |
| ("post acute sequela" OR "post acute sequela") AND ("immunised" OR "immunise" OR "immunisation" OR "immunisations" OR "immunized" OR "immunize" OR "immunization" OR "immunizations" OR "immunity" OR "vaccine" OR "vaccines" OR "vaccination" OR "vaccinations" OR "vaccinated" OR "jab" OR "jabs" OR "shot" OR "shots" OR "booster" OR "boosters" OR "revaccination" OR "revaccinations" OR "revaccinated" OR "unvaccinated") ***Limited to preprints*** | 0 |

| **Total (including duplicates)** | **233 (334 refs)** |
|---|---|

***The Cochrane COVID-19 Study Register strategy was updated on 1.2.24 (0 records) and 1.3.24 (0 records).***

**WHO COVID-19 (Internet): 2022-2024/01/24**
**Searched 24.1.24**
https://search.bvsalud.org/global-literature-on-novel-coronavirus-2019-ncov/?lang=en

| **Search terms** | **Results** |
|---|---|
| ("long-term" OR "long haul" OR "long tail" OR "long duration" OR "long lasting" OR "long standing" OR "medium term" OR "late effects" OR "prolonged" OR "persistent" OR "chronic" OR "post viral" OR "post acute") TITLE, ABSTRACT, SUBJECT<br>AND<br>("COVID-19" OR "SARS-CoV-2" OR "coronavirus" OR "COVID" OR "NCOV" OR "omicron" OR "Omicron" OR "B.1.1.529" OR "B11529" OR "xbb")<br>AND<br>("immunised" OR "immunise" OR "immunisation" OR "immunisations" OR "immunized" OR "immunize" OR "immunization" OR "immunizations" OR "immunity" OR "vaccine" OR "vaccines" OR "vaccination" OR "vaccinations" OR "vaccinated" OR "jab" OR "jabs" OR "shot" OR "shots" OR "booster" OR "boosters" OR "revaccination" OR "revaccinations" OR "revaccinated" OR "unvaccinated")<br>***Limited to preprints*** | 674 |
| ("PASC" OR "chronic covid syndrome")<br>AND<br>("immunised" OR "immunise" OR "immunisation" OR "immunisations" OR "immunized" OR "immunize" OR "immunization" OR "immunizations" OR "immunity" OR "vaccine" OR "vaccines" OR "vaccination" OR "vaccinations" OR "vaccinated" OR "jab" OR "jabs" OR "shot" OR "shots" OR "booster" OR "boosters" OR "revaccination" OR "revaccinations" OR "revaccinated" OR "unvaccinated")<br><br>***Limited to preprints*** | 25 |
| ("post acute sequela" OR "post acute sequela") AND ("covid" OR "coronavirus" OR "coronavirus" OR "corona virus" OR COV OR "omicron" OR "Omicron" OR "B.1.1.529" OR "B11529" OR "xbb")<br>AND<br>("immunised" OR "immunise" OR "immunisation" OR "immunisations" OR "immunized" OR "immunize" OR "immunization" OR "immunizations" OR "immunity" OR "vaccine" OR "vaccines" OR "vaccination" OR "vaccinations" OR "vaccinated" OR "jab" OR "jabs" OR "shot" OR "shots" OR "booster" OR "boosters" OR "revaccination" OR "revaccinations" OR "revaccinated" OR "unvaccinated")<br><br>***Limited to preprints*** | 0 |
| **Total (including duplicates)** | **699** |

* WHO COVID-19 database ceased in June 2023, therefore update searches were not necessary.

#### 1.2 Meta-analysis

##### 1.2.1 Feasibility assessment methods

The feasibility assessment determined whether it was possible to conduct pair-wise analyses of the risk of developing long COVID in those who had contracted the Omicron variant of COVID-19 after receiving a vaccine and/or booster dose compared with those who were unvaccinated from the studies retrieved. This assessment determines how comparable the studies are in terms of population, design, and outcomes assessed.

##### 1.2.2 Sensitivity analyses

The following sensitivity analyses were conducted to determine the impact of the addition or removal of specific studies on the pooled risk of long COVID development:

- Leave-one-out analysis
- Exclusion of adolescent and children-only studies[40,45,63]
- Exclusion of pre-print studies[45]
- Exclusion of potentially overlapping populations[41]
- Exclusion of high hospitalization for acute COVID illness (>5%)[39,44]

### 2. Supplementary Results

#### 2.1 Meta-analyses feasibility assessment

Eleven studies were excluded as they did not report risk outcomes[38,43,46-48,51,52,56-58,62]. One study was excluded from the analyses as this included only liver transplant patients[50].Three studies were excluded as they reported on the risk of specific symptoms of COVID that were not comparable with other studies[53,60,66]. Two studies were excluded as they had less than 20 individuals in a comparison group, as such small samples increase uncertainty[49,64,95]. Three studies reported outcomes that were not reported by enough studies to perform a meta-analysis (three or more studies)[54,59,65].

#### 2.2 Supplementary figures and tables

##### Supplementary Figure 1. Egger's test funnel plot for any vaccination (analysis A)

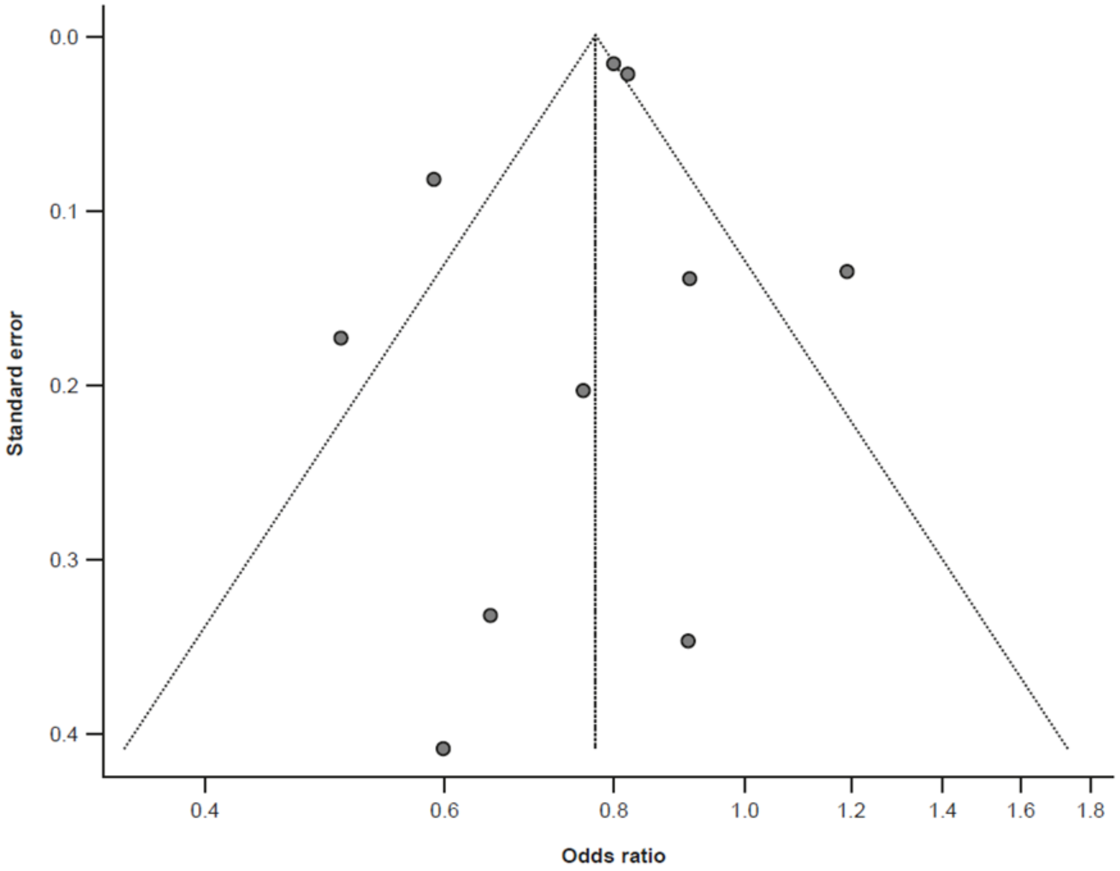

Two separate analyses were performed for "any vaccination" versus no vaccination to account for stratification of results in three studies by primary course vaccination and booster vaccination; analysis A (presented here) included the primary course vaccination data for those stratified studies. In the funnel plot, the central dotted line represents the overall effect, the vertical lines represent the 95% CI, and the dots represent the individual studies. A symmetrical funnel plot indicates a low likelihood of publication bias.

#### Supplementary Figure 2. Egger's test funnel plot for any vaccination (analysis B)

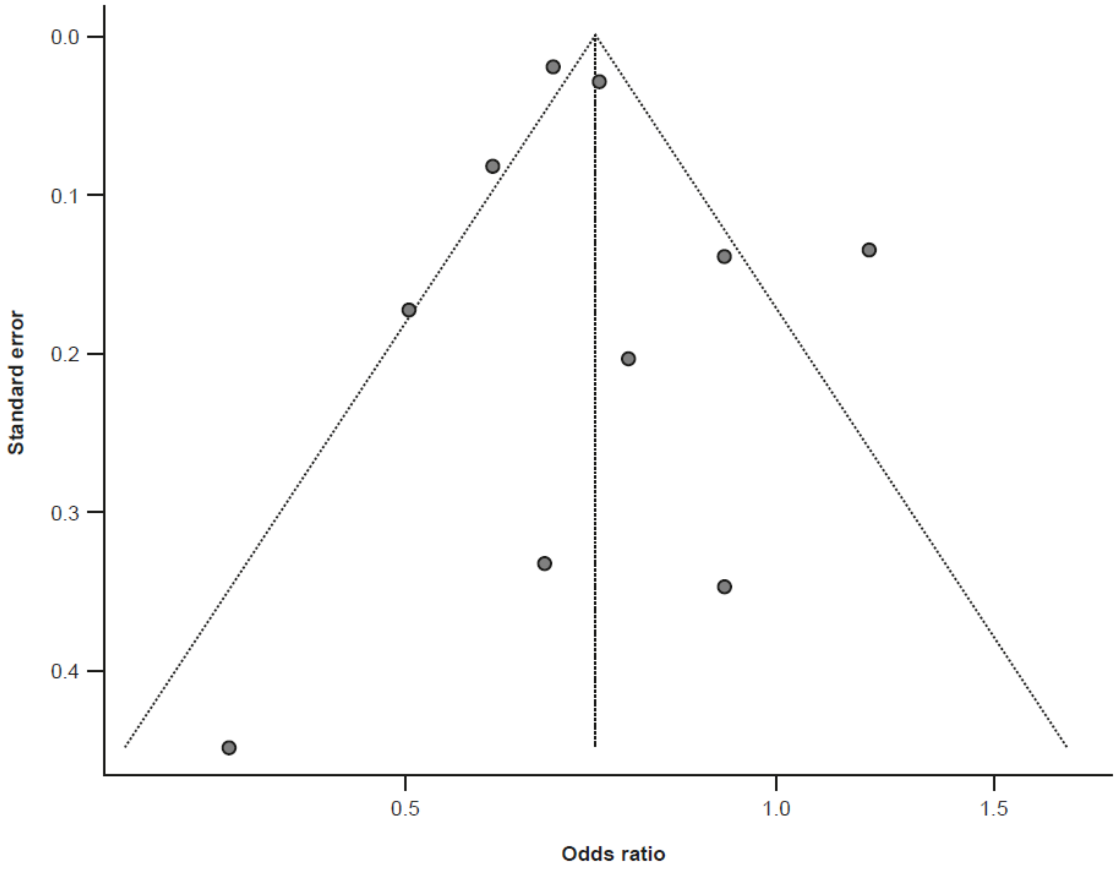

Two separate analyses were performed for "any vaccination" versus no vaccination to account for stratification of results in three studies by primary course vaccination and booster vaccination; analysis B (presented here) included the booster vaccination data for those stratified studies. In the funnel plot, the central dotted line represents the overall effect, the vertical lines represent the 95% CI, and the dots represent the individual studies. A symmetrical funnel plot indicates a low likelihood of publication bias.

#### Supplementary Figure 3. Sensitivity analysis for the effect of any vaccination (analysis A) on the risk of long COVID compared with no vaccination

| Study | RR (95% CI) |
|---|---|
| Omitting Congdon 2023 | 0.77 (0.71; 0.85) |
| Omitting Di Fusco 2023 | 0.78 (0.71; 0.85) |
| Omitting Diexer 2023 | 0.76 (0.70; 0.82) |
| Omitting Hammel 2023 | 0.76 (0.64; 0.90) |
| Omitting Li 2023 | 0.78 (0.71; 0.85) |
| Omitting Lundberg-Morris 2023 | 0.82 (0.76; 0.88) |
| Omitting Mazzitelli 2023 | 0.78 (0.71; 0.85) |
| Omitting Razzaghi 2024 | 0.80 (0.73; 0.87) |
| Omitting Wander 2023 | 0.76 (0.64; 0.91) |
| Omitting Wu 2024 | 0.77 (0.70; 0.84) |
| Total | 0.78 (0.71; 0.85) |

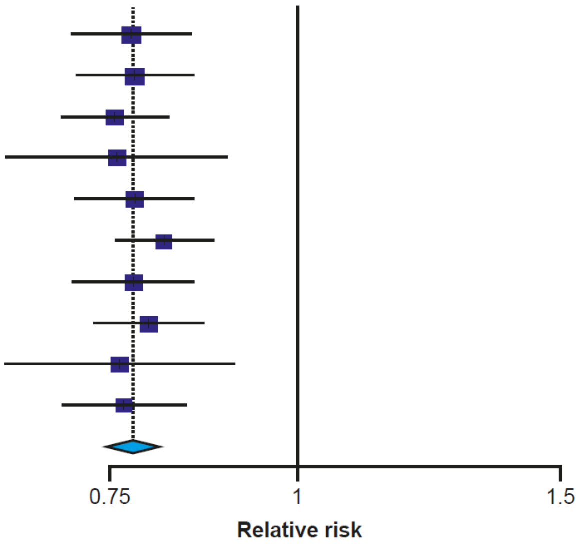

CI, confidence interval; RR, relative risk

Forest plot for the leave-one-out analysis (random effects). The squares and horizontal bars represent the relative risk and 95% CI, respectively, for each study. The dotted line and diamond represent the pooled relative risk and 95% CI, respectively, for all the studies included in the analysis.

#### Supplementary Figure 4. Sensitivity analysis forest plot for the effect of any vaccination (analysis B) on the risk of long COVID compared with no vaccination

| Study | RR (95% CI) |
|---|---|
| Omitting Congdon 2023 | 0.71 (0.64; 0.79) |
| Omitting Di Fusco 2023 | 0.72 (0.65; 0.80) |
| Omitting Diexer 2023 | 0.68 (0.62; 0.74) |
| Omitting Hammel 2023 | 0.72 (0.60; 0.85) |
| Omitting Li 2023 | 0.72 (0.64; 0.80) |
| Omitting Lundberg-Morris 2023 | 0.74 (0.66; 0.83) |
| Omitting Mazzitelli 2023 | 0.71 (0.64; 0.80) |
| Omitting Razzaghi 2024 | 0.73 (0.66; 0.82) |
| Omitting Wander 2023 | 0.73 (0.61; 0.87) |
| Omitting Wu 2024 | 0.70 (0.62; 0.78) |
| Total | 0.71 (0.64; 0.80) |

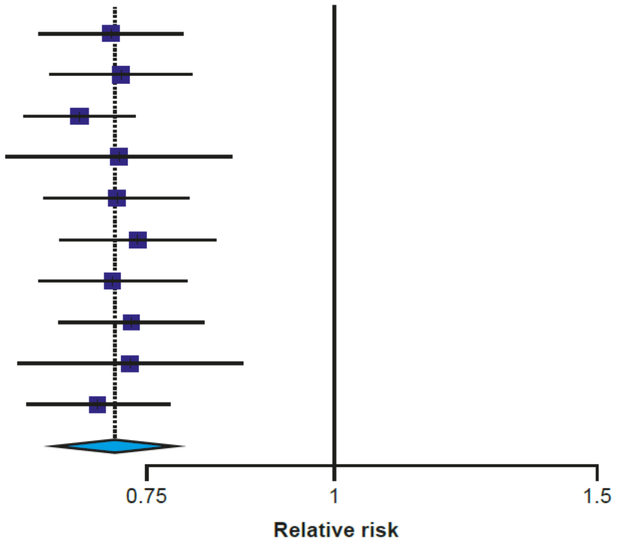

CI, confidence interval; RR, relative risk

Forest plot for the leave-one-out analysis (random effects). The squares and horizontal bars represent the relative risk and 95% CI, respectively, for each study. The dotted line and diamond represent the pooled relative risk and 95% CI, respectively, for all the studies included in the analysis.

#### Supplementary Figure 5. Sensitivity analysis forest plot for the effect of booster vaccination on the risk of long COVID compared with no vaccination

| Study | RR (95% CI) |
|---|---|
| Omitting Di Fusco 2023 | 0.76 (0.66; 0.87) |
| Omitting Diexer 2023 | 0.68 (0.62; 0.75) |
| Omitting Hammel 2023 | 0.74 (0.44; 1.23) |
| Omitting Wander 2023 | 0.78 (0.49; 1.23) |
| Total | 0.74 (0.65; 0.86) |

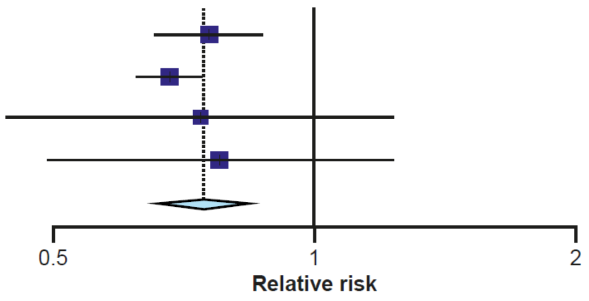

CI, confidence interval; RR, relative risk

Forest plot for the leave-one-out analysis (random effects). The squares and horizontal bars represent the relative risk and 95% CI, respectively, for each study. The dotted line and diamond represent the pooled relative risk and 95% CI, respectively, for all the studies included in the analysis.

#### Supplementary Table 1. Modified NOS questions

| Question | Decision | Stars awarded |
|---|---|---|
| **Selection (max 4)** | | |
| **1. Representativeness of the exposed cohort [vaccinated/COVID-19 positive]** | a) Truly representative of target population (e.g., nationwide database) | 1 |
| | b) Somewhat representative of target population (e.g., city, hospital/hospital system, social media survey) | 1 |
| | c) Selected groups of participants (i.e., by subgroup: sex, race, occupation, insurance coverage, comorbidity, disease severity, ICU treatment status, pre-existing condition). Restricting inclusion criteria to adults does not count as a subgroup | 0 |
| | d) No description of the derivation of the cohort | 0 |
| **2. Selection of the non-exposed cohort [unvaccinated]** | a) Drawn from the same community/database/hospital as the exposed cohort | 1 |
| | b) Drawn from a different source | 0 |
| | c) No description of the derivation of the non-exposed cohort | 0 |

| | | |
|---|---|---|
| **3. Ascertainment of exposure [vaccination]** | a) Diagnosis based upon clinical judgment, or record-linkage (e.g., ICD) | 1 |
| | b) Parental/personal recall only (self-report of vaccination) | 1 |
| | c) No information | 0 |
| **4. Evaluation of whether outcome of interest was present prior to COVID-19 infection/symptoms were worsened post-infection** | a) Yes | 1 |
| | b) No | 0 |
| **Comparability (max 2)** | | |
| **1. Comparability of cohorts on the basis of the design or analysis, or statistical consideration of confounders** | a) Controls/adjusts and/or matches and/or regression analysis for both age and sex | 1 |
| | b) Controls/adjusts and/or matches and/or regression analysis for comorbidities | 1 |
| **Outcome (maximum 3 stars [2 for retrospective])** | | |
| **1. Assessment of outcome [long COVID symptom under investigation]** | a) Validated objective assessment tool (e.g., established cognitive testing or fatigue measurement tool: MoCA, FACIT) for at least one outcome of interest | 1 |
| | b) Structured/systematic interview or questionnaire conducted by trained healthcare or research professional or ICD 10 code | 1 |

| | | |
|---|---|---|
| | c) Unstructured self-report (i.e., open question regarding symptoms) and/or not conducted by trained healthcare or research professional (i.e., self-administered) or not stated | 0 |
| | d) No description | 0 |
| **2. Adequacy of follow-up of cohorts (NA for retrospective)** | a) Complete follow-up; all subjects accounted for | 1 |
| | b) Subjects lost to follow-up unlikely to introduce bias: ≤10% of initial sample size lost, or description provided of those lost | 1 |
| | c) Lost >10% of initial sample size during follow-up, and no description of those lost | 0 |
| | d) No statement | 0 |
| **3. Statistical methodology** | a) Statistical test used to analyze the data clearly described, appropriate and measures of association presented including confidence intervals and probability level (*P*-value) | 1 |
| | b) Statistical test not appropriate, not described, or incomplete | 0 |

FACIT, Functional Assessment of Chronic Illness Therapy; ICD, International Classification of Diseases; ICU, intensive care unit; MoCA, Montreal Cognitive Assessment; NA, not applicable; NOS, Newcastle-Ottawa scale

#### Supplementary Table 2. Quality assessment NOS scores of included studies (excluding cross-sectional studies)

| Author/year | Selection (max 4 stars) | Comparability (max 2 stars) | Outcome (max 3 stars) | Total (out of 9 stars) | Risk of bias |
|---|---|---|---|---|---|
| **Prospective** | | | | | |
| Di Fusco 2023[10] | 3 | 1 | 1 | 5 | Medium |
| Sun 2023[47] | 3 | 1 | 1 | 5 | Medium |
| Ballouz 2023[48] | 4 | 2 | 1 | 7 | Low |
| Gallant 2023[49] | 4 | 2 | 3 | 9 | Low |
| Herting 2023[50] | 3 | 2 | 1 | 6 | Medium |
| Kahlert 2023[51] | 3 | 2 | 2 | 7 | Low |
| Reme 2023[52] | 3 | 2 | 3 | 9 | Low |
| Spiliopoulos 2023[53] | 4 | 2 | 2 | 8 | Low |
| Woldegiorgis 2023[54] | 4 | 2 | 2 | 8 | Low |
| Diexer 2023[55] | 4 | 1 | 2 | 7 | Low |
| de Bruijn 2023[56] | 3 | 2 | 3 | 8 | Low |
| Thaweethai 2023[57] | 4 | 1 | 2 | 7 | Low |
| Nehme 2023[58] | 4 | 2 | 2 | 8 | Low |
| Brown 2023[59] | 4 | 1 | 1 | 6 | Medium |
| Richard 2023[60] | 4 | 2 | 2 | 8 | Low |
| Antonelli 2023[61] | 3 | 2 | 2 | 7 | Low |
| Di Fusco 2024[62] | 3 | 2 | 1 | 6 | Medium |
| **Retrospective** | | | | | |
| Domenech-Montoliu 2023[38] | 3 | 2 | 2 | 7 | Low |
| Mazzitelli 2023[39] | 4 | 1 | 2 | 7 | Low |
| Razzaghi 2024[40] | 3 | 2 | 2 | 7 | Low |
| Hammel 2023[41] | 3 | 1 | 2 | 6 | Medium |
| Congdon 2023[42] | 4 | 2 | 1 | 7 | Low |
| Hedberg 2023[43] | 4 | 2 | 2 | 8 | Low |
| Lundberg-Morris 2023[22] | 4 | 2 | 2 | 8 | Low |
| Wander 2023[44] | 1 | 2 | 2 | 6 | Medium |
| Wu 2024[45] | 2 | 2 | 2 | 6 | Medium |
| Cortellini 2023[46] | 3 | 2 | 2 | 7 | Low |
| Huh 2024[66] | 4 | 2 | 2 | 8 | Low |

NOS, Newcastle-Ottawa Scale

#### Supplementary Table 3. Classification of vaccination status by this review and the definitions reported in the included studies

| Study | Population | Vaccination status | Definition of status reported by study |
|---|---|---|---|
| AlBahrani 2023[64] | Adults | Unvaccinated | No vaccination |
| | | Vaccinated | 2 to 4 doses (not reported in publication, confirmed in correspondence with author) |
| Antonelli 2023[61] | Adults | Primary course | 2 vaccine doses |
| | | Booster dose | 3 vaccine doses |
| Ballouz 2023[48] | Adults | Unvaccinated | No vaccination |
| | | Vaccinated | 1–3 vaccine doses |
| Brown 2023[59] | Adults | No booster dose | <3 vaccine doses |
| | | Booster dose | 3 or more vaccine doses |
| de Bruijn 2023[56] | Adults | Primary course | 2 vaccine doses (or 1 if Ad26.COV2.S) |
| | | Booster dose | 3 vaccine doses |
| Cortellini 2023[46] | Adults | Unvaccinated | No vaccination, or an incomplete primary course |
| | | Vaccinated | Either 2 or 3 vaccine doses |
| Diexer 2023[55] | Adults | Unvaccinated | 0 vaccine doses |
| | | Primary course | 1–2 vaccine doses |
| | | Booster dose | 3+ vaccine doses |
| Di Fusco 2023[10] | Adults | Unvaccinated | No vaccination |
| | | Primary course | Receiving primary series of vaccine |
| | | Booster dose | ≥1 dose after primary series |
| Di Fusco 2024[62] | Adults | No booster dose | No vaccination or monovalent dose >12 months before enrolment |
| | | Booster dose (bivalent) | BioNTech booster vaccine dose post 9/1/22 |
| Domenech-Montoliu 2023[38] | Adults | Primary course | 1–2 doses |
| | | Booster dose | 3 doses |
| Gallant 2023[49] | Adults | Unvaccinated | Unvaccinated |
| | | Vaccinated | ≥2 mRNA doses |
| Hammel 2023[41] | Adults | Unvaccinated | No vaccination |
| | | Primary course | 2 vaccine doses (or 1 if Ad26.COV2.S) |
| | | Booster dose | 3 vaccine doses (or 2 if Ad26.COV2.S) |
| Hedberg 2023[43] | Adults | Unvaccinated | 0–1 vaccine doses |
| | | Vaccinated | 2 or more doses |
| Herting 2023[50] | Adults | Vaccinated | 2 vaccine doses |
| | | Booster dose | ≥3 vaccine doses |
| Huh 2024[66] | Adults | Unvaccinated | 0 vaccine doses |
| | | Primary course | 2 doses of ChAdOx1 or mRNA vaccines |

| | | | |
|---|---|---|---|
| | | Booster dose | 3 vaccine doses (mRNA booster) |
| Kahlert 2023[51] | Adults | Unvaccinated | No vaccination |
| | | Primary course | 1–2 vaccine doses |
| | | Booster dose | ≥3 vaccine doses |
| Li 2023[63] | Children | Unvaccinated | No vaccination |
| | | Vaccinated | ≥2 vaccine doses |
| Lundberg-Morris 2023 | Adults | Unvaccinated | No vaccination |
| | | Vaccinated | 1–5 vaccine doses |
| Mazzitelli 2023[39] | Adults | Unvaccinated | NR |
| | | Vaccinated | NR |
| Mikolajczyk 2023[65] | Adults | Booster dose | 3 vaccine doses (first booster) |
| | | Additional booster dose | 4 vaccine doses |
| Nehme 2023[58] | Adults | Unvaccinated | 0–1 vaccine doses |
| | | Vaccinated | 2–3 doses |
| Razzaghi 2024[40] | Adolescents | Unvaccinated | 0 vaccine doses |
| | | Vaccinated | ≥1 vaccine doses |
| Reme 2023[52] | Adults | Unvaccinated | 0 vaccine doses |
| | | Vaccinated | ≥1 vaccine doses |
| Richard 2023[60] | Adults | Unvaccinated | 0–1 vaccine doses |
| | | Primary course | ≥2 doses of an mRNA vaccine |
| | | Booster dose | ≥1 vaccine dose received after primary course |
| Spiliopoulos 2023[53] | Adults (15+) | Primary course | 2 vaccine doses |
| | | Booster dose | 3 vaccine doses |
| Sun 2023[47] | Adults | Unvaccinated | 0 vaccine doses |
| | | Primary course | 2 doses of BNT162b2 |
| | | Booster dose | ≥1 vaccine dose received after primary course |
| Thaweethai 2023[57] | Adults | Unvaccinated | 0 vaccine doses |
| | | Primary course | 2 vaccine doses (or 1 if Ad26.COV2.S) |
| Wander 2023[44] | Adults | Unvaccinated | No vaccination |
| | | Primary course | 2 vaccine doses (or 1 if Ad26.COV2.S) |
| | | Booster dose | ≥1 vaccine dose received after primary course |
| Woldegiorgis 2023[54] | Adults | No booster dose | 0–2 vaccine doses |
| | | Booster dose | 3 vaccine doses |
| | | Additional booster dose | ≥4 vaccine doses |
| Wu 2024[45] | Adolescents (12–20) | Unvaccinated | No vaccination |
| | | Vaccinated | ≥1 BNT162b2 vaccine doses |

NR, not reported

#### Supplementary Table 4. Summary of long COVID outcomes reported by included studies

| Outcome reported | Number of studies | Studies | Overall summary |
|---|---|---|---|
| Mean number of long COVID symptoms | 5 | Di Fusco 2023, Di Fusco 2024, Domenech-Montoliu 2023, Kahlert 2023, Sun 2023 | Most studies reported that primary course and booster vaccination was associated with a lower number of symptoms compared with no vaccination, and that booster doses were associated with a lower number of symptoms compared with just primary course vaccination. |
| Prevalence or incidence of long COVID | 18 | Antonelli 2023, Ballouz 2023, de Bruijn 2023, Cortellini 2023, Diexer 2023, Di Fusco 2023, Domenech-Montoliu 2023, Hedberg 2023, Huh 2024, Lundberg-Morris 2023, Mikolajczyk 2023, Nehme 2023, Reme 2023, Spiliopoulos 2023, Thaweethai 2023, Wander 2023, Woldegiorgis 2023, Wu 2024 | The majority of studies reported that long COVID prevalence and incidence were lower in those receiving vaccination and further doses (booster, additional dose) compared with no vaccination or vaccination with a lower number of doses. |
| Risk of long COVID development | 20 | AlBahrani 2023, Antonelli 2023, Brown 2023, Congdon 2023, Diexer 2023, Di Fusco 2023, Gallant 2023, Hammel 2023, Herting 2023, Huh 2024, Li 2023, Lundberg-Morris 2023, Mazzitelli 2023, Mikolajczyk 2023, Razzaghi 2024, Richard 2023, Spiliopoulos 2023, Wander 2023, Woldegiorgis 2023, Wu 2024 | Overall, primary course vaccination, booster vaccination, and additional vaccine doses were associated with a numerically or statistically significantly lower risk of long COVID compared with either no vaccination or vaccination with a lower number of doses. |

#### Supplementary Table 5. Reported average number of long COVID symptoms

| Study | Vaccine | Time from infection to long COVID assessment | Vaccine dose | N participants | Mean (SD) number of symptoms | Comparison | *P* value |
|---|---|---|---|---|---|---|---|
| **Omicron-infected patients** | | | | | | | |
| Di Fusco 2023[10] | BNT162b2 | 1 month | Booster dose | 87 | 2.0 (2.3) | Between all | <0.01 |
| | | | Primary course | 86 | 3.1 (3.5) | | |
| | | | Unvaccinated | 155 | 3.7 (4.1) | | |
| | | 3 months | Booster dose | 73 | 1.4 (1.9) | Between all | <0.01 |
| | | | Primary course | 77 | 2.8 (3.5) | | |
| | | | Unvaccinated | 142 | 3.3 (4.0) | | |
| | | 6 months | Booster dose | 67 | 1.1 (1.8) | Between all | <0.001 |
| | | | Primary course | 72 | 2.8 (3.6) | | |
| | | | Unvaccinated | 121 | 3.4 (4.2) | | |
| Domenech-Montoliu 2023[38] | NR (mixed) | 3 months | Booster dose | 90 | 1.5 (2.55) | Between both | NR |
| | | | Primary course | 27 | 2.4 (3.7) | | |
| Kahlert 2023[51] | Mixed | 3 months | Booster dose | 727 | 0.49 (95% CI, 0.41–0.58) | Unvaccinated | 0.295 |
| | | | Primary course | 242 | 0.71 (95% CI, 0.53–0.95) | Unvaccinated | 0.028 |
| | | | Unvaccinated | 102 | 0.36 (95% CI, 0.22–0.60) | NA | NA |
| Sun 2023[47] | BNT162b2 | Up to 6 months | Booster dose | NR | NR (data in figure) | None | NA |
| | | | Primary course | NR | | | |
| | | | Unvaccinated | NR | | | |
| **Long COVID patients only** | | | | | | | |
| Di Fusco 2024[62] | BNT162b2 (bivalent) | 1 month | Booster dose | 260 | 2.4 (3.0) | Between both | 0.006 |
| | | | Unvaccinated | 245 | 2.9 (3.2) | | |
| | | 3 months | Booster dose | 244 | 2.1 (2.4) | Between both | 0.028 |
| | | | Unvaccinated | 226 | 2.8 (3.7) | | |
| | | 6 months | Booster dose | 233 | 2.0 (2.5) | Between both | 0.115 |
| | | | Unvaccinated | 211 | 2.4 (2.7) | | |

CI, confidence intervals; NA, not applicable; NR, not reported; SD, standard deviation

#### Supplementary Table 6. Prevalence of ≥3 and ≥2 symptoms reported by Di Fusco 2024

| Study | Time from infection to long COVID assessment | Number of symptoms | Unvaccinated† | Prevalence | |
|---|---|---|---|---|---|
| | | | | Booster dose | *P* value |
| Di Fusco 2024[62] | 1 month | ≥3 symptoms | 35.5% | 24.2% | 0.006 |
| | 3 months | | 27.0% | 21.3% | 0.15 |
| | 6 months | | 26.5% | 17.2% | 0.017 |
| | | ≥2 symptoms | 37.0% | 25.8% | 0.017 |

†Baseline prevalence in that study (least vaccinated group)

#### Supplementary Table 7. Prevalence of long COVID after Omicron infection by vaccination status

| Study | Vaccine(s) | Time from infection to long COVID assessment | Unvaccinated | Primary course | Booster dose | Additional booster dose | Vaccinated (N doses not specified) | *P* value |
|---|---|---|---|---|---|---|---|---|
| Antonelli 2023[61] | NR (mixed) | ≥4 weeks | - | 4.9% (NR/NR) | 5.1% (NR) | - | - | - |
| | | ≥12 weeks | - | 0.34% (NR/NR) | 0.27% (NR/NR) | - | - | - |
| Ballouz 2023[48] | mRNA | 6 months | Data presented as figure | Data presented as figure | | | | |
| de Bruijn 2023[56] | Mixed | 3 months | - | 28.8% (NR/853) | 27.4% (NR/2,970) | - | - | - |
| Cortellini 2023[46] | Mixed | ≥4 weeks | 9.4% (3/32) | - | - | - | 6.1% (12/196) | 0.49 |
| Diexer 2023[55] | NR (mixed) | ≥12 weeks | No previous infection<br>11.36% (NR/1,093) | No previous infection<br>16.76% (NR/1,780) | No previous infection<br>13.17% (NR/8,518) | - | - | - |
| | | | Previous infection<br>0.43% (NR/235) | Previous infection<br>1.94%% (NR/419) | Previous infection<br>1.56%% (NR/262) | - | - | - |
| Di Fusco 2023[10] | BNT162b2 | 1 month | 43.9% (68/155) | 41.9% (36/86) | 29.9% (26/87) | - | - | ≥0.05 |
| | | 3 months | 42.3% (60/142) | 41.6% (32/77) | 23.3% (17/73) | - | - | <0.05 |
| | | 6 months | 44.6% (54/121) | 37.5% (27/72) | 14.9% (10/67) | - | - | <0.001 |
| Domenech-Montoliu 2023[38] | NR (mixed) | 3 months | - | 13.3% (12/90) | 29.6% (8/27) | - | - | - |
| Hedberg 2023[43] | NR (mixed) | ≥90 days (max 240 days) | 0.20% (104/62,019) | 0.20% (287/147,118) | | - | - | - |
| Lundberg-Morris 2023[22] | Mixed | ≥28 days | 0.6% (202/36,060) | 0.3% (727/224,330) | | | | - |

| | | | | | | | | |
|---|---|---|---|---|---|---|---|---|
| Mikolajczyk 2023[65] | NR (mixed) | ≥4 months | No previous infection<br>29.54% (NR/5,088) | No previous infection<br>33.04% (NR/7,942) | No previous infection<br>34.32% (NR/29,249) | No previous infection<br>23.51% (NR/1,127) | - | - |
| | | | Previous infection<br>6.61% (NR/469) | Previous infection<br>7.16% (NR/1,075) | Previous infection<br>8.67% (NR/784) | Previous infection<br>3.85% (NR/26) | - | - |
| Nehme 2023[58] | mRNA | 3 months | 18.1% (NR/394) | 9.7% (NR/1,338) | | - | - | 0.001 |
| Reme 2023[52] | NR (mixed) | ≥3 months | 0.06% (NR/6,348) | - | - | - | 0.12% (NR/84,347) | - |
| Thaweethai 2023[57] | NR (mixed) | ≥6 months | Acute cohort (enrolled ≤30 days since infection)<br>17% (15/86) | Acute cohort (enrolled ≤30 days since infection)<br>9.7% (195/2,016) | - | - | - | - |
| | | | Post-acute cohort (enrolled >30 days after infection)<br>22% (50/232) | Post-acute cohort (enrolled >30 days after infection)<br>16% (367/2,208) | - | - | - | - |
| Wander 2023[44] | Mixed | ≥1 month (max 12 months) | 33.6% (6,244/104,236) | 35.5% (6,596/134,603) | 30.9% (5,747/150,141) | - | - | - |
| Woldegiorgis 2023[54] | NR (mixed) | 90 days | 19.9% (140/703) | | 18.5% (1,652/8,919) | 4+ doses<br>16.2% (337/2,075) | - | - |

NR, not reported

#### Supplementary Table 8. long COVID risk outcomes

| Study | Vaccine(s) | Time from infection to long COVID assessment | Subgroup | Dose | Vaccine dose | Measurement | Risk (95% CIs) | *P* value |
|---|---|---|---|---|---|---|---|---|
| **Vaccinated (any no. of doses) vs unvaccinated** | | | | | | | | |
| AlBahrani 2023[64] | NR (mixed) | ≥3 months | - | 2–4 | 0 | OR | 0.53 (0.18–1.56) | 0.25 |
| Congdon 2023[42] | NR (mixed) | 4 months | - | 2-–3 | 0 | OR (multivariate) | *0.91 (0.46–1.79)* | 0.78 |
| Gallant 2023[49] | mRNA | 1 month | - | 2+ | 0 | RR (multivariate) | 0.92 (0.79–1.98)[§] | 0.489 |
| Li 2023[63] | mRNA | 3 months | - | 2+ | 0 | OR (multivariate) | 0.65 (0.34–1.25) | 0.20 |
| Lundberg-Morris 2023[22] | Mixed | ≥28 days | - | 1–3 | 0 | HR (partially adjusted) | 0.59 (0.50–0.69) | Significant |
| | | | | | | HR (fully adjusted) | 0.59 (0.50–0.69) | <0.001 |
| Mazzitelli 2023[39] | NR (mixed) | 1 month | - | NR | NR | OR (univariate) | 0.63 (0.44–0.89) | 0.009 |
| | | 3 months | | | | OR (univariate) | 0.76 (0.51– 1.13) | 0.175 |
| Razzaghi 2024[40] | mRNA | ≥1 month | 5–11 yrs; symptom-based or diagnosed long COVID | 1+ | 0 | VE (adjusted) | 23.8% (4.9–39.0) | Significant |
| | | | 5–11 yrs; diagnosed long COVID | 1+ | 0 | VE (adjusted) | 48.2% (-20.8–77.8) | Non-significant |

| | | | | | | | | |
|---|---|---|---|---|---|---|---|---|
| | | | 12–17 yrs; symptom-based or diagnosed long COVID | 1+ | 0 | VE (adjusted) | 49.6% (29.2–64.1) | Significant |
| | | | 5–17 yrs: symptom-based or diagnosed long COVID | 2+ | 0 | VE (adjusted) | 45% (35–53) | Significant |
| | | | 5–11 yrs; symptom-based or diagnosed long COVID | 2+ | 0 | VE (adjusted) | Data presented as figure | NR |
| | | | 12–17 yrs; symptom-based or diagnosed long COVID | 2+ | 0 | VE (adjusted) | Data presented as figure | NR |
| | | | 5–17 yrs: diagnosed long COVID | 2+ | 0 | VE (adjusted) | Data presented as figure | NR |
| | | | 5–11 yrs; diagnosed long COVID | 2+ | 0 | VE (adjusted) | Data presented as figure | NR |
| | | | 12–17 yrs; diagnosed long COVID | 2+ | 0 | VE (adjusted) | Data presented as figure | NR |
| Wu 2024[45] | BNT162b2 | ≥28 days | 5–11 yrs | 1+ | 0 | RR (adjusted) | 1.24 (0.92–1.66) | Non-significant |
| | | | 12–20 yrs | 1+ | 0 | RR (adjusted) | 0.91 (0.69–1.19) | Non-significant |

| | | | | | | | | |
|---|---|---|---|---|---|---|---|---|
| **Primary course vs unvaccinated** | | | | | | | | |
| Di Fusco 2023[10] | BNT162b2 | 6 months | - | 2 | 0 | OR (adjusted) | 0.60 (0.27–1.34) | 0.296 |
| Hammel 2023[41] | Mixed | ≥4 weeks | - | 2 | 0 | HR (adjusted) | 0.82 (0.79–0.86) | Significant |
| Wander 2023[44] | Mixed | ≥1 month | - | 2 | 0 | HR (adjusted) | 0.80 (0.78–0.83) | Significant |
| **Booster dose vs unvaccinated** | | | | | | | | |
| Di Fusco 2023[10] | BNT162b2 | 6 months | - | 3 | 0 | OR (adjusted) | 0.36 (0.15–0.87) | 0.019 |
| Diexer 2023[55] | NR (mixed) | ≥12 weeks | - | 3 | 0 | OR (adjusted) | *1.19 (0.92–1.56)* | Non-significant |
| Hammel 2023[41] | Mixed | ≥4 weeks | - | 3 | 0 | HR (adjusted) | 0.72 (0.68–0.76) | Significant |
| Wander 2023[44] | Mixed | ≥1 month | - | 3 | 0 | HR (adjusted) | 0.66 (0.64–0.69) | Significant |
| **Booster dose vs primary course** | | | | | | | | |
| Antonelli 2023[61] | NR (mixed) | ≥4 weeks | All adults | 3 | 2 | OR (adjusted) | 1.01 (0.85–1.19) | 0.948 |
| | | | Younger adults (18–59 yrs) | 3 | 2 | OR (adjusted) | 0.91 (0.73–1.12) | 0.368 |
| | | | Older adults (60+ yrs) | 3 | 2 | OR (adjusted) | 1.10 (0.83–1.45) | 0.001[‡] |
| | | ≥12 weeks | All ages | 3 | 2 | OR (adjusted) | 0.77 (0.39–1.52) | 0.448 |
| | | | Younger adults (18–59 yrs) | 3 | 2 | OR (adjusted) | 0.73 (0.29–1.86) | 0.511 |
| Di Fusco 2023[10] | BNT162b2 | 6 months | - | 3 | 2 | OR (adjusted) | 0.59 (0.21–1.65) | 0.459 |

| | | | | | | | | |
|---|---|---|---|---|---|---|---|---|
| Diexer 2023[55] | NR (mixed) | ≥12 weeks | - | 3 | 1–2 | OR (adjusted) | *0.78 (0.64–0.93)* | Significant |
| Herting 2023[50] | Mixed | 1 to >3 months | - | 3+ | 2 | OR (multivariate) | 2.94 (0.84–10.28) | 0.085 |
| **Booster dose vs no booster dose** | | | | | | | | |
| Brown 2023[59] | NR (mixed) | 12 weeks | - | 3+ | 0–2 | OR (multivariate) | 0.73 (0.56–0.96) | 0.02 |
| **Additional booster dose vs no booster dose** | | | | | | | | |
| Woldegiorgis 2023[54] | NR (mixed) | 90 days | - | 4+ | 0-2 | RR (adjusted) | *0.63 (0.52– 0.77)* | <0.001[†] |
| **Additional booster dose vs booster dose** | | | | | | | | |
| Mikolajczyk 2023[65] | NR (mixed) | ≥4 months | - | 4 | 3 | OR (multivariate) | 0.52 (0.41–0.61) | Significant |
| Woldegiorgis 2023[54] | NR (mixed) | 90 days | - | 4+ | 3 | RR (adjusted) | *0.71 (0.63–0.77)* | <0.001[†] |

CI, confidence interval; HR, hazard ratio; NR, not reported; OR, odds ratio; RR, relative risk; VE, vaccine effectiveness

Data in italics have had their direction adjusted to match that of other studies (risk of higher number of doses compared with lower number of doses/no doses)

[†]*P*-value reported for overall comparison of ≥4 doses, 3 doses, and 0–2 doses; [‡]Likely error in publication; [§]Vaccine dose comparison unclear and assumed to be comparing vaccinated with unvaccinated; the corresponding author has been contacted for clarification

#### Supplementary Table 9. Table of data used in the five meta-analyses

| Study | Vaccine dose comparison in study | Risk data (95% CIs) included in meta-analysis |
|---|---|---|
| **Vaccinated vs unvaccinated (A)** | | |
| Congdon 2023[42] | Vaccinated (2–3 doses) vs no vaccination | OR 0.91 (0.46–1.79)† |
| Di Fusco 2023[10] | Primary course vs no vaccination | OR 0.60 (0.27–1.34) |
| Diexer 2023[55] | Booster dose (3+) vs no vaccination | OR 1.19 (0.92–1.56)† |
| Hammel 2023[41] | Primary course vs no vaccination | HR 0.82 (0.79–0.86) |
| Li 2023[63] | Vaccinated (≥2 doses) vs no vaccination | OR 0.65 (0.34–1.25) |
| Lundberg-Morris 2023[22] | Vaccinated (1–5 doses) vs no vaccination | HR 0.59 (0.50–0.69) |
| Mazzitelli 2023[39] | Vaccinated (doses NR) vs no vaccination | OR 0.76 (0.51–1.13) |
| Razzaghi 2024[40] | Vaccinated (≥1 dose) vs no vaccination | VE 0.50 (0.36–0.71) |
| Wander 2023[44] | Primary course vs no vaccination | HR 0.80 (0.78– 0.83) |
| Wu 2024[45] | Vaccinated (≥1 dose) vs no vaccination | RR 0.91 (0.69– 1.19) |
| **Vaccinated vs unvaccinated (B)** | | |
| Congdon 2023[42] | Vaccinated (2–3 doses) vs no vaccination | OR 0.91 (0.46–1.79)† |
| Di Fusco 2023[10] | Booster dose vs no vaccination | OR 0.36 (0.15– 0.87) |
| Diexer 2023[55] | Booster dose vs no vaccination | OR 1.19 (0.92–1.56)† |

| | | |
|---|---|---|
| Hammel 2023[41] | Booster dose vs no vaccination | HR 0.72 (0.68–0.76) |
| Li 2023[63] | Vaccinated (≥2 doses) vs no vaccination | OR 0.65 (0.34–1.25) |
| Lundberg-Morris 2023[22] | Vaccinated (1–5 doses) vs no vaccination | HR 0.59 (0.50–0.69) |
| Mazzitelli 2023[39] | Vaccinated (doses NR) vs no vaccination | OR 0.76 (0.51–1.13) |
| Razzaghi 2024[40] | Vaccinated (≥1 dose) vs no vaccination | VE 0.50 (0.36–0.71) |
| Wander 2023[44] | Booster dose vs no vaccination | HR 0.66 (0.64–0.69) |
| Wu 2024[45] | Vaccinated (≥1 dose) vs no vaccination | RR 0.91 (0.69–1.19) |
| **Primary course vs unvaccinated** | | |
| Di Fusco 2023[10] | Primary course vs no vaccination | OR 0.60 (0.27–1.34) |
| Hammel 2023[41] | Primary course vs no vaccination | HR 0.82 (0.79–0.86) |
| Wander 2023[44] | Primary course vs no vaccination | HR 0.80 (0.78–0.83) |
| **Booster dose vs unvaccinated** | | |
| Di Fusco 2023[10] | Booster dose vs no vaccination | OR 0.36 (0.15–0.87) |
| Diexer 2023[55] | Booster dose vs no vaccination | OR 1.19 (0.92–1.56) |
| Hammel 2023[41] | Booster dose vs no vaccination | HR 0.72 (0.68–0.76) |
| Wander 2023[44] | Booster dose vs no vaccination | HR 0.66 (0.64–0.69) |
| **Booster dose vs primary course** | | |
| Antonelli 2023[61] | Booster dose vs primary course | OR 0.77 (0.39–1.52) |
| Di Fusco 2023[10] | Booster dose vs primary course | OR 0.59 (0.21–1.65) |

| | | |
|---|---|---|
| Diexer 2023[55] | Booster dose vs primary course | OR 0.78 (0.64–0.93) |

CI, confidence interval; HR, hazard ratio; NR, not reported; OR, odds ratio; RR, relative risk; VE, vaccine effectiveness

†Directionality adjusted by reviewers

Two separate analyses were performed for “any vaccination” versus no vaccination to account for stratification of results in three studies by primary course vaccination and booster vaccination; analysis A included the primary course vaccination data for those stratified studies and analysis B included the booster vaccination data for those stratified studies.

#### Supplementary Table 10. Sensitivity analyses and test of heterogeneity for the effect of any vaccination (analysis A) on the risk of long COVID compared with no vaccination (random effects)

| Sensitivity analysis | Risk ratio (95% confidence intervals) | Q | Degrees of freedom | Q: *P* value | $I^2$ |
|---|---|---|---|---|---|
| Exclude potential population overlap | 0.76 (0.64–0.90) | 31.19 | 8 | 0.0001 | 74.3% |
| Exclude children/adolescents | 0.79 (0.72–0.87) | 24.30 | 6 | 0.0005 | 75.3% |
| Exclude preprint | 0.77 (0.70–0.84) | 31.9 | 8 | <0.0001 | 74.9% |
| Exclude high hospitalization rate | 0.76 (0.63–0.92) | 32.58 | 7 | <0.0001 | 78.5% |

$I^2$, quantifying heterogeneity; Q, test of heterogeneity

#### Supplementary Table 11. Sensitivity analyses and test of heterogeneity for the effect of any vaccination (analysis B) on the risk of long COVID compared with no vaccination (random effects)

| Sensitivity analysis | Risk ratio (95% confidence intervals) | Q | Degrees of freedom | Q: *P* value | $I^2$ |
|---|---|---|---|---|---|
| Exclude potential population overlap | 0.72 (0.60–.85) | 31.71 | 8 | 0.0001 | 74.8% |
| Exclude children/adolescents | 0.72 (0.64–0.80) | 29.74 | 6 | <0.0001 | 79.8% |
| Exclude preprint | 0.70 (0.62–0.78) | 32.73 | 8 | <0.0001 | 75.6% |
| Exclude high hospitalization rate | 0.72 (0.60–0.88) | 29.79 | 7 | 0.0001 | 76.5% |

$I^2$, quantifying heterogeneity; Q, test of heterogeneity

#### Supplementary Table 12. Sensitivity analyses and test of heterogeneity for the effect of any booster vaccination on the risk of long COVID compared with no vaccination (random effects)

| Sensitivity analysis | Risk ratio (95% confidence intervals) | Q | Degrees of freedom | Q: *P* value | $I^2$ |
|---|---|---|---|---|---|
| Population overlap | 0.74 (0.44–1.23) | 20.66 | 2 | <.0001 | 90.3% |

$I^2$, quantifying heterogeneity; Q, test of heterogeneity
